# Cell-type-resolved somatic variant discovery from bulk long-read sequencing

**DOI:** 10.64898/2026.09.01.26361966

**Authors:** Yilei Fu, Caoimhe Morley, Lauren M. Masters, Adam C. English, Yiming Zhu, Abraham G. Moller, Luis F. Paulin, Ben Thompson, Ester Kalef-Ezra, George Weissenberger, Helen Shen, Melissa Meredith, Arianna Manini, Dominic Horner, Xylena Reed, Donna Muzny, Zane Jaunmuktane, Ziad M. Khan, Heer Mehta, Winston Timp, Kimberley Billingsley, Graham S. Erwin, Christos Proukakis, Fritz J. Sedlazeck

## Abstract

Somatic mutations arise throughout life, with functional consequences tied to the cell populations in which they occur. Genome-wide studies measure somatic variations in bulk tissue, whereas single-cell approaches resolve cell identity but provide limited sensitivity for complex alleles. Here we developed SniffCell, which uses DNA methylation carried on native long reads to assign somatic variant-supporting molecules to methylation-resolvable cell types. SniffCell builds cell-type-discriminatory methylation signatures across eight tissues, assigns long reads to cell types, and provides cell-type-specific variant calling. Across peripheral blood mononuclear cells and brain benchmarks, SniffCell recovered sorted cell identities and validated cell-type-specific variant assignments using purified immune-cell, neuronal, and oligodendrocyte fractions. In blood, SniffCell recovered lineage-restricted antigen receptor rearrangements and localized a somatic tandem-repeat expansion to T cells. In the frontal cortex, SniffCell identified recurrent neuron-specific tandem-repeat expansions in genes including *FGF14, LRRC7* and *SH3RF3*. Across three brain cohorts comprising 172 donors, recurrent neuron-associated expansions were enriched for GAA-rich motifs. In donors with matched blood, and diverged more strongly from the inherited repeat length, whereas oligodendrocyte-associated alleles more often tracked it. SniffCell transforms native bulk long-read genomes into a cell-type-aware resource for somatic variant discovery and reveals recurrent somatic instability in human tissues at cell-type resolution.

## Main

Somatic mutations arise throughout development and aging, creating somatic mosaicism, the accumulation of genetic differences among cells of the same individual^1^. Their impact depends on cellular context, as the same mutation can have very different consequences in neurons, glia, immune cells or epithelial cells^2–4^. Yet most genome-wide studies of somatic variation report variants without resolving which cell population carries them^5^. This missing cellular context limits interpretation across human tissues and can obscure disease-relevant variants that contribute to unexplained phenotypic variation.

Single-cell whole genome sequencing (scWGS) offers one route to cellular resolution^6^, but it is not a replacement for bulk genome sequencing. Current approaches remain costly and difficult to scale across large cohorts and multiple tissues, and whole-genome amplification (WGA) introduces allelic imbalance, chimeras, dropout and false positives that, together with low per-cell coverage, limit sensitivity for somatic variant detection^7^. ScWGS requires tissue dissociation and fluorescence-activated cell sorting (FACS), which further bias which cells are recovered since some cell types or states are too large, too small, too fragile or too difficult to release intact^8^. In the brain, where nuclear isolation is required prior to fluorescence-activated nuclear sorting (FANS), certain important cell types such as dopaminergic neurons do not have highly specific nuclear markers. Additionally, most single-cell genome methods resolve substitutions or large copy-number changes but cannot readily characterize larger insertions, deletions, structural variants (SVs) and tandem repeat (TR) expansions, many of which are associated with neurological disease (e.g., *HTT, FMR1*)^9,10^. Long-read scWGS has the potential to resolve all variant classes, but SV detection is still limited by chimeras^11^. Thus, scWGS is a powerful tool to discover heterogeneity, but remains poorly suited to comprehensive detection of complex and repetitive variation affecting genes or pathways associated with disease.

Bulk long-read sequencing offers comprehensive nucleotide variant detection (including SV, TR and complex/repetitive regions) while simultaneously measuring DNA methylation on the same molecules^12,13^. Critically, native long reads preserve genetic variation and DNA methylation on the same physical molecule^14^. This creates an opportunity to infer the cellular origin of a variant directly from the variant-supporting read. Long-read methylation has previously been used for tissue-of-origin inference and cell-type deconvolution from bulk sequencing data^15–17^. More recently, read-level methylation has been used to reconstruct cell-type-specific methylation profiles from blood and to distinguish tumor-derived from non-tumor molecules in cancer^18^. However, these studies have focused primarily on epigenetic profiling or binary tumor-non-tumor classification rather than assigning complex somatic variants to multiple defined cell populations across heterogeneous tissues.

Here we introduce SniffCell, a framework that uses cell-type-specific DNA methylation as an endogenous barcode to assign variant-supporting long reads to methylation-resolvable cellular origin in bulk sequencing data. SniffCell builds dense, tissue-specific reference atlases of cell-type-discriminatory methylation regions, assigns individual reads to cell classes, and integrates these assignments with the detection of single nucleotide variations (SNVs), small insertions and deletions (indels), tandem repeats (TRs) and structural variations (SVs) to generate cell-type-aware mutational maps from standard bulk long-read sequencing. We validate SniffCell in peripheral blood mononuclear cells (PBMCs) and human frontal cortex using matched sorted populations, demonstrating strong concordance between methylation-based read assignments and experimentally purified cell identities. In blood, SniffCell resolves a T cell-enriched deletion at the TRB locus, and in the brain it identifies recurrent neuron- and oligodendrocyte-associated TR expansions, including neuron-specific somatic expansions. Replication in an independent brain cohort further supported recurrent, cell-type-specific TR instability at motif-selective loci. Together, these results establish bulk long-read sequencing as a scalable approach for cell-type-aware discovery of somatic SVs and TRs without requiring additional single-cell experiments.

### SniffCell links somatic variants to cell type from bulk long-read genomes

SniffCell assigns somatic variants to cell types by linking variant-supporting reads to cell-type-specific methylation. A prebuilt ctDMR reference atlas is available and SniffCell enables the construction of custom reference atlases. This atlas is built from cell- or nuclei-sorted, cell-type-specific DNA methylation profiles (**Figure 1a**). For this study, we built a novel reference methylation atlas from Loyfer et al.’s DNA methylation data^14^. For processing long-read sequencing data from bulk samples, SniffCell begins by associating methylation signals from aligned long reads at overlapping ctDMR reference loci to assign a cell type to each read (**Figure 1b**). In cases where a read cannot be assigned a cell-type-group of origin, SniffCell assigns it to a cell-type-group of origin or leaves it unassigned (**Figure 1b**). For variant calling, each group of reads is analyzed independently, simplifying the discovery of overlapping variants across cell types, after which the resulting calls are consolidated into a standard VCF. Finally, SniffCell provides cell-type annotations for discovered somatic variants in the VCF, enabling standard downstream analyses such as functional annotation. To enable manual inspection, SniffCell provides an HTML report with schematic visualizations that allow users to review each variant, its supporting reads and the evidence underlying the cell-type assignment (**Figure 1c**). Overall, SniffCell is capable of providing cell-type specific variants across eight tissues using its default ctDMR catalog (**Figure 1d**).

**Figure 1:**
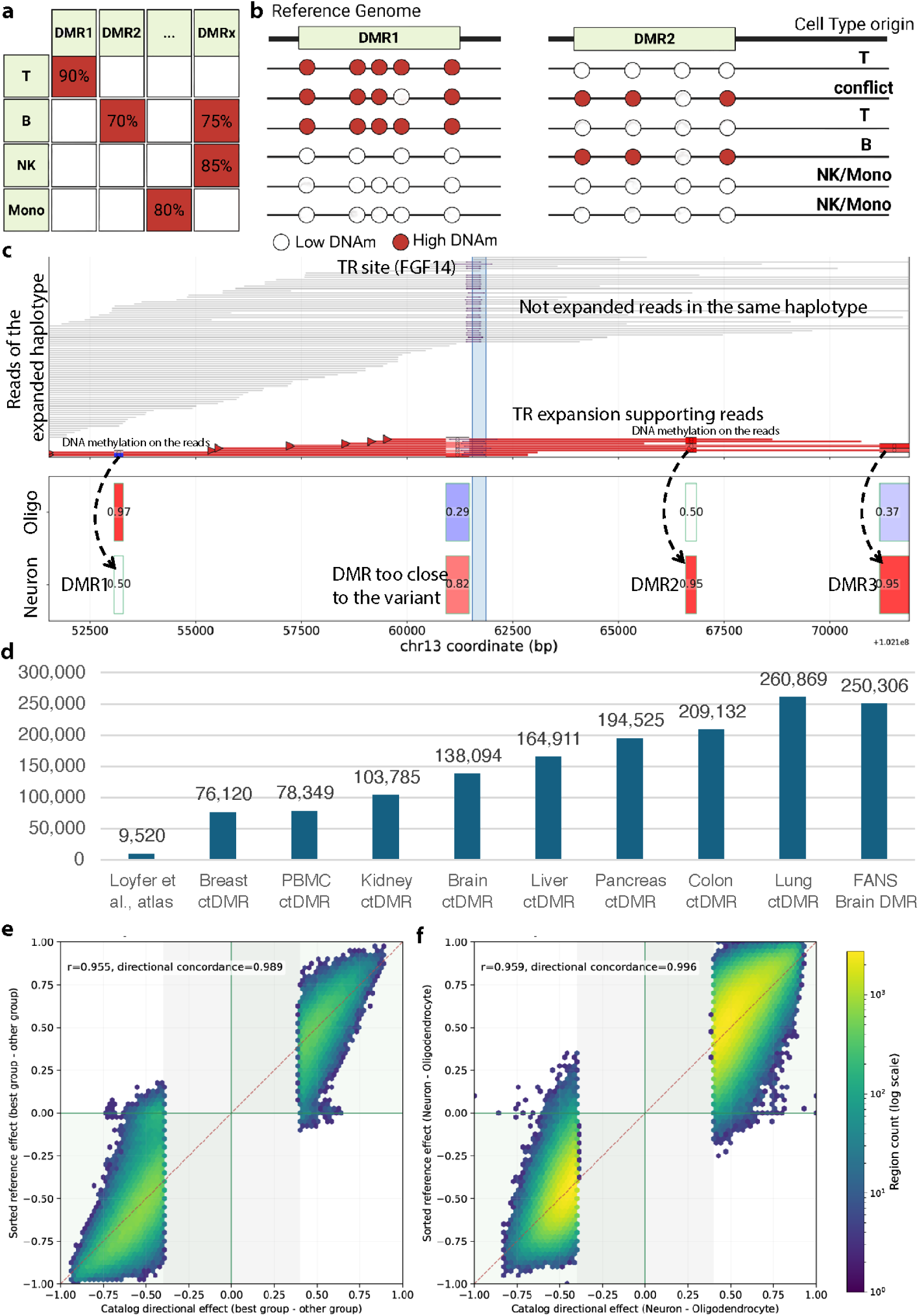
Overview of SniffCell framework, re-processing of the DNA methylation reference atlas, and sorting validation. **a.** SniffCell reference atlas links cell-type-specific differentially methylated regions (ctDMR) to methylation-resolvable cell types. **b.** SniffCell inspects methylation signals to infer the cell-type origin of long reads and collects cell-type-specific subsets of reads for variant calling. **c.** Example SniffCell report at FGF14 showing ctDMRs and TR-expansion-carrying reads supporting a somatic TR expansion. The top panel shows the reads from the bulk BAM files, where the grey lines indicate the reads that do not carry the repeat expansion while the red lines indicate the reads that carry the repeat expansion. The color blocks on the red reads represent the DNA methylation level of that read within a ctDMR. Bottom panel: The DNA methylation of the read at ctDMR corresponds to the ctDMR from the atlas, so that we assign the reads from the bulk sample to the corresponding cell type. The numbers in the atlas blocks represent the mean DNA methylation value of the ctDMR from the cell types in the atlas. In this example, three independent ctDMRs assign all the TR expansion-carrying reads to neurons. **d.** Number of tissue-specific ctDMRs identified by SniffCell across eight tissues. The original Loyfer et al. atlas^14^ and ctDMRs derived independently from the FANS brain samples are shown for comparison. **e.** Comparison of ctDMR methylation differences between two PBMC cell-type groups. The x-axis shows the methylation difference measured in bulk SniffCell assignments and the y-axis the corresponding difference measured in FACS-sorted samples. **f.** Corresponding comparison between neurons and oligodendrocytes in brain samples using FANS-sorted data. x: the ctDMR region’s DNA methylation delta in two brain samples’ neuron versus oligodendrocyte. y: neuron and oligodendrocyte’s DNA methylation delta in the same ctDMR region reported from FANS samples.

Within SniffCell results, a variant is considered somatic when it is consistently restricted to, or strongly skewed toward, a cell-type-assigned or a cell-type-group-assigned (e.g., lymphocytes in blood) reads population. This follows from basic inheritance logic: A constitutive heterozygous variant is expected to be represented across cell types, whereas strong and reproducible enrichment of variant-supporting molecules within one cell-type assignment is consistent with somatic restriction. SniffCell therefore combines cell-type skew with coverage, read support and variant-level validation rather than interpreting single-cell-type absence alone as evidence of somatic origin.

#### Expanded methylation markers enable accurate read-level cell assignment

SniffCell was benchmarked at three levels: DNA methylation marker directionality, read-level assignment and variant-level cell-type assignment. Marker directionality tested whether the expanded ctDMR atlas retained expected methylation differences, read-level assignment tested whether cell-sorted reads were classified into the correct cell-type or cell-group, and variant-level validation tested whether variant-supporting reads were recovered in the corresponding FANS or purified PBMC fraction.

The resolution of genome-wide cell-type inference depends on the spatial density and accuracy of a ctDMR reference atlas (**Supplementary Fig. 1, Supplementary Table 1**). As the number of ctDMR loci increases, the probability that a variant-spanning long read carries enough cell-type-informative methylation sites also increases. To accomplish this, we expanded the prior ctDMR atlas by using the sequenced tissue type to restrict comparisons to the expected set of resolvable cell classes within each tissue. Across eight human tissues (Blood/PBMC, Brain, Kidney, Breast, Liver, Colon, Pancreas, and Lung, **Figure 1d**), SniffCell identified a set of 76,120 to 260,869 ctDMRs, expanding the reference atlas by 8 to 27.4 fold relative to the 9,520-region reference atlas from Loyfer et al. (Loyfer atlas)^14^. Each resulting ctDMR includes at least three cell-type-specific CpGs (**methods**) and their average sizes among tissue types range from 165bp to 261bp (**Supplementary Fig. 2B**). All markers in the SniffCell ctDMR reference atlas that overlapped the Loyfer atlas maintained the expected cell-type classification. Most importantly, the SniffCell ctDMR reference atlas increased marker density within comparable cell-class definitions rather than imposing unsupported subcell-type distinctions. Thus, SniffCell operates at the same practical cell-type resolution as the Loyfer atlas, while providing substantially more informative methylation sites for read-level assignment (**Supplementary Fig. 2D**).

To validate the SniffCell ctDMR reference atlas, we benchmarked against methylation truth sets obtained by separating cell types using fluorescence-activated cell or nuclear sorting (FACS/FANS) from two healthy PBMC controls and four brain samples. The PBMC cohort included two healthy controls (pbmc1, pbmc2). We sorted PBMCs into T cells, B cells, natural killer (NK) cells, and monocytes. The brain cohort included two controls (bcontrol1, bcontrol2) and two samples from individuals with neurodegenerative disease: Parkinson’s disease (PD; bparkinsons) and multiple system atrophy (MSA; bmsa). Brain nuclei were sorted into neuronal and oligodendrocytic fractions, the two most abundant cell types. For each sample, we performed ONT whole-genome-sequencing of the sorted cells and bulk tissue (see **methods, Supplementary Fig. 3, Supplementary Table 2**). The benchmarking procedure assessed concordance between SniffCell atlas-derived ctDMR directional effects and methylation differences measured in the sorted reference samples. Across PBMC and brain datasets, the SniffCell reference atlas showed strong agreement with methylation signals of the sorted truth sets, with Pearson correlations ranging from 0.939 to 0.972 in PBMC and from 0.952 to 0.971 in brain. Directional concordance was similarly high, ranging from 0.982 to 0.996 in PBMC and from 0.994 to 0.998 in brain, supporting robust recovery of both the magnitude and direction of cell-type-specific methylation differences (**Figure 1e, f; Supplementary Fig. 4A-F**).

Given our extended SniffCell ctDMR reference atlas, we next assessed the SniffCell discovery module’s ability (**methods**) to deconvolute reads across cell-types. We compared the DNA methylation of each CpG supported by ≥5 reads per cell-type to the corresponding FACS/FANS-sorted reads. SniffCell cell-type-derived methylation across these cell-type-specific BAM signals closely matched the corresponding sorted methylation profiles. In the PBMC samples, the correlation between methylation values ranged from 0.967-0.974 for monocytes and 0.954-0.967 for lymphocytes (T, B and NK cells) (**Figure 2a, Supplementary Fig. 5**) across the 35 million matched CpGs tested. Only 5.1% of classified reads exceeded the ctDMR-assignment methylation-difference threshold of 0.4 relative to sorted references, corresponding to the points outside the red dashed lines in **Figure 2a**. Disagreement was low across most PBMC compartments, including monocytes, T cells, NK cells and broader lymphocyte assignments, with B cells, which have the smallest read set, showing the highest disagreement rate at approximately 10.2-10.3% (**Supplementary Fig. 6**). In the brain samples, SniffCell deconvolved reads’ methylation profiles showed similar concordance with 158 million CpGs, with Pearson correlations of 0.928-0.947 for neuron and 0.937-0.962 for oligodendrocyte (**Figure 2b**). The mean absolute error (MAE) of DNA methylation between SniffCell deconvoluted reads and sorted reads was below 0.075 in all comparisons (≤0.056 in PBMC, ≤0.073 in brain) across 5.6-20.9 million shared CpGs per comparison (**Supplementary Fig. 5**). Overall, only 6.4% of classified reads exceeded the ctDMR-assignment methylation-difference threshold (**Figure 2b**), with similarly low disagreement across neuron and oligodendrocyte assignments. Together, these results demonstrate the accuracy of SniffCell read-level deconvolution.

**Figure 2:**
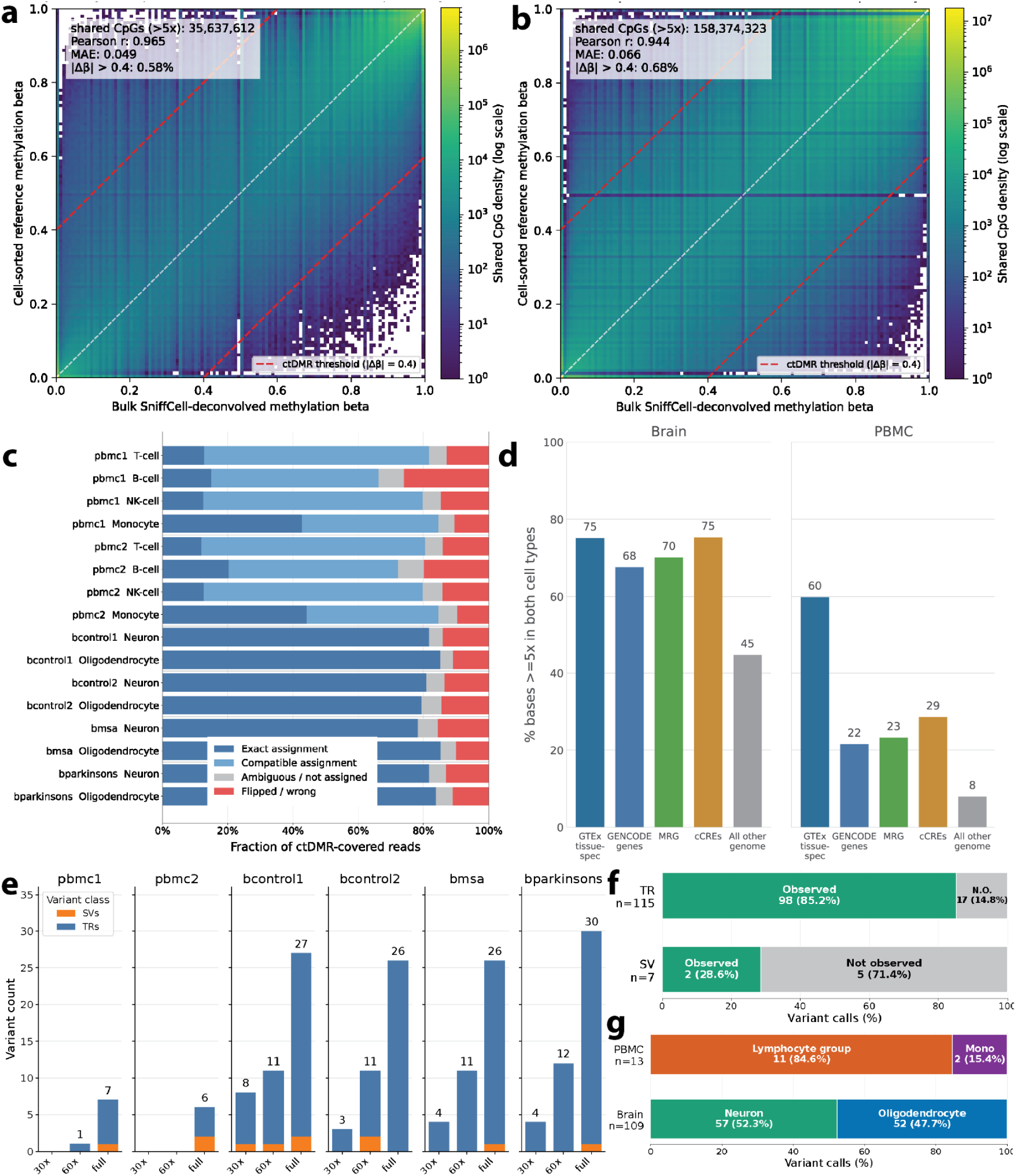
Genome-wide coverage and variant calling from SniffCell-classified reads. **a,** Agreement between DNA methylation values from SniffCell-deconvoluted reads and FACS-sorted reads at matched CpGs in PBMC samples. Dashed red lines indicate the ctDMR-definition threshold. **b,** Corresponding comparison between SniffCell-deconvoluted and FANS-sorted reads in brain samples**. c,** Read-assignment accuracy in FACS/FANS-sorted samples. **d,** Genomic coverage of SniffCell-deconvoluted reads**. e,** Variant counts across sequencing-depth settings**. f,** Recovery rate of variants detected at full coverage**. g,** Cell-type distribution of SniffCell-called cell-type-specific variants across PBMC and brain samples.

To benchmark read-level cell-type assignment, we used FACS/FANS validation tissue set for confirmation. SniffCell was applied independently to reads from each sorted fraction using the corresponding tissue atlas, and the fraction assigned to the expected cell type or compatible cell-type group was calculated as the read-assignment accuracy. Across eight brain samples, including neuron- and oligodendrocyte-enriched fractions from four donors, the mean read-assignment accuracy was 82.05%. Across eight PBMC samples representing T cells, B cells, NK cells, and monocytes from two donors, the mean compatible accuracy was 78.71%. Overall, the tissue-stratified accuracy across all 16 samples was 80.38%, demonstrating that SniffCell read assignments were strongly concordant with experimentally sorted cell identities (**Figure 2c**). Reads carrying conflicting DNA methylation signatures across the multiple ctDMRs they spanned (e.g., both neuron and oligodendrocyte DNA methylation signatures present on a single read), accounted for 6.0-7.5% of PBMC reads and 6.1-9.05% in brain. More than half of the conflicting brain reads had monocyte characteristics (**Supplementary Fig. 7)**. These may represent microglia, which are replenished in the aging brain by infiltrating monocytes from peripheral blood, which retain peripheral monocyte methylation profiles^19^. Principal component analysis of all deconvoluted reads from SniffCell confirmed high agreement with sorted reads (**Supplementary Fig. 8**). Because sorted fractions were generated independently from the bulk sequencing, agreement between SniffCell’s methylation-based read assignment and sorted-cell identity cannot be explained by coverage loss or noise introduced by splitting bulk reads. These independent validations therefore support a genuine cell-type-resolved signal rather than an artifact of the deconvolution procedure itself. In the bulk sequencing files, neuron- and oligodendrocyte-associated candidates were detected at near-equal proportions, despite oligodendrocytes representing the majority cell population in the sample (SniffCell resolved cell-type-specific read numbers: oligodendrocyte: 21,489,902 reads (53.53%) vs neuron: 18,653,355 reads (46.47%)) and correspondingly larger cell-type-assigned read pools.

Variant discovery additionally depends on whether cell-type-assigned reads provide sufficient coverage at the locus (**Figure 2d; Supplementary Fig. 9, Supplementary Table 3**). As expected for a methylation-guided approach, callable regions were enriched in tissue-relevant genes and regulatory elements rather than uniformly distributed across inactive genomic space. SniffCell produced ≥5x coverage over 60% of tissue-expressed genes in blood (720 genes, **methods**) or brain (4,727 genes, **methods, Supplementary Table 1**) with deconvoluted cell-type-specific reads (**Supplementary Fig. 10**). Furthermore we observed the density of ctDMR increases with the GTEx expression level of the tissue (**Supplementary Fig. 10C, D**). Thus, methylation-informative coverage is non-uniform and preferentially intersects genes expressed in the profiled tissue^14^. We next measured this callable space across general gene and regulatory annotations (**Supplementary Fig. 9C**). In PBMCs, SniffCell-deconvolved reads covered 28% of GENCODE^20^ genes, 22% of medically relevant genes^21^ and 28% of cCRE regions with ≥5x coverage. Additionally, 35 of 80 pathogenic TR loci curated in STRchive^22^ had sufficient cell-type-assigned coverage **(Supplementary Fig. 11)**. In the brain, where neuron- and oligodendrocyte-associated ctDMRs are denser across the genome, SniffCell covered 67% of GENCODE genes, 61% of medically relevant genes and 67% of cCRE regions. This included 67 of 80 pathogenic TR loci^22^ **(Supplementary Fig. 11)**. In total, SniffCell deconvoluted 31.2% of reads in the brain and 15.4% in PBMC, therefore providing sufficient coverage to enable downstream cell-type-aware variant discovery.

#### SniffCell recovers lineage-restricted somatic variation in blood

We focused the downstream analysis on somatic SNV, TR and SV candidates supported by cell-type-assigned reads rather than on germline variation itself (**Supplementary Table 4**). For SNVs, the effective sensitivity of bulk long-read calling is in the variant allele fraction range above 5%, which is expected to be enriched for earlier developmental events shared across tissues or cell types, while highly cell-type-restricted SNVs are likely rarer^23^. Consistent with this expectation, we did not identify high-confidence SNVs uniquely enriched in cell-types after FACS comparison and manual review (**Supplementary Fig. 12**). In contrast, SV and TR instability can arise recurrently at the same repeat locus across many cells post-mitotically within a susceptible cell-type, producing a detectable shift or expansion tail in the TR distribution^24^.

Across the two PBMC samples, we identified three lymphocyte-associated SVs and no monocyte-associated SVs (**Supplementary Figures 13-15, Supplementary Table 5**). Two out of three SVs occurred within antigen receptor variable-diversity-joining (V(D)J) rearrangements, which generate programmed somatic deletions at T cells receptor (TCR) loci in T cells and immunoglobulin loci in B cells, providing events of known lineage specificity^25^. SniffCell identified cell-type-specific deletion signatures at the canonical TRB, TRA/TRD, TRG, IGH, IGK and IGL loci (**Figure 3a, Supplementary Table 6**), with TCR-locus deletions assigned predominantly to T cells and immunoglobulin-locus deletions assigned predominantly to B cells. In total, SniffCell identified 13 read-level T cells-associated and 3 B cells-associated V(D)J events from bulk ONT sequencing. At the TRB locus, deletion-supporting reads were consistently enriched in the T cells-assigned fraction, and this assignment was independently confirmed by sorted T cells, which showed clear breakpoint-spanning support for the deletion, while sorted NK cells and monocytes did not (**Figure 3a, b; Methods**).

**Figure 3:**
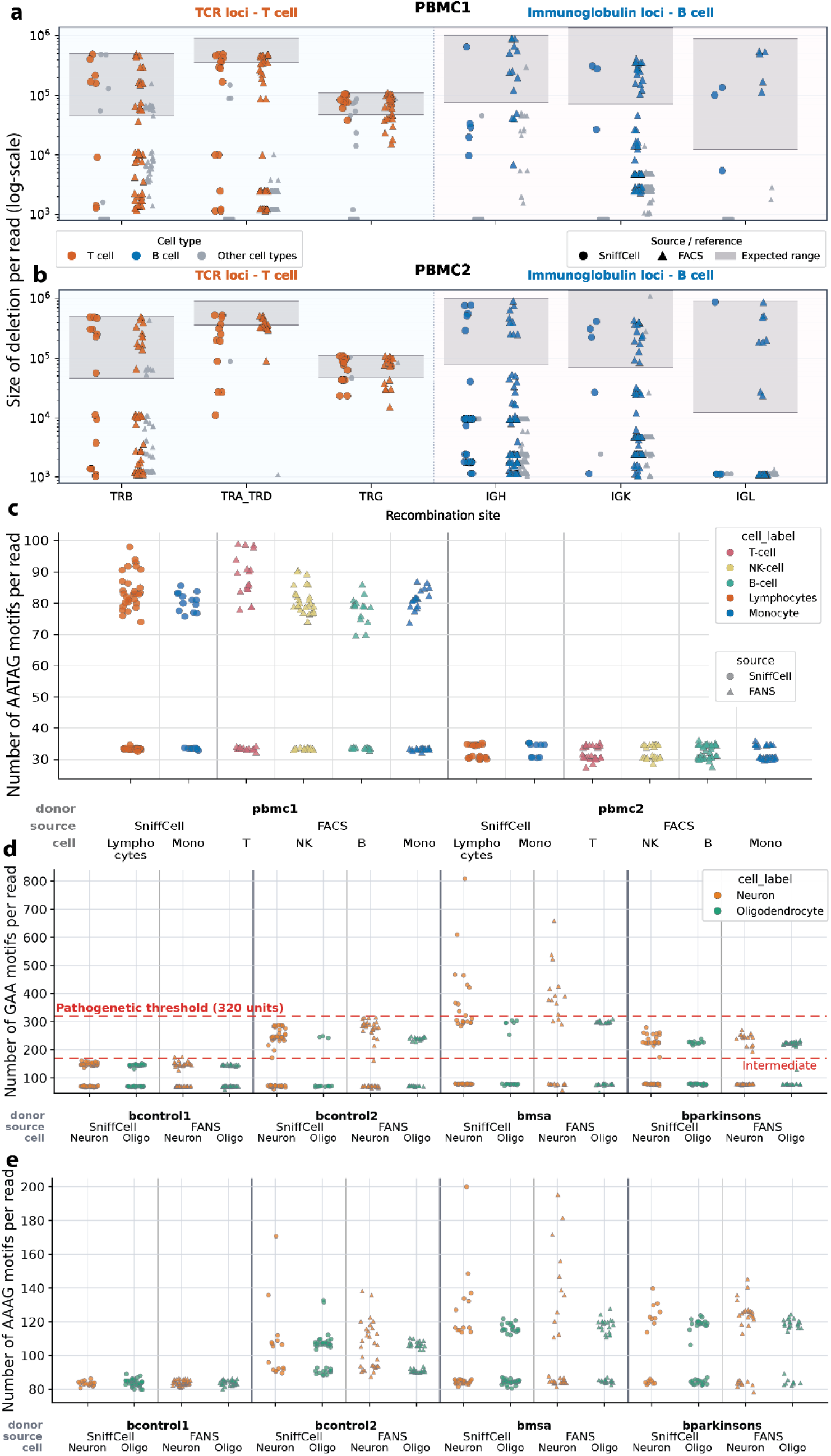
SniffCell detected cell-type-specific variants. **a.** SniffCell-clustered reads at six blood somatic recombination sites in pbmc1. Boxes indicate the expected deletion lengths derived from the distance between the genes in GRCh38. **b.** SniffCell-clustered reads at six blood somatic recombination sites in pbmc2. **c.** T-cell-associated somatic expansion at the CALCOCO2 repeat. **d.** Somatic neuronal FGF14 expansion across four samples. The germline expansion in bmsa is borderline pathogenic, but neuronal alleles extend beyond the reported pathogenic threshold^22^. **e.** Recurrent neuron-associated expansion of a GAAA repeat in LRRC7 in three of four FANS validation brains.

**Figure 3c** shows that SniffCell highlights an interesting somatic TR expansion specific to lymphocytes in the PBMC1 bulk data. The SniffCell-annotated lymphocyte reads carried repeat alleles up to 490 bp while the monocyte reads remained below 450 bp. The AATAG repeat in *CALCOCO2* was previously reported to undergo somatic expansion in blood^26^, and here we show that the expansion occurs predominantly in lymphocytes. Validation using sorted fractions showed TR expansion only in T cells. Together, the PBMC examples demonstrated that SniffCell correctly recovered known lineage-restricted somatic rearrangements directly from bulk long-read data and cell-type-specific tandem repeat expansions. The concordance between methylation-informed bulk assignments and sorted-cell validation supports the use of SniffCell to resolve the cellular origin of somatic variants beyond antigen receptor loci, where the relevant cell-type is not known a priori. When comparing the cell-type-specific variants, we observed an enrichment of variants in certain cell-types. Across the PBMC samples, SniffCell identified 12 cell-type-associated TR expansions, including 10 lymphocyte-associated (T, B, and NK cells) and 2 monocyte-associated events (**Figure 2e, Supplementary Table 5**). In PBMCs, the larger number of lymphocyte-associated candidates is consistent with the clonal expansion and programmed DNA remodelling of adaptive immune cells^27,28^. This distribution is consistent with the expected composition of PBMCs, where lymphocytes typically represent approximately 70-90% of cells and monocytes approximately 10-20%^29^. We therefore interpret the PBMC candidate distribution as broadly concordant with expected cell composition and callable cell-type-assigned read space, rather than as evidence for an increased intrinsic mutation rate in lymphocytes.

Somatic variant detection is inherently coverage-dependent, because lower allele fractions require sufficient sequencing depth to observe them^5^. For SniffCell, this general constraint is compounded by the need for the same reads to carry both variant evidence and informative ctDMRs for cell-type assignment. To estimate the coverage required for this combined genetic and epigenetic readout, we downsampled the four-flow-cell ONT datasets (∼120x) to approximate two-flow-cell (∼60x) and one-flow-cell (∼30x) sequencing depths and quantified recovery of SniffCell-called SV and TR candidates. Figure 2e shows variant counts across sequencing depths. We didn’t recover the somatic SVs in the 60x and 30x PBMC subsamples, owing to reduced coverage and the lower density of informative ctDMRs in the four blood cell-types. Only one TR candidate was recovered in the 60x subsamples.

##### SniffCell reveals recurrent neuron-associated tandem-repeat expansions

We next applied SniffCell to four frontal cortex samples with matched sorted neuronal and oligodendrocyte fractions to determine whether bulk long-read methylomes could resolve somatic variation in major brain cell-types. Bulk germline variant profiles were comparable to those observed in PBMCs, with 3,805,804-4,036,555 SNVs and indels, 24,275-24,456 SVs and 613,802-616,454 genotyped TR loci per sample (**Supplementary Table 4)**. SniffCell resolved 5 cell-type-specific SVs across the four donors. These included 5 neuron-associated SVs (**Supplementary Fig. 16-20, Supplementary Table 5**), with available FANS data supporting the cell-type assignment for manually reviewed events. For tandem repeats, SniffCell identified 53 neuron-specific TR events and 52 oligodendrocyte-specific TR events.

In the brain samples, SniffCell identified 51% of all cell type specific TRs as neuron-associated and 49% as oligodendrocyte-associated. This near-parity, rather than an oligodendrocyte-skewed distribution proportional to cell abundance, suggests that detection here is not simply tracking coverage or cell representation. Comparison with paired FANS calls and manual review validated the cell-type-specific signal for 83 of 92 unique brain TR loci in at least one paired FANS comparison (**Figure 2f, Supplementary Table 5**). The nine loci not observed in paired FANS data may reflect differences in cellular sampling between bulk and FANS-sorted brain tissues, insufficient coverage of one haplotype among SniffCell-deconvoluted reads or the selection of partially aligned reads in TR regions.

Several neuron-associated expansions recurred at the same loci across donors and were enriched for GAA-rich motifs. This sequence bias is notable given previous evidence that repeat composition influences somatic instability through replication-independent DNA repair mechanisms^30,31^. FANS-validated neuron-associated expansions were significantly enriched for manually annotated GAA-rich motifs compared with oligodendrocyte-associated expansions (43.9% of neuron-associated versus 16.3% of oligodendrocyte-associated loci, Fisher’s exact P = 0.0083, **Figure 2f**). In contrast, oligodendrocyte-associated TR expansions were less often recurrent across donors. At many of these loci, the matched blood comparison suggested a germline-expanded allele, with additional cell-type differences in repeat length: neurons could also carry expanded alleles, but these were typically longer than those observed in oligodendrocytes. For example, a TR expansion in a parkinsonism-related gene *SGIP1*^32^ (chr1:66630776-66631127) with a GAAA motif was detected in both neurons and oligodendrocytes, but neuronal reads showed a longer tail than oligodendrocyte reads in bcontrol2 (median Δ = +34.5 bp; 11/48 neuronal reads exceeded the oligodendrocyte maximum) and bmsa (median Δ = +11 bp; 5/79 exceeded the oligodendrocyte maximum), indicating support from multiple reads rather than a single outlier (**Supplementary Fig. 21**).

**Extended Data Figure 1.**
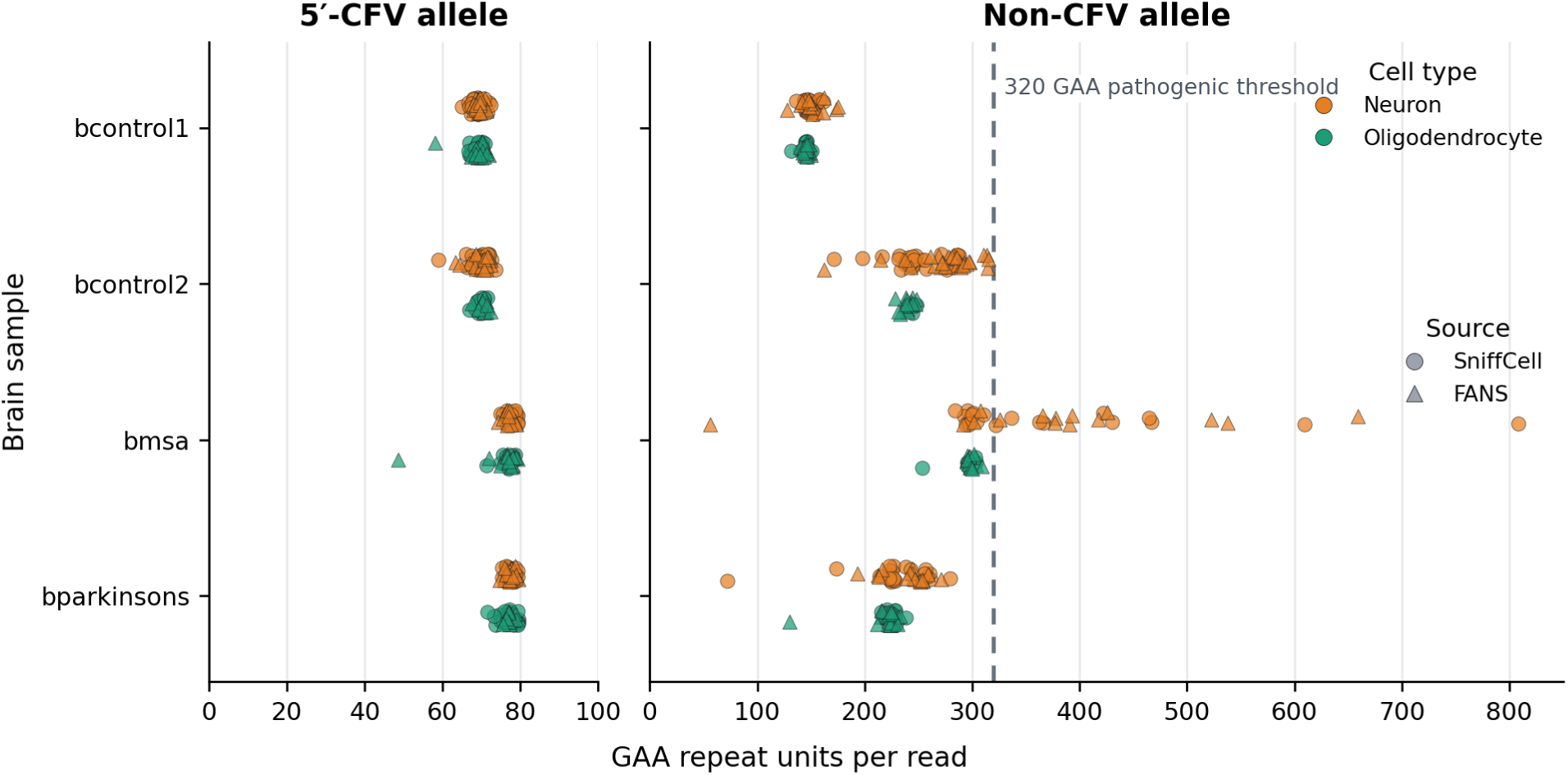
| The stability-associated FGF14 5′-CFV was confined to the shorter, non-expanded allele, whereas expanded alleles lacked the 5′-CFV. Circles represent reads assigned to CFV-containing alleles, and triangles represent reads assigned to non-CFV alleles. The x-axis shows the read-level GAA repeat-unit count; neuron and oligodendrocyte reads are distinguished by color.

We further analyzed the recurrent TR expansion events in neurons. All six recurrent neuron expansion events overlapped with gene bodies. These included loci within genes with neuronal or synaptic functions, including *SH3RF3, SHANK1, PLXNA4, LRRC7* and *FGF14*. *FGF14* is a pathogenic tandem repeat locus and a common cause of autosomal dominant spinocerebellar ataxia^33–35^ with repeat sizes ≥320 classified as pathogenic and 180–319 as intermediate with partial penetrance. Expansions at this locus have also been reported in association with MSA, including faster disease progression in carriers^36^, and with PD^37^. Three nearby ctDMRs enabled methylation-based assignment of expansion-supporting reads, which were enriched for a neuronal signature in bulk tissue and independently confirmed in matched FANS-sorted neuronal DNA. Neuron-restricted somatic expansion of *FGF14* was observed on the larger allele in three of the four samples, including one control (bcontrol2), the MSA sample (bmsa) and the PD sample (bparkinsons), but was not detected in bcontrol1 with the lowest germline tandem repeat size (**Figure 3d**). Interestingly, in the MSA sample, neuronal expansion further extended the germline-expanded *FGF14* allele into the range reported as pathogenic^22^, with the longest read having 500 repeats above the reported pathogenic threshold. The somatic expansions in the PD and control samples, where the germline alleles were intermediate in size (226 and 245 respectively), were more modest and did not reach the full pathogenic threshold. At the consensus level, we detected two AGG interruptions in bcontrol1 and two GAAA interruptions in bcontrol2 and bparkinsons. These interruptions did not differ between cell types, whereas the somatically expanded sequence consisted only of uninterrupted GAA motifs. This result links the somatically expanded repeat molecules to neurons and distinguishes the inherited repeat expansion from an additional neuron-associated expansion at the same locus. As an independent test of this allele-level assignment, we examined a previously described 5′ common flanking variant (5′-CFV) at *FGF14* that is known from germline genetic studies to segregate exclusively with the non-expanded allele^33^. Using SniffCell-deconvolved and sorted reads, we found the 5′-CFV present only on the shorter allele in every sample, with stable read lengths (694-738 bp) in both neurons and oligodendrocytes, whereas the alleles lacking the flanking variant showed pronounced neuronal instability, extending to substantially longer repeat lengths in neurons than in oligodendrocytes from the same donors (**Extended Data Figure 1**). This cell-type-autonomous relationship, recovered independently at the single-molecule level, supports the conclusion that the neuron-specific expansion signal reflects genuine somatic instability.

**Extended Data Figure 2.**
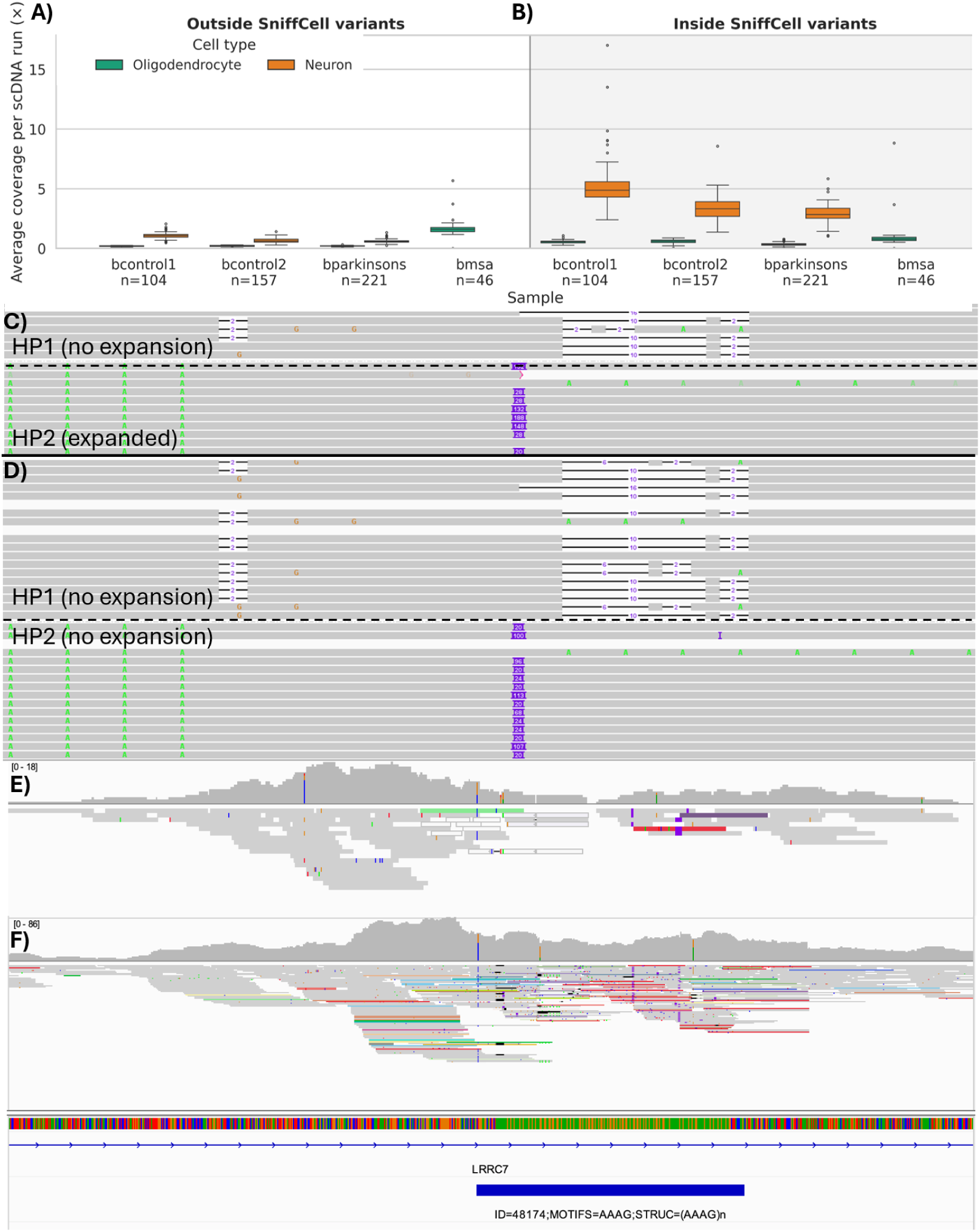
| Paired single-cell Illumina DNA sequencing of four brain samples. **A)** Average coverage per scDNA run for neurons and non-neuronal cells/oligodendrocytes genome-wide. Higher coverage indicates the repeat expansion inside SniffCell-called variant intervals. Boxes show the median and interquartile range, whiskers extend to 1.5 times the interquartile range, and points indicate outliers. **B)** Total coverage, calculated as the sum of per-run mean coverage, inside of SniffCell-called variant intervals for each sample and cell type. Numbers below sample names indicate the number of scDNA runs. Green denotes oligodendrocytes (for MSA) and non-neuronal cells for the other samples, and orange denotes neurons; the bmsa sample contains oligodendrocytes only. **C)** Bcontrol2 SniffCell-classified neuron read carrying the repeat expansion at HP2, reflected by the size of the insertion. **D)** Bcontrol2 SniffCell-classified oligodendrocyte read not carrying the repeat expansion. **E)** Bcontrol2 Oligodendrocyte scDNA read alignments at the LRRC7 repeat. Colored reads indicate discordant mate alignments and local alignment disruption consistent with an unresolved expansion. **F)** Bcontrol2 Neuronal scDNA read alignments at the same locus.

A second example was a highly polymorphic tetranucleotide repeat within an intron of *LRRC7* (chr1:69781654-69782004, *GAAA*, **Figure 3e**). *LRRC7* encodes a brain-enriched postsynaptic scaffold that links cytoskeletal and signalling proteins at glutamatergic synapses^38^. SniffCell assigned the longest alleles preferentially to neurons, and this neuron-associated expansion pattern was further validated by sorted neuronal fractions. These examples show that neuron-associated TR expansions can recur across independent brains^24^. Given the high recurrence of the *LRRC7* repeat expansion across three samples, we next examined whether matched scWGS could recover the recurrent *LRRC7* expansion. In the paired bcontrol2 sample, despite profiling 93 neuronal and 64 non-neuronal single-cell genomes, local coverage averaged only 1.23x and 0.13x (**Extended Data Figure 2A**), respectively. The single-cell Illumina data provided orthogonal support for a repeat abnormality at the SniffCell-identified locus (**Extended Data Figure 2B**), although they could not resolve the expansion itself. The expanded repeat produced a local pile-up of reads together with disrupted alignments, but the short-read data lacked sufficient repeat-spanning information for accurate genotyping (**Extended Data Figure 2C, D**). Together, these results show that bulk native long reads can resolve cell-type-associated complex variation that could not be directly resolved in these matched single-cell short-read data.

Finally, we evaluated whether SniffCell could detect cell-type-enriched somatic SNVs using matched scWGS as orthogonal evidence. Detection of low-allele-fraction somatic variants remains challenging in ONT data because true variants must be distinguished from residual, often sequence-context-dependent sequencing errors^39^. We identified 356 recurrent apparent neuron-enriched SNVs that were selected from FANS neuronal ONT data but were not confirmed as neuron-restricted by scWGS. These sites showed a pronounced T>G/A>C substitution bias, accounting for 73.0% of candidates, and 81.5% were observed in both FANS-sorted and SniffCell-classified neuronal reads from at least two of the four benchmark donors. However, the exact variants were observed less frequently in oligodendrocyte ONT and scWGS datasets, supporting their interpretation as reproducible sequence-context-dependent artifacts in neurons (**Supplementary Fig. 12; Supplementary Table 7**).

Across the benchmark datasets, TRs provided by far the strongest and most reproducible cell-type-associated somatic signal, whereas robust detection of cell-type-restricted SNVs and non-repeat SVs remained limited. This difference reflects both the sensitivity constraints of bulk sequencing and the distinct biology of repeat instability. A cell-type-restricted SNV or SV must reach sufficient cellular prevalence to generate multiple supporting molecules, whereas unstable repeat loci can undergo recurrent length changes within a susceptible cell population, producing a detectable shift in the distribution of repeat lengths. We therefore focused subsequent analyses on TRs, which provided the strongest and most reproducible cell-type-associated somatic signal in these datasets.

#### Cell type, repeat motif and brain region shape somatic TR instability

The four validation brain samples with matched sorting data revealed different patterns of neuron-specific and oligodendrocyte-specific TR expansion. Previous studies suggest that neuronal repeat expansions can show locus- and region-specific patterns in the brain^40,41^. Differences between *FGF14 GAA*-repeat and *HTT CAG*-repeat somatic expansion patterns may reflect repeat motif-specific mechanisms, regional neuronal vulnerability or expansion in distinct neuronal subtypes. To test whether neuron-associated repeat expansions extend beyond the frontal cortex, we analyzed 86 brain sequencing runs (SMaHT multi-region donor cohort) from the SMaHT^42^ consortium paired with matched long-read whole-blood genomes from the same 13 donors (data.smaht.org). These included ONT and PacBio data from frontal lobe, temporal lobe, cerebellum and hippocampus. Matched blood from the same donor was used as a germline repeat-length baseline.

Using SniffCell, we first investigated neuron-specific expansions across these samples to identify recurrent neuron and oligodendrocyte expansion events. We initially identified 56 neuron-associated and 74 oligodendrocyte-associated TR expansion events. Because some loci showed opposite expansion directions across different samples (i.e., neuronal alleles were longer in some samples, whereas oligodendrocyte alleles were longer in others), we further restricted the analysis to loci consistently associated with only one cell-type. This resulted in 25 neuron-specific and 43 oligodendrocyte-specific expansion loci (**Figure 4a**). We examined the subset of cell-type-specific expansion loci that recurred in at least two independent donors. Ten (40%) of the 25 neuron-specific loci and 17 (39.5%) of the 43 oligodendrocyte-specific loci met this recurrence criterion (**Supplementary Table 8**). Recurrent neuron-associated loci also showed a distinct motif composition. AAG-like motifs occurred at 4 of 10 recurrent neuron-specific loci but only 1 of 17 recurrent oligodendrocyte-specific loci (Fisher’s exact P = 0.047). Thus, recurrent neuron-specific expansions showed a pronounced *AAG*-like sequence pattern, whereas recurrent oligodendrocyte-specific expansions exhibited a more heterogeneous motif composition (**Supplementary Table 9**).

**Figure 4:**
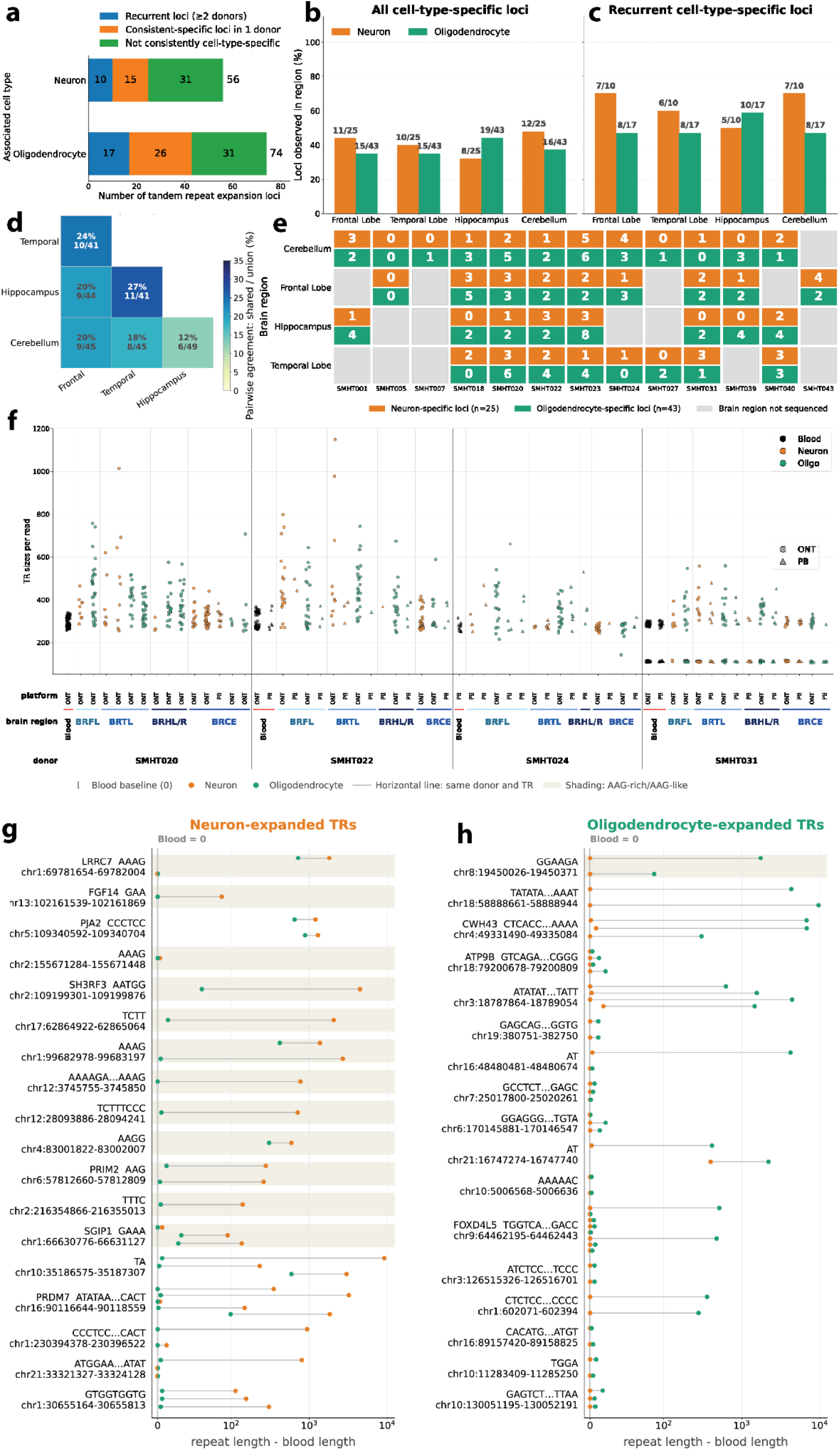
**a.** numbers and proportions of total SniffCell-called somatic TR expansions, single-donor-consistent cell-type-specific loci, cross-donor recurrent cell-type-specific loci. **b.** Percentage of all cell-type-specific TR expansion loci observed in each brain region, stratified by neuron-specific and oligodendrocyte-specific loci. Bars show the fraction of loci detected in a given region among all loci in that cell-type-specific category. Neuron-specific loci are shown in orange and oligodendrocyte-specific loci are shown in green. **c.** Percentage of recurrent cell-type-specific TR expansion loci observed in each brain region**. d.** Pairwise overlap of all cell-type-specific TR expansion loci across brain regions. Each heatmap cell shows the percentage of shared TR loci between two regions, calculated as the number of shared loci divided by the union of loci observed in either region. The cerebellum shows the weakest sharing with other regions. **e.** Donor- and region-level distribution of cell-type-specific TR expansion loci. Each tile represents one donor-brain-region sample. The upper half of each tile shows the number of neuron-specific TR expansion loci detected in that sample, and the lower half shows the number of oligodendrocyte-specific TR expansion loci. Numbers indicate unique globally cell-type-specific TR loci detected in each donor–region sample. **f.** Read-level tandem-repeat sizing at the PJA2 locus (chr5:109340592-109340704, motif CCCTCC) across six expanded SMaHT donors. Each point represents one read, with TR size shown on the y-axis. Samples are grouped by donor and tissue, ordered as blood, frontal lobe (BRFL), temporal lobe (BRTL), hippocampus (left and right, BRHL/R), and cerebellum (BRCE). Colors denote cell-type or tissue class: blood, neurons, and oligodendrocytes. Marker shape denotes sequencing/source category. **g-h.** Cell-type-specific tandem-repeat expansion relative to matched blood. Donor-normalized repeat-length differences are shown for 88 donor–locus pairs across 35 tandem-repeat loci classified by SniffCell as neuron-expanded **g.** or oligodendrocyte-expanded **h.**. Each horizontal line connects neuron (orange) and oligodendrocyte (green) measurements from the same donor and locus. Values represent the median cell-type repeat length minus the longest matched-blood allele; values at or below blood are displayed at zero and are not interpreted as contractions. Exact overlaps are shown as split orange–green circles. Multiple donors at the same locus occupy separate vertical positions. The gray vertical line marks the matched-blood baseline, and shaded rows denote AAG-rich or AAG-like repeats. The x-axis uses a symmetric logarithmic scale.

We next examined the regional distributions of cell-type-specific TR expansions. Among the 25 neuron-specific loci, 12 were detected in the cerebellum, 11 in the frontal lobe, 10 in the temporal lobe, and 8 in the hippocampus. After accounting for the number of sampled donors, neuron-specific expansions were descriptively most prevalent in the frontal lobe and least prevalent in the hippocampus (**Figure 4b**). A one-sided donor-stratified permutation test comparing the mean prevalence in the frontal lobe and cerebellum with that in the hippocampus was nominally significant (P = 0.039), although this association did not remain significant after multiple-testing correction (FDR = 0.110). Among the 43 oligodendrocyte-specific loci, 19 were observed in the hippocampus, 16 in the cerebellum, and 15 each in the frontal and temporal lobes. Oligodendrocyte-specific expansions were descriptively most prevalent in the hippocampus, although the corresponding one-sided permutation test did not reach significance (P = 0.062; FDR = 0.110, **Figure 4b**).

A similar descriptive pattern was observed among recurrent expansions. Seven of the 10 recurrent neuron-specific loci were detected in each of the frontal lobe and cerebellum, compared with six in the temporal lobe and five in the hippocampus (**Figure 4c**). Conversely, 10 of the 17 recurrent oligodendrocyte-specific loci were detected in the hippocampus, whereas eight were detected in each of the other regions. These findings suggest contrasting regional tendencies, with neuron-specific expansions occurring more frequently in the frontal lobe and cerebellum and oligodendrocyte-specific expansions occurring more frequently in the hippocampus. However, the global permutation tests did not establish significant regional heterogeneity, and larger cohorts will be required to validate these patterns.

The cerebellum consistently shared the fewest cell-type-specific TR loci with other brain regions, in both neuron- and oligodendrocyte-specific categories, while the temporal lobe and hippocampus showed the highest pairwise overlap (**Figure 4d**). This pattern was retained after stratification by cell type (**Supplementary Fig. 22**). This regional distinctiveness may reflect the cerebellum’s largely homogeneous granule-cell population, in contrast to the diverse excitatory and inhibitory neuronal populations of the frontal and temporal cortices. **Figure 4e** shows the corresponding donor- and region-level distribution of cell-type-specific TR expansion loci.

Although cerebellum samples consistently carried expanded alleles, their repeat lengths were generally shorter than those observed in the frontal and temporal lobes. The most striking example is the *PJA2* repeat expansion. The recurrent *PJA2* expansion (chr5:109,340,592–109,340,704; CCCTCC/GGAGGG) was previously detected in four sorted validation brain samples, and we observed the same expansion in 6 of the 13 SMaHT multi-region donors. In neural systems, *PJA2*-mediated ubiquitination promotes neurite and axonal growth through the proteasomal degradation of NOGO-A and regulates cAMP-PKA signaling involved in synaptic plasticity and long-term memory, making recurrent variation at this locus particularly notable, although the functional consequences of this repeat expansion remain unknown^43,44^. As shown in **Figure 4f**, the *PJA2* repeat was expanded to broadly similar lengths in neurons and oligodendrocytes, although the neuronal alleles were slightly longer on average. Repeat size also differed significantly among brain regions, with the shortest alleles observed in the cerebellum. These regional differences are consistent with different neuronal subtypes carrying distinct somatic repeat-length distributions.

We next asked whether cell-type-specific expansions exceeded the donor’s inherited repeat length. Across the 10 donors with recurrent TR instabilities in both neurons and oligodendrocytes, the median donor-level fraction of events with a concordant expanded allele in matched blood from the same donor was 29.2% for neuron-associated events and 66.7% for oligodendrocyte-associated events (paired Wilcoxon P=0.0156). These results indicate that neuron-associated expansions more frequently diverged from the inherited repeat-length baseline than oligodendrocyte-associated expansions. In contrast, oligodendrocyte-specific TR expansions more closely tracked the inherited repeat-length baseline observed in matched blood, consistent with less additional oligodendrocyte-specific expansion beyond the inherited repeat-length baseline. It is striking that neuron-associated expansions showed substantially greater repeat-length divergence, especially at *GAA*-rich tandem repeats. **Figures 4g** and **4h** compare cell-type-specific repeat lengths with the longest matched-blood allele. We used the median length of all the expanded reads from the donor regardless of the brain region to represent the repeat expansion size in the figure. GAA-rich TR expansions were predominantly neuron-specific, with a single exception (chr8:19450026–19450371; GGAAGA) that was oligodendrocyte-associated. The recurrent signal was driven primarily by additional repeat expansion in neurons, rather than by a shared increase across all brain-derived cell populations. Thus, blood sequencing alone substantially underestimates the repeat-length distributions present in brain cell populations.

Together, the matched SMaHT multi-region donor cohort highlights that somatic TR instability in the brain is shaped by cell identity, repeat motif and anatomical region. Neuron-associated expansions were enriched for GAA-rich motifs and diverged most strongly from matched blood, whereas oligodendrocyte-associated expansions were more heterogeneous and more often tracked the donor’s blood allele length. These results indicate that bulk brain repeat profiles reflect a mixture of inherited repeat length, regional cellular composition and additional somatic expansion within specific cell populations.

##### Neuron-associated repeat expansions recur across independent brain cohorts

To replicate the somatic tandem-repeat expansion patterns observed in the multi-region donor brains, we applied SniffCell to frontal cortex samples from 155 donors. Although the CARD genomes were sequenced at approximately 37x, their substantially longer read lengths increased (**Supplementary Fig. 23**) the number of molecules spanning both tandem repeats and nearby informative ctDMRs, providing greater effective coverage for cell-type-aware analysis than nominal depth alone would suggest. Across the 155 CARD brain donors from the NIH CARD consortium^45^, SniffCell identified 2,268 neuron-specific and 3,707 oligodendrocyte-specific cell-type-differential TR loci. Of these, 157 neuron-specific and 311 oligodendrocyte-specific TRs were recurrent in at least three donors (**Supplementary Table 10, 11**). Motivated by the enrichment of GAA-rich motifs among neuron-specific expansions in the multi-region donor brains, we next examined whether recurrent CARD brain expansions showed a similar motif bias. Pure GAA repeats showed the clearest neuron enrichment among loci recurrent in at least three donors, with three neuron loci and no oligodendrocyte loci. Pure GAAA repeats showed a weaker difference (i.e., four neuron versus two oligodendrocyte loci) whereas pure GGAA repeats showed no neuron enrichment, with six neuron versus nine oligodendrocyte loci. A permutation test supported enrichment of pure GAA repeats among neuron-associated loci recurring in at least three donors (p=0.0061 for loci recurrent in >=3 donors, **Supplementary Fig. 24**). More broadly, among short-motif loci recurrent in at least three donors, GAA-rich motifs accounted for 22 of 73 neuron-skewed loci compared with 12 of 74 oligodendrocyte-skewed loci (**Figure 5a**). These results suggest that the neuron-associated signal is motif-context dependent rather than a general property of all GAA-rich repeats. One notable example was a GAATG repeat within *SH3RF3*. *SH3RF3* has previously been implicated in protection against Alzheimer’s disease through genetic and functional evidence^46^.

**Figure 5:**
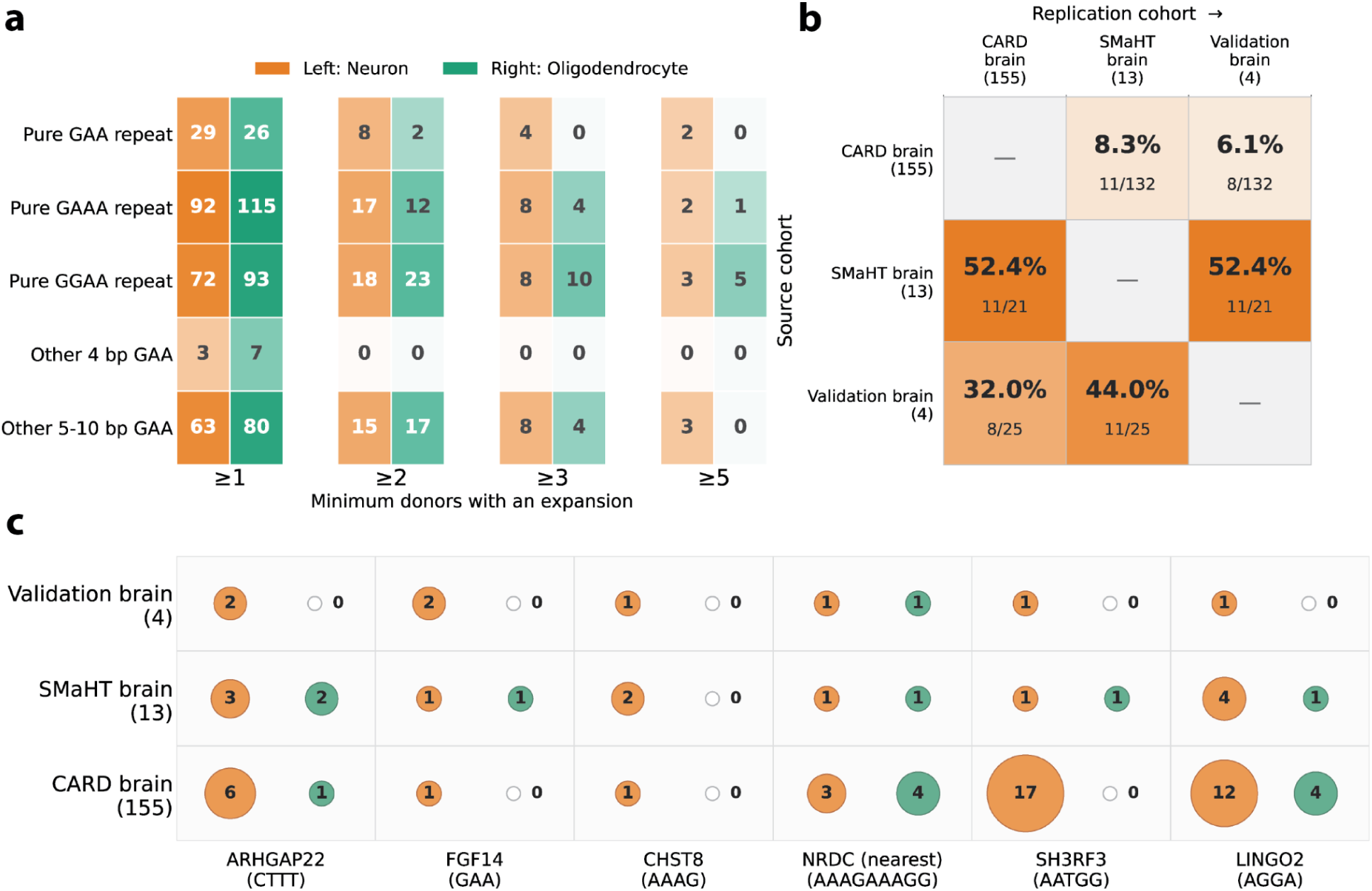
Neuron-associated tandem-repeat expansion patterns in 155 CARD frontal cortex cohort. **a.** Number of CARD loci with expansions in at least 1, 2, 3, or 5 donors, stratified by GAA-element motif family. Each box is divided between neuron (left, orange) and oligodendrocyte (right, green); darker shading indicates more loci. Loci may be counted in both cell types. **b.** Directional replication of GAA-element neuron expansion loci among the CARD brain cohort (155 donors), SMaHT brain cohort (13 donors), and validation brain cohort (4 benchmark brains). Rows indicate the source cohort and columns indicate the replication cohort. Each off-diagonal cell shows the replication percentage, followed by the number of exactly coordinate-matched loci divided by the total number of loci in the source cohort. **c.** Six GAA-element neuron expansion loci detected at identical GRCh38 coordinates in all three cohorts. For each locus and cohort, the left orange bubble represents neuron expansions and the right green bubble represents oligodendrocyte expansions. Bubble size increases with donor count, and the exact count is printed within each bubble (hollow means 0). Genes and repeat motifs are shown below the matrix.

We detected neuron-specific expansion at this locus in 17 CARD brain donors and no oligodendrocyte-specific calls, and noted an increase in neuronal expansion size with age (Spearman ρ = 0.36, P = 0.155, **Supplementary Fig. 25A**). Similarly, with the *LINGO2* repeat expansion re-occurring in 12 donors from the CARD cohort, we also observe a positive correlation with age (Spearman ρ = 0.16, P = 0.618, **Supplementary Fig. 25B**). The *SH3RF3* and *LINGO2* expansions were independently identified as a recurrent brain expansion across three SMaHT multi-region donors and two benchmarking donors (**Figure 4g)**.

Finally, we examined cross-cohort overlap among the large-scale CARD brain cohort, the multi-region donor cohort and the FANS validation brains. **Figure 5b** shows that GAA-rich element neuron expansion loci replicated across cohorts at appreciable rates despite the large differences in cohort size. Of the 132 CARD brain GAA-element neuron loci, 11 (8.3%) were recovered in SMaHT multi-region donors and 8 (6.1%) in the four-donor FANS validation brain cohort. Conversely, 11 of 21 SMaHT donor loci (52.4%) were detected in CARD brains and the same number were present in the benchmark brains; among the 25 FANS-sorted brain loci, 8 (32.0%) replicated in CARD brains and 11 (44.0%) replicated in SMaHT multi-region donors. Six GAA-element loci were observed in all three cohorts (**Figure 5c**), five of which occur in genes with reported roles in brain biology or neurological disease, demonstrating recurrence at identical genomic sites across independently generated datasets. These included *SH3RF3* gene and *FGF14*, whose functions have been mentioned above. *LINGO2* has been associated with autism^47^, *CHST8* marks von Economo neurons^48^, and *ARHGAP22* shows high expression in the developing central nervous system^49^. Together, these analyses show that neuron-associated TR expansion is not simply a property of individual donors, but recurs across independent cohorts at a defined subset of repeat loci shaped by sequence context and cellular identity.

## Discussion

SniffCell turns native bulk long-read WGS into a cell-type-aware mutation discovery experiment. The approach requires no change to the sequencing protocol and instead uses cell-type-specific DNA methylation as an endogenous barcode carried by each native DNA molecule. The same read therefore reports both the genetic variant and evidence about the cellular compartment from which it originated. This extends previous long-read methylation approaches for tissue-of-origin inference and tumor-normal read classification by assigning complex variant-supporting molecules to multiple defined cell populations using a reference cell-type methylation atlas^18^. Where the methylation evidence is sufficiently informative, SniffCell assigns variant-supporting reads to a cell-type; where a unique assignment is not possible, it can still exclude incompatible cell populations or retain the event as unresolved rather than forcing an unsupported classification. In matched PBMC and brain samples, SniffCell-assigned reads closely reproduced the methylation profiles of independently sorted populations, while antigen receptor rearrangements were assigned to their expected lymphocyte lineages, providing biological validation beyond methylation concordance alone. SniffCell further reports the sequence and methylation evidence in an interactive report, enabling users to inspect the variant, its supporting reads and the basis of the cellular assignment. Cell-type annotations are also written to a standards-compliant VCF, allowing the results to enter established variant filtering, annotation and interpretation workflows. This physical linkage is particularly important for TRs and complex SVs, where long molecules are needed to span the repeat or reconstruct the breakpoint. By comparison, short-read scWGS commonly provides sparse and uneven coverage and remains affected by allelic dropout, amplification imbalance and limited molecule length^50^. SniffCell’s sensitivity depends on both sequencing depth and ctDMR density. Brain samples retained detectable cell-type-associated TR signals after downsampling, whereas PBMC analyses required greater depth because informative methylation markers were sparser across the relevant immune-cell classes. A single routine long-read genome can therefore provide cell-type-aware resolution of complex variation without generating and sequencing hundreds or thousands of separately amplified single-cell libraries, which would entail substantially greater experimental cost^51^. SniffCell makes cell-type-specific mutation detection an analytical extension of routine long-read WGS rather than a separate experimental design and enables the growing collection of existing native long-read genomes to be revisited for somatic variation that was previously inaccessible at cell-type resolution.

Cellular origin is essential for understanding somatic mosaicism and its potential impact because a mutation present in neurons may have fundamentally different consequences from the same event in oligodendrocytes, immune cells or another tissue compartment. This distinction is particularly important in neurological disease, where most genetic studies rely on blood and even brain sequencing is generally performed in bulk, although somatic mutations may have a role in neurodevelopmental and neurodegenerative diseases^52,53^. Blood can define the inherited allele, but it cannot fully represent the somatic allele distribution in the cells affected by disease. Bulk brain sequencing retains the relevant molecules but averages their signal across cell populations, obscuring whether an expansion is broadly distributed or concentrated within neurons. Previous locus-focused studies, most prominently at HTT, have shown that large somatic expansions can accumulate selectively in vulnerable neuronal populations and directly contribute to neurodegeneration^24^. SniffCell extends this principle from an individual pathogenic repeat to a genome-wide analysis of TR instability. Neuron-associated expansions recurred at specific loci across donors and brain regions and showed a preference for sequence motifs, indicating that neuronal repeat instability is structured rather than random. In contrast, oligodendrocyte-associated repeat lengths more frequently tracked the donor’s blood allele, whereas neuronal alleles showed additional expansion beyond this inherited baseline. This pattern is unlikely to reflect a simple coverage or cell-abundance artifact. Oligodendrocytes are the more abundant cell population in cortical tissue and are represented by correspondingly higher read counts in SniffCell-deconvolved bins (53.53% vs 46.47% in four brain benchmarking samples), which should, if anything, favor more sensitive detection of somatic variation in oligodendrocytes. The observation that neuron-assigned reads instead showed larger expansions and greater divergence from matched blood at recurrent neuron-associated loci despite this lower relative representation argues against a detection-power explanation and instead points to a genuine, cell-intrinsic difference in repeat instability between these two populations. These findings suggest that inherited repeat length and sequence establish the substrate for instability, but that the final allele distribution is reshaped within neurons. Blood-based analysis may therefore identify the inherited expansion while substantially underestimating the repeat lengths present in disease-relevant cells.

In the present datasets, TRs provided substantially greater cell-type-associated variability than SNVs or non-repeat SVs and therefore offered the strongest opportunity to examine somatic variation across individuals and tissues. The broader advance is the ability to identify cell-type-associated and phased SNVs, TRs and complex SVs directly from bulk tissue. Most single-cell genome studies have focused on SNVs, small indels and large copy-number changes, while long repeats and breakpoint-complex rearrangements remain difficult to reconstruct from short, amplified fragments. SniffCell retains the long-molecule information required to characterize these events and adds cellular context that is otherwise lost during bulk analysis. The PBMC results demonstrate that the framework is not restricted to the brain and can recover known lineage-specific rearrangements as well as localize less predictable repeat instability to a particular immune-cell compartment. We further generated a novel tissue-specific ctDMR reference atlas, which expands the previous atlas by 8- to 27-fold across eight tissues, including blood, brain, kidney, breast, liver, colon, pancreas and lung, providing a basis for extending cell-type-aware variant analysis across diverse human tissues. Cancer represents a particularly important future application. Long-read tumor sequencing already resolves complex rearrangements, viral integrations, extrachromosomal DNA and allele-specific methylation from the same genome, but these signals remain mixtures of malignant, immune and stromal populations^54^. Linking variant-supporting molecules to their cellular origin could help distinguish tumor-specific rearrangements from variants carried by the surrounding microenvironment and reveal how complex somatic mutations are distributed across tumor compartments.

The current resolution of SniffCell is determined by the cell populations represented in the methylation reference and by the number of informative ctDMRs carried on each read. In the brain, the presented reference atlas robustly distinguishes neurons from oligodendrocytes but does not yet resolve individual neuronal subtypes or less abundant glial populations. We deliberately excluded the matched FANS data from the reference atlas construction so that these samples could provide a fully independent benchmark, making the callable genomic space reported here a conservative lower bound. Incorporating these and future sorted-cell or single-cell methylation references will increase marker density, expand the callable genome and support finer cellular distinctions without requiring any change to the underlying sequencing experiment. More broadly, SniffCell transforms native long-read genomes into a resource for cell-type-specific mutation discovery without the additional experimental cost and complexity of single-cell sequencing. It therefore enables existing and future long-read cohorts to be revisited for somatic TRs, SVs and other complex mutations at population scale. This level of cellular resolution should facilitate the discovery of somatic variation that has remained hidden within bulk tissues and help resolve persistent questions across human development, aging, neurological disease and other phenotypes.

## Acknowledgements

We thank the Queen Square Brain Bank for providing brain tissue used in this study. We are grateful to Viorica Chelban and Alexandra Pancikova for sharing *FGF14* genotypes for the MSA and PD brain samples. We thank George Morrow and Yanping Guo at the UCL Flow Cytometry Translational Technology Platform for their support with nuclei sorting of the brain samples, as well as UCL Genomics and the Earlham Institute for sequencing of single-nuclei brain samples.

This study was conducted in part as part of the NIH Common Fund consortium initiative, Somatic Mosaicism across Human Tissues (SMaHT) through awards UG3NS132105 and 1DA058229. Some data used in this work were generated by the SMaHT Network and provided by the SMaHT Data Analysis Center (DAC; UM1DA058230) on behalf of the Network. This research was supported by the NIH Common Fund through the Office of Strategic Coordination/Office of the NIH Director under awards UG3NS132105 and UM1DA058229. We acknowledge the SMaHT tissue procurement, data production and data coordination teams that generated and distributed the samples and datasets used in this study. The SMaHT Network and data resource are described in the SMaHT marker publication^1^.

This research was funded in whole or in part by Aligning Science Across Parkinson’s (grant no.: 000430) through Michael J. Fox Foundation for Parkinson’s Research. For the purpose of open access, the author has applied a CC BY public copyright license to all Author Accepted Manuscripts arising from this submission.

This project was supported by the Cytometry and Cell Sorting Core at Baylor College of Medicine with funding from the CPRIT Core Facility Support Award RP240432 and the NIH (CA125123, OD036336, and OD038251), and the assistance of Joel M. Sederstrom.

## Data availability

SniffCell is available at https://github.com/Fu-Yilei/SniffCell

Analysis scripts used in this study are available at https://github.com/Fu-Yilei/SniffCell-analysis

mtrplotter is available at https://github.com/Fu-Yilei/mTRplotter

The data from the four QSBB brains (bulk and sorted long read WGS, short read scWGS) will be released officially for controlled access via ASAP CRN Cloud and described in Zenodo

All SMAHT data are available at phs004193 and at https://data.smaht.org/.

Large-scale brain CARD brain data is available on ANVIL https://anvilproject.org/news/2025/08/25/card-data-release-2 or dbGap phs000979

## Methods

### Consent agreements

Two PBMC donors provided written informed consent for genomic studies under Baylor College of Medicine Institutional Review Board approved protocol H-43884. All individual IDs throughout this work are anonymized.

### Biological methods

#### FANS validation cohort brain cell sorting and sequencing

Fresh-frozen post-mortem human brain tissue was obtained from Queen Square Brain Bank (QSBB) for Neurological Disorders, London, United Kingdom. All subjects had given consent for brain donation and use in research, and ethics approval was provided by the NHS Health Research Authority Ethics Committee London-Central (REC reference 23/LO/0044). The donors included two neurologically healthy controls (bcontrol1 and bcontrol2), one Parkinson’s disease (bparkinsons) case and one multiple system atrophy (bmsa) case. All were male, and the post-mortem interval was 38-44 hours. The ages at death were 71, 76, 74, and 56, respectively. The frontal cortex was used for long-read sequencing. Brains with known *FGF14* genotypes were chosen to ensure a range of values^55^. Single-cell whole-genome sequencing (scWGS) datasets were generated from the internal capsule of the MSA donor and the cingulate cortex of both controls and the PD donor.

Nuclei isolation was performed using one of two methods depending on the sequencing experiment and tissue cohort. For all bulk long-read sequencing experiments, nuclei were isolated from 100-500 mg of fresh-frozen frontal cortex using a previously described iodixanol density-gradient protocol^56–59^. For scWGS, approximately 10-50 mg of fresh-frozen tissue was used. Nuclei from the MSA internal capsule were isolated using the same manual iodixanol density-gradient protocol, whereas nuclei from cingulate cortex tissue from bcontrol1, bcontrol2 and the bparkinsons donor were isolated using the Minute™ Single Nucleus Isolation Kit for Neuronal Tissues/Cells (Invent Biotechnologies, BN-020).

Following isolation, immunolabelling and preparation of nuclei for FANS were performed^60^. with modifications according to the target nuclear population. Isolated nuclear pellets were gently resuspended in 500 μL of pre-chilled blocking buffer (PBS; Invitrogen™, AM9625) supplemented with 10% goat serum (Sigma-Aldrich, G6767) and 1× cOmplete™ EDTA-free Protease Inhibitor Cocktail (Roche, 04693132001), and incubated for 30 min at 4 °C with continuous rotation to block non-specific antibody binding. Depending on the experiment, nuclei were labelled using either directly conjugated primary antibodies or an unconjugated primary antibody followed by a fluorophore-conjugated secondary antibody. Primary antibodies were incubated at 4 °C with continuous rotation for 30 min - 2 h or overnight, as appropriate. Where secondary antibody labelling was required, nuclei were pelted by centrifugation (800 × g, 10 min, 4 °C), washed in pre-chilled blocking buffer, and re-pelleted under the same conditions. Secondary antibodies were then added in the blocking buffer and incubated for 1 h at 4 °C with continuous rotation. Unstained controls and, where secondary antibody labelling was required, secondary-only controls were prepared alongside the stained samples.

For bulk long-read sequencing experiments, neuronal nuclei were labelled with Alexa Fluor 488-conjugated anti-NeuN (Millipore, clone A60, MAB377X; 1:100), and oligodendrocyte nuclei were labelled with Alexa Fluor 647-conjugated anti-Olig2 (Abcam, clone EPR2673, ab225100; 1:500). For scWGS of Control 1, Control 2 and the PD donor, neuronal nuclei were labelled using the same NeuN antibody. For scWGS of the MSA donor, oligodendrocyte nuclei were labelled with anti-SOX10 (clone SP267, ab227680; 1:100) and anti-α-synuclein (Santa Cruz, clone 211, sc-12767; 1:100), followed by Alexa Fluor 488- and 647-conjugated secondary antibodies (Thermo Fisher Scientific; 1:1,000). Alpha-Synuclein staining was detected in 3.9% of SOX10+ nuclei63. Both α-synuclein-positive and -negative SOX10+ nuclei were included to increase sample size.

DAPI (Sigma-Aldrich, D9542) was added at a final concentration of 0.1 μg/mL, and nuclei were incubated for 30 min. Following staining, nuclei were pelted by centrifugation (800 × g, 10 min, 4°C), washed twice with PBS, and resuspended in a sorting buffer (PBS containing 5 mM EDTA; Sigma-Aldrich, 03690). Suspensions were passed through a 70 μm Flowmi™ cell strainer (Bel-Art, 15346248) to remove aggregates and debris prior to fluorescence-activated nuclei sorting (FANS) on a BD FACSAria™ Fusion cell sorter at the UCL Cancer Institute Flow Cytometry Translational Technology Platform.

During FANS, doublets and aggregates were sequentially excluded using forward- and side-scatter parameters. Forward-scatter singlets were identified using FSC-A versus FSC-H, followed by selection of side-scatter singlets using SSC-W versus SSC-A. DAPI-positive events were then gated to identify intact nuclei, after which antibody-positive populations were selected based on clear separation relative to the corresponding negative controls.

For bulk long-read sequencing experiments, approximately 400,000–500,000 nuclei were collected per tube into 100 µL PBS. Sorted nuclei were pelted by centrifugation (800 × g, 10 min, 4 °C), and the supernatant was removed before DNA extraction. For single-cell whole-genome sequencing experiments, individual nuclei were sorted directly into 96-well PCR plates (Starlabs, E1403-0100), with each well containing 5 µL TE buffer (Thermo Fisher Scientific, 12090-015).

#### DNA extraction for long-read sequencing of brain samples

For sorted bulk nuclei and brain tissue samples intended for Oxford Nanopore long-read sequencing, genomic DNA was extracted using a phenol–chloroform-based method. For tissue extractions, approximately 20 mg of fresh-frozen tissue was used. Briefly, 300 µL of lysis buffer (5 mM EDTA, 200 mM NaCl, 100 mM Tris pH 8.0, 0.2% SDS) was added to each sample and supplemented with 20 µL of Proteinase K (20 mg/mL; Thermo Fisher Scientific, EO0491). Samples were incubated for 5 min at room temperature with vigorous shaking, followed by 2 h at 56 °C. Subsequently, RNAse A Solution (1.5 µL for isolated nuclei, 20 µL for tissue, QIAGEN, 158153) was added to each sample and left for 30 minutes at 37 °C. Phenol:chloroform:isoamyl alcohol (25:24:1, v/v; Invitrogen, 15593031) was added at a 1:1 volume, and samples were vortexed for 30 seconds and centrifuged at maximum speed for 5 min at 4 °C. The aqueous phase was transferred to a fresh tube, mixed with 1 µL of glycogen (20 mg/mL; QIAGEN, 158930), and DNA was precipitated with 3 volumes of ice-cold 100% ethanol. Precipitation was carried out at -20 °C overnight. DNA was pelleted by centrifugation (30 min, maximum speed, 4°C), washed with 70% ethanol, and centrifuged again for 5 min. After removing residual ethanol, pellets were air-dried for 5–10 min and resuspended in 50–100 µL of TE buffer (pH 8.0) (Invitrogen, 12090015) or ultrapure nuclease-free water (Ambion, 10977015).

#### Single-cell whole-genome amplification and sequencing

scWGS was performed on FANS-isolated nuclei. DNA from individual SOX10-positive oligodendrocyte nuclei from the bmsa donor was amplified and libraries were prepared using the PicoPLEX Gold® Single Cell WGA Kit (Takara Bio, R300669) according to the manufacturer’s instructions. Libraries were sequenced using 150 bp paired-end reads on an Illumina NovaSeq platform at the Earlham Institute, Norwich UK. For the control and PD donors, DNA from individual NeuN-positive neuronal nuclei was amplified using the PicoPLEX® Single Cell WGA Kit (Takara Bio, R300672) according to the manufacturer’s instructions, followed by library preparation with a PCR-free kit (Illumina, 20041795) and sequenced using 150 bp paired-end reads on an Illumina NextSeq.

#### PBMC cell sorting and sequencing

Fresh whole blood (40 mL) was diluted 1:1 with PBS (Gibco 10010049), layered over Ficoll-Paque (Fisher Scientific 45001749), and centrifuged at 300 × g for 30 min at room temperature with slow acceleration and minimal brake. The peripheral blood mononuclear cell (PBMC) layer was collected from the plasma-Ficoll interface, washed in PBS, and residual red blood cells were lysed with RBC Lysis Buffer (BioLegend 420301) for 5 min on ice. Viable cells were counted with an automated counter (Invitrogen AMQAX2000). 5 × 10⁶ cells were reserved as the PBMC control and washed with PBS, pelleted, and snap-frozen with liquid nitrogen. 1 × 10⁶ cells were collected for unstained and single-stain controls. The remaining cells were used for sorting.

Single-stain controls and the sorting population were resuspended in PBS at 1 × 10⁷ cells/mL. Cells were stained with Zombie Violet™ (1 μL per million cells) (BioLegend 423113) for 15-30 min at 4 ℃ in the dark then washed. Fc receptors were blocked with Human TruStain FcX™ (BioLegend 422301) for 5-10 min at room temperature. Cells were then stained on ice for 15-20 min in the dark with antibodies against CD3 (FITC; clone SK7 or OKT3), CD19 (APC; clone SJ25C1 or HIB19), CD56 (PE; clone 5.1H11 or W22097A), CD14 (Brilliant Violet 605™; clone HCD14 or 63D3), and CD45 (APC/Cyanine 7; clone 2D1), supplied either as a pre-mixed TBNK cocktail (BioLegend 391501, 20 μL per million cells) with CD14 added separately (BioLegend 325639, 1:30), or as individually titrated antibodies (BioLegend 317306, 302212, 343754, 367126, and 368516; 5 μL per million cells each). Unless otherwise noted, blocking and staining were performed according to the BioLegend Cell Surface Flow Cytometry Protocol.

Cells were sorted on Sony MA900 (70 μm nozzle) into T cells (CD3⁺), B cells (CD19⁺), monocytes (CD14⁺), and NK cells (CD56⁺) (**Supplementary Fig. 26**), collected into 15-mL conical tubes with a maximum of 1 × 10⁷ cells per tube. Sorted cells were washed with PBS, pelleted, and snap-frozen with liquid nitrogen.

#### ONT library preparation and sequencing

Genomic DNA was quantified using the Qubit dsDNA Quantification Broad Range Assay (Thermo Fisher Scientific). DNA size was determined using the Agilent Femtopulse. For bulk samples, 6-8 µg of DNA was sheared using g-tubes (Covaris PN 520079) and size selected on the PippinHT instrument (Sage Science) using the 6-10 kb High-Pass definition to achieve an average size of 18-25 kb. The size-selected DNA was used as input for the ONT SQK-LSK114 library preparation kit. End repair/damage repair and adapter ligation were performed according to the manufacturer’s instructions. Final libraries were quantified using the Qubit dsDNA Quantification Broad Range Assay (Thermo Fisher Scientific). Each library had sufficient yield to load at least 2 ONT PromethION R10.4.1 flow cells with 15 fmoles of material. Sequencing was performed for 72 hours and if needed, flow cells were washed and re-loaded up to 2 times. For samples requiring more than 2 flow cells, additional libraries were prepared.

DNA extracted from FACS/FANS samples typically had a low yield (< 1 µg). A modified, low-input library preparation protocol was optimized for these samples. Starting with 500 ng or 1,000 ng of genomic DNA, samples were sheared at 1,100 rcf to an average size of 10-15 kb. Sheared samples were purified using 1X AMPure XP beads and used as input for the ONT SQK-LSK114 library preparation kit. End repair/damage repair and adapter ligation were performed according to the manufacturer’s instructions. Final libraries were quantified using the Qubit dsDNA Quantification Broad Range Assay (Thermo Fisher Scientific). Libraries were loaded on ONT PromethION R10.4.1 flow cells with 7-10 fmoles of material. Sequencing was performed for 72 hours and if needed, flow cells were washed and re-loaded up to 2 times. If sufficient library was available for loading a second flow cell, additional sequencing was performed.

#### Large-scale brain cohort (CARD/HBCC)

Long-read whole genome sequencing data for the large-scale brain cohort of 155 donors were generated previously as part of the CARD long-read initiative^45^. In brief, frontal cortex tissue from donors of African or African admixed ancestry was obtained from the Human Brain Collection Core (HBCC), NIMH Intramural Research Program, under protocols approved by the CNB IRB (NCT03092687). Mean age at death was 44.2 years (range 18.1 to85.2 years) and 36.1% were female. Samples were sequenced on the ONT PromethION platform (R10.4.1 flow cells, SQK-LSK114 kit) to an average depth of 37.42x. Full donor demographics, sequencing statistics and sample-processing protocols are described previously^61^. Sequencing data are available under dbGaP accession phs000979 and via AnVIL.

### Computational methods

#### SniffCell program

The SniffCell program consists of 4 major modules: **find, deconv, discover, and anno**. The SniffCell find module detects cell-type-specific differentially methylated regions (ctDMRs) from aggregated methylation profiles using a multi-class, one-versus-rest framework. Methylation input is provided as a matrix with genomic bins as rows and samples/cell-types as columns (aggregated raw methylation calls from GEO: GSE186458, see **supplementary**), where each value represents the mean methylation level of a bin in a given sample. A companion index table defines genomic coordinates and CpG span for each bin. Sample identities are read from metadata, and a user-defined JSON mapping assigns samples to major cell-type groups, allowing aggregation across biological replicates or related subtypes.

##### SniffCell find

We identified cell-type-differentially methylated regions (ctDMRs) from atlas methylation profiles using the SniffCell find module (v.0.9.4). The input comprised a genomic-bin-by-sample methylation matrix, corresponding sample identifiers and a genomic index specifying the chromosome, interval and CpG-index bounds of each bin. Samples were assigned to biological groups using a JSON mapping, with a user-specified top-level key defining the grouping scheme. For each genomic bin, reference methylation for each group was calculated as the mean across matched atlas samples.

SniffCell evaluated all non-redundant bipartitions of the specified groups. For each bin and bipartition, the absolute difference between the mean methylation of the two group sets was calculated, and the partition with the largest difference was selected. Both hypermethylated and hypomethylated contrasts were considered, and bins with an absolute methylation difference of at least 0.4 were retained. The smaller side of the selected partition was designated the target group set to favour more specific methylation signatures; when the two sides contained equal numbers of groups, the hypermethylated side was designated the target.

Qualifying bins were merged when they occurred on the same chromosome, shared the same selected bipartition, target group set and methylation direction, and were separated by no more than 2 kb. A non-qualifying bin interrupted merging. Candidate regions were required to comprise at least two index bins and three CpGs. For each ctDMR, SniffCell reported its genomic coordinates, target and comparator group sets, methylation direction, mean absolute methylation difference, mean methylation of the target and comparator sets, and group-specific regional methylation values, together with diagnostic measures of partition separation and comparator-group variability. The resulting annotation-ready TSV catalogue and IGV-compatible BED9 file were used in downstream SniffCell analyses to classify FANS-derived reads according to atlas reference methylation and to associate structural-variant-supporting reads with cell-type methylation signatures.

##### SniffCell deconv

We estimated cell-type composition from long-read methylation data using the SniffCell deconv module (v.0.9.4), with an aligned BAM file, the corresponding reference genome and an annotation-ready ctDMR catalogue generated by SniffCell find. For each ctDMR, CpG positions were identified from the reference sequence and methylation probabilities were extracted from the BAM base-modification tags to construct a read-by-CpG matrix. Probabilities for 5-methylcytosine and 5-hydroxymethylcytosine were combined as modified C. Unmapped, secondary and supplementary alignments were excluded. Reads without an informative CpG measurement were removed, and regions containing no reference CpGs or fewer than two informative reads were excluded. Missing measurements were imputed, where possible, using the mean value for that CpG across reads.

Reads were classified independently within each ctDMR as supporting either its target group set or the complementary group set. In the default closest_reference_mean mode, the mean methylation probability of each read across the ctDMR was compared with the atlas-derived target and comparator means (mean_best_value and mean_rest_value), and the read was assigned to the closer reference state. If these reference values were unavailable, SniffCell reverted to two-cluster k-means classification. The optional kmeans mode partitioned standardized read-level CpG profiles into two clusters (k = 2; random seed = 42) and oriented the clusters according to the expected hypermethylated or hypomethylated direction of the target group; atlas reference means were used when directionality was unavailable. Each local assignment was represented as a binary membership vector in the ordered cell-type schema of the ctDMR catalogue. Bits set to 1 denoted the leaf cell types compatible with the read’s assignment at that ctDMR, whereas bits set to 0 denoted the excluded cell types.

Evidence from multiple ctDMRs was subsequently integrated for each read. Within the shared cell-type schema, the binary membership vectors were combined by bitwise intersection, thereby retaining the cell types compatible with all local assignments and preserving hierarchical assignments when the evidence did not resolve a single leaf cell type. A non-empty intersection was accepted directly. When distinct ctDMR assignments produced an empty intersection, SniffCell used the most frequent complete assignment as a fallback and accepted it when its support was at least the specified agreement threshold (per_read_min_agreement; default, 0.66); otherwise, the read was designated as mixed. SniffCell reported individual ctDMR–read assignments, per-read consensus classifications and cell-type counts and fractions calculated either from all ctDMR–read observations or from one non-mixed consensus assignment per read. When requested, consensus-classified reads were further partitioned into user-defined cell-type groups and exported as indexed BAM files and accompanying read-level tables.

##### SniffCell discover

We performed cell-type-stratified variant discovery using the SniffCell discover workflow (v.0.9.4). The workflow compared two group-specific BAM files generated by SniffCell deconv; group identities and file paths were obtained from the accompanying split-group manifest. Structural variants, tandem repeats and CpG methylation were analysed in parallel, with optional small-variant calling.

Structural variants were initially called independently in each group using Sniffles in mosaic-aware mode, with germline inclusion and supporting-read reporting enabled. Calls were restricted to PASS variants satisfying the configurable mosaic expression (INFO/MOSAIC=1 by default). Sample genotypes were removed, the two site sets were concatenated, and redundant representations were collapsed using Truvari (maximum reference distance, 500 bp; minimum sequence and size similarity, 0.95). The resulting candidate sites were jointly genotyped against both group-specific BAMs using Kanpig. Variants required a minimum depth of five reads in both groups. Group-specific calls required at least two alternative-supporting reads in the designated group and no alternative-supporting reads in the comparator group, whereas variants supported by at least two alternative reads in both groups were classified as shared. Supporting-read names were retained for each group.

Tandem repeats were genotyped independently in each group using TRGT, which is the default repeat genotyper for both Oxford Nanopore and PacBio HiFi data in the current workflow. TRGT was run against a common repeat catalogue and reference genome, and its spanning-read BAM files were used for comparative post-processing. Repeat-length estimates were adjusted for the flanking sequence using the TRGT FL tag and pooled across haplotypes within each group. A locus was called as a cell-type or cell-type-group-associated expansion when both groups contained at least five spanning reads and the three longest reads from the expanded group each exceeded the longest comparator-group read by at least 50 bp. Homopolymer-associated calls were excluded by requiring a repeat-unit length of at least 2 bp. The expanded and baseline groups, supporting-read counts, effect size and supporting-read names were recorded for each locus.

Structural-variant, tandem-repeat and, when available, passing single-nucleotide-variant results were harmonized into a genomic-position-sorted TSV containing variant coordinates, class and subtype, group-specific category, change size, alternative-read counts and supporting-read names. All structural-variant categories, including shared calls, and all tandem-repeat candidates were retained in this table; downstream SniffCell anno excluded shared variants when evaluating cell-type-specific associations.

##### SniffCell anno

We assigned cell-type methylation evidence to candidate variants using SniffCell anno (v0.9.4). Candidate variants were supplied as either a structural-variant VCF or the harmonized variant table generated by SniffCell discover. For BAM-based annotation, the ctDMR reference was restricted to regions within a user-defined distance of each candidate variant (10 kb by default). ctDMRs overlapping the variant interval were excluded, and the exclusion interval could be extended on either side by a specified fraction of the absolute variant length to reduce the influence of methylation changes immediately adjacent to breakpoints. For harmonized variants with precomputed SniffCell deconv classifications, per-read methylation states could instead be imported directly; this route bypassed BAM rescanning and spatial ctDMR filtering and linked methylation evidence to variants through supporting-read identifiers. An important detail in SniffCell is of course also that we do not consider ctDMR that are within 50 bp of a variant of interest as this variant might impact the ctDMR directly.

Methylation was evaluated at each retained ctDMR using the read-classification procedure described above for deconvolution. Observations from multiple ctDMRs covered by the same read were consolidated before variant-level assignment. If the intersection yielded no compatible cell type, a plurality assignment was accepted only when its support exceeded the per-read agreement threshold (0.66 by default); otherwise, the read was classified as mixed and excluded from variant-level voting. Each remaining supporting read contributed a single vote. For every variant, SniffCell reported the total number of supporting reads, the number carrying informative methylation evidence, the number excluded as mixed, the majority binary code and the agreement fraction among retained votes. The binary assignments were decoded into human-readable cell-type labels using the hierarchy encoded in the ctDMR reference. An assigned code was reported only when the required overlap and agreement thresholds were met and the retained read-level assignments contained no hard conflict. Variants failing these criteria remained in the output as unassigned, preserving their underlying read and methylation evidence for subsequent review. Human-readable cell-type summaries could be calculated from either all ctDMR–read observations or one consolidated classification per read.

Individual loci can be examined using SniffCell viz. The visualization module recovered the annotation inputs and results from the run manifest and generated integrated locus plots showing the candidate variant, local read alignments, supporting and non-supporting reads, large insertion and deletion events inferred from read alignments, per-read ctDMR methylation measurements, atlas reference methylation states and the resulting cell-type assignments. Supporting reads with and without resolved cell-type assignments were displayed separately. Linked ctDMRs outside the plotted interval could be represented as distal callouts, allowing methylation evidence elsewhere on a long read to remain visible without expanding the primary locus view.

Annotation results were summarized using SniffCell report, which generated an interactive HTML report and corresponding tabular outputs for variant review. Report-level filtering was performed independently of the annotation thresholds. By default, high-confidence variants were required to have a linked cell-type assignment, no hard assignment conflict, a majority agreement of at least 0.8 and sufficient informative-read support. In the default gradient mode, the required number of informative reads was calculated as ceiling[0.8 × √(n supporting reads)], reducing the dependence of selection on variant read depth; a fixed overlap-fraction filter was also available. Tandem-repeat records were retained more permissively when linked cell-type evidence was present to facilitate manual review. Reports could optionally include the locus panels generated by SniffCell viz, IGV snapshots or interactive IGV views, together with review fields for recording variant-level assessment.

#### FACS/FANS validation cohort analysis

##### Processing of long-read sequenced samples

Oxford Nanopore Technologies (ONT) sequencing data were basecalled using the R10.4.1/E8.2 400-bp super-accuracy model (dna_r10.4.1_e8.2_400bps_sup@v4.3.0). Modified-base calling was enabled during basecalling. Reads passing the upstream quality-control and duplicate-filtering procedures were retained for downstream analysis.

Basecalled reads were aligned to the GRCh38 human reference genome using minimap2 v2.24 with the ONT long-read alignment preset (‘-ax map-ont’). The ‘-Y’, ‘--MD’, and ‘-y’ options were used to preserve supplementary alignment information, generate MD tags, and retain ONT-associated alignment information, respectively. Read-group information was assigned during alignment. The resulting alignments were coordinate-sorted using SAMtools, and BAM files generated from independent sequencing runs or flow-cell components corresponding to the same biological sample were subsequently merged to produce a single sample-level alignment file.

For haplotype-resolved analysis, phased germline variants were generated using Clair3 and used to assign individual ONT reads to parental haplotypes. Read-level haplotype tagging was performed with WhatsHap v2.3 using the GRCh38 reference genome. Read-group information was ignored during haplotagging to enable haplotype assignment across reads originating from multiple sequencing runs. The resulting haplotagged BAM files were used for downstream haplotype-specific genomic and epigenomic analyses.

##### Benchmarking of ctDMR from SniffCell find

To benchmark the SniffCell find module, we performed a parameter sweep over the minimum number of CpGs, the methylation difference between categories and the maximum gap between CpGs within a ctDMR. We selected the least stringent parameter combination that achieved >0.9 methylation similarity with the sorted reference data, resulting in default thresholds of three CpGs, a methylation difference of 0.4 and a maximum gap of 2,000 bp (**Supplementary Fig. 1**). We then characterized the number, size and genomic span of the resulting ctDMRs (**Supplementary Fig. 2a,b**). The cCRE v4 BED file was obtained from Moore et al^62^ and overlap with ctDMRs was quantified using BEDTools intersect (Supplementary Fig. 2c). Finally, SniffCell ctDMRs were compared with the Loyfer et al. atlas using BEDTools intersect (**Supplementary Fig. 2d**).

We next used the sorted samples to evaluate concordance between SniffCell ctDMRs and independently measured FACS/FANS methylation profiles. CpG methylation in the sorted samples was quantified using modkit v0.6.1. For each ctDMR, atlas-derived methylation values were compared with modkit pileup values from the corresponding sorted populations. We compared neurons with oligodendrocytes across four brain samples and monocytes with lymphocytes across the two PBMC samples. For the PBMC benchmark, T cells, B cells and NK cells were grouped as lymphocytes because SniffCell uses binary classification at each ctDMR and these populations show similar methylation profiles relative to monocytes. Concordance was quantified using Pearson’s correlation coefficient and directional concordance (**Supplementary Fig. 3**).

##### Comparison of SniffCell deconvolution with cell-sorted methylation data

We then compared the SniffCell deconvoluted reads’ DNA methylation with the FACS/FANS sorted data. We ran modkit on the SniffCell deconvoluted reads on 6 benchmarking samples (4 brains and 2 PBMCs) and then selected CpGs with >5x coverage on the genome and cross compared with sorted samples with the same locations. The correlation was evaluated with Lin’s Concordance Correlation Coefficient (CCC) and Pearson’s r and Mean Absolute Error (MAE) (**Supplementary Fig. 4**). We further investigated the concordance on the read level. This way we used a more stringent threshold where we investigated a random 1,000 ctDMRs and checked assigned reads on those regions having a single-read-level ctDMR similarity compared to the FACS/FANS sample. For example if a read in the PBMC sample is assigned to B cell we then compare it to B cell’s DNA methylation value on the read’s carried ctDMR region based on the read alignment (**Supplementary Fig. 5**). We assessed GENCODE v49 gene bodies and compared this to the rest of the genomic regions. A gene was considered accessible if at least 80% of its bases were covered by SniffCell-deconvoluted reads at ≥5× depth.

We further quantified the proportion of reads carrying conflicting cell-type information. A read was considered conflicting when multiple ctDMRs supported incompatible cell-type assignments; such reads were classified as unassigned by SniffCell. To test whether these conflicting brain reads carried blood monocyte-like methylation information, we built a three-way ctDMR reference using neuron, oligodendrocyte, and raw blood monocyte methylation profiles. For each exact ctDMR interval present in the original brain deconvolution output, we compared the monocyte methylation beta value to the neuron and oligodendrocyte beta values from the three-way reference atlas. The ctDMR was labeled monocyte-like for the brain state with the smaller absolute beta distance to the monocyte.

We then scanned the original per-read ctDMR assignment rows and restricted to the previously identified conflicting reads. For each read-ctDMR observation, the original brain assignment token was compared with the monocyte-like token for the same exact ctDMR interval. A conflicting read was counted as carrying monocyte-like information if at least one of its assessed ctDMR observations matched the monocyte-like state. We also computed stricter summaries requiring at least half, or all, of the assessed ctDMR observations on a read to be monocyte-like.

##### Reference genome coverage of deconvoluted reads

To evaluate sequencing coverage across genomic annotation classes, we used cell-type-partitioned BAM files generated by SniffCell from the new-catalog deconvolution analysis. The brain analysis included four bulk samples (bcontrol1–bparkinsons), each partitioned into Neuron and Oligodendrocyte reads, producing eight sample-by-cell-type BAM files. The PBMC analysis included two bulk samples (pbmc1 and pbmc2), each partitioned into Monocyte and combined T-cell/NK-cell/B-cell reads, producing four sample-by-cell-type BAM files.

Coverage was evaluated for five genomic region classes: GENCODE v49 basic gene bodies, GTEx tissue-specific gene bodies, medically relevant genes (MRGs), ENCODE candidate cis-regulatory elements (cCREs), and the remaining genome. The “all other genome” category was defined as the complement of the union of GENCODE genes, MRGs, and cCREs. The annotation classes were analyzed independently and were not otherwise required to be mutually exclusive.

Coverage was calculated using mosdepth with the options “-n -x --by regions.bed --thresholds 1,5,10.” For each annotation interval, mosdepth reported the number of bases covered by at least five reads. For each sample-by-cell-type BAM and annotation class, the percentage of bases covered at ≥5× was calculated as 100 multiplied by the total number of interval bases covered at ≥5×, divided by the total number of annotated bases in that class. This is therefore a base-weighted coverage measurement rather than the percentage of genes or intervals passing a coverage threshold. Annotated interval lengths were summed directly, so a genomic base contained in overlapping gene annotations could contribute once for each annotated gene interval.

GTEx tissue-specific genes were defined using the GTEx v8 median TPM expression matrix and GENCODE v49 gene identifiers after removal of gene-version suffixes. Expression values were transformed as log2(TPM + 1). For the brain, the matched expression group consisted of “Brain – Cerebellar Hemisphere” and “Brain – Cerebellum,” with expression averaged across these two tissues. For PBMC, “Whole Blood” was used as the matched GTEx tissue.

A gene was classified as tissue-specific when three criteria were met: its maximum GTEx median expression occurred in the matched tissue group; its mean matched-tissue expression was at least 1 on the log2(TPM + 1) scale; and its matched-tissue expression exceeded the median expression across all other GTEx tissues by at least 1 on the same scale. The analysis was restricted to 54,626 GENCODE genes represented in the GTEx matrix. These criteria identified 4,727 brain-specific genes and 720 whole-blood-specific genes. Coverage of these genes was obtained by subsetting the existing per-gene mosdepth results and applying the same base-level ≥5× calculation.

For each annotation class, the final brain value was calculated as the arithmetic mean of the eight brain sample-by-cell-type percentages. The final PBMC value was the arithmetic mean of the four PBMC sample-by-cell-type percentages. The values displayed above the bars were rounded to the nearest whole percentage. No statistical comparisons or error bars are shown in this figure.

The unrounded brain coverage values were 71.19% for GENCODE genes, 76.76% for GTEx tissue-specific genes, 73.55% for MRGs, 78.35% for cCREs, and 48.02% for the remaining genome. The corresponding PBMC values were 29.47%, 69.26%, 31.56%, 37.59%, and 11.81%, respectively.

##### FACS/FANS validation cohort read-assignment analysis

Read-assignment accuracy was evaluated using 16 experimentally sorted samples, comprising neuron and oligodendrocyte brain fractions from four donors and T cells, B cells, NK cells, and monocyte PBMC fractions from two donors. SniffCell deconvolution was performed independently for each sample using the tissue-matched ctDMR reference atlas. A ctDMR-covered read was classified as an exact assignment when its non-mixed result contained only the expected sorted cell-type. A read was classified as compatible when the expected cell-type was included in a non-mixed multi-label assignment, such as T cells|B cells or T cells|NK cells for a T cells FACS sample. Mixed assignments and reads without a resolved leaf-cell-type assignment were classified as ambiguous/not assigned. Resolved assignments that did not contain the expected FACS/FANS cell-type were classified as flipped/wrong. Sample-level compatible accuracy was calculated as the combined number of exact and compatible reads divided by all ctDMR-covered reads, and tissue-level accuracy was calculated as the macro-average across samples.

##### Single-cell DNA sequencing analysis

Sequencing reads from the bmsa donor were trimmed using Trimmomatic v0.39^63^ to remove the 14-bp PicoPLEX amplification adapter and aligned to the GRCh38 reference genome (no-alt) using Bowtie2 v2.5^64^. Resulting BAM files were sorted and indexed using SAMtools v1.22^65^. Read groups were added and duplicate reads were marked using Picard v3.3.0^66^.

For bcontrol1, bcontrol2 and the bparkinsons donor, reads were trimmed using Trim Galore v0.6.10 with Cutadapt v5.2 to remove the 14-bp PicoPLEX amplification adapter and Illumina adapters (stringency 8; no mismatches allowed), together with poly-G trimming (minimum length 10 bp; no mismatches allowed). Trimmed reads were aligned to the GRCh38 reference genome (no-alt) using BWA-MEM v0.7.17-r1188, and resulting BAM files were sorted using SAMtools v1.21. Duplicate reads were marked using Picard v3.3.0.

Single-cell whole-genome sequencing (scWGS) coverage was evaluated across four brain samples: two controls (bcontrol1 and bcontrol2), one Parkinson’s disease sample (bparkinsons), and one multiple system atrophy sample (bmsa). Each sequencing run corresponded to an individually processed cell. Runs were grouped according to their curated neuronal or oligodendrocyte/glial assignment. The oligodendrocyte category included NeuN-negative glial cells when more specific glial subtype information was unavailable. The final dataset comprised 528 scDNA runs: bcontrol1, 104 runs (90 neuronal and 14 oligodendrocyte/glial); bcontrol2, 157 runs (93 neuronal and 64 oligodendrocyte/glial); bparkinsons, 221 runs (93 neuronal and 128 oligodendrocyte/glial); and bmsa, 46 oligodendrocyte runs. No neuronal runs were available for bmsa. BAM integrity was assessed before analysis, and one neuronal bcontrol1 BAM that failed validation was excluded.

##### Plotting of somatic tandem repeats

Somatic tandem-repeat candidates were visualized using mTRplotter (v0.1.1, https://github.com/Fu-Yilei/mTRplotter). The tool accepts either a single genomic interval or a BED file of candidate loci, together with a sample manifest specifying the sample identifier, tandem-repeat analysis directory, caller (Medaka tandem or TRGT), flanking-sequence length and plotting metadata. TRGT loci were validated against the corresponding VCF and reads were retrieved from the spanning-read BAM. The length of each repeat-containing molecule was calculated after removing the left and right flanking sequences. Flank lengths were obtained from per-read padding coordinates for Medaka FASTA records, from the FL tag for TRGT reads or, when these were unavailable, from the sample manifest. Haplotype labels were recovered from the Medaka read identifiers or the TRGT HP tag.

For each locus, mTRplotter displayed one point per spanning read, with samples or cell-type groups arranged along the horizontal axis and flank-corrected repeat length on the vertical axis. The same framework was used for the FACS/FANS validation analyses, the SMaHT brain cohort and the large-scale brain cohort analyses. In comparisons of SniffCell-derived and experimentally purified populations, reads were coloured by cell type and distinguished by source, with SniffCell-classified reads shown as circles and FACS/FANS reads as triangles. In the multi-region SMaHT analyses, colours denoted brain region or blood and marker shapes distinguished sequencing platforms. Candidate loci were supplied as cohort-specific BED files, and loci not assayed in every sample were permitted. In addition to the locus-level figures, mTRplotter generated per-read tract-length tables and sample- and haplotype-level summaries used for downstream quantitative analyses.

##### FACS validation cohort PBMC sample recombination analysis

Deletion-like V(D)J recombination events were identified directly from long-read alignments at known T cells receptor and immunoglobulin recombination loci. We used two complementary sources of evidence: large reference deletions encoded in the CIGAR string and deletion-like split-read alignments inferred from supplementary alignment tags. For each BAM file, reads overlapping the predefined recombination intervals were scanned, and candidate events were retained if the estimated deletion span was at least 1,000 bp. For CIGAR-supported events, the deletion size was taken directly from reference-consuming D operations, which represent reference bases skipped by the read. For split-read-supported events, the primary and supplementary alignments were parsed and converted into both reference-space and read-space coordinates using their CIGAR strings. A split-read event was considered deletion-like only if the two aligned segments mapped to the same chromosome, had the same strand orientation, followed the expected read-to-reference order, and produced a positive genomic gap within the recombination interval. The deletion size was then calculated as the distance between the end of the upstream alignment block and the start of the downstream alignment block, corresponding to the reference sequence absent from the read but spanned by the read across the recombination junction. We further applied k-means clustering to deletion sizes in the SniffCell output, yielding 13 T-cell and 3 B-cell events.

To reduce false-positive events from ambiguous supplementary alignments in repetitive immune loci, especially IGH, we applied strict alignment filters. Both the primary and supplementary alignments were required to have mapping quality ≥20, the inferred breakpoints had to fall within the designated recombination locus, and the two aligned read segments had to be nearly adjacent on the read, allowing no more than 100 bp of query gap or query overlap. This query-adjacency filter is biologically motivated: true V(D)J recombination can delete tens to hundreds of kilobases, or more, from the reference genome, but the recombined molecule contains a clean junction between two distant genomic segments, with only a short junctional sequence on the read. Thus, a large genomic gap together with a small read-level gap provides a structurally coherent signature of recombination rather than a chimeric or repeat-driven alignment artifact. CIGAR-based deletions and split-read-based deletion estimates were combined for downstream visualization because both represent deletion-like loss of reference sequence at the recombination site.

The expected size range of these events was determined from the hg38 reference organization of antigen-receptor gene segments. Because V(D)J recombination joins selected V, D, and J segments and removes the intervening genomic sequence, the expected deletion span is defined by the physical distance between the recombined segments on the reference genome, rather than by the length of the V, D, or J genes themselves. For two non-overlapping segments, this distance was calculated from the hg38 segment boundaries using a consistent coordinate convention. This reference-based calculation predicts locus-specific deletion ranges, including approximately 46–498 kb for TRB, 0.36–0.85 Mb for TRA/TRD examples, ∼0.11 Mb for TRG, 0.08–1.01 Mb for IGH, 0.09–0.24 Mb for common deletion-like IGK V–J examples, and up to ∼0.89 Mb for distal IGL V–J examples. Observed long-read deletion estimates were therefore interpreted relative to these locus-specific reference-derived ranges. Events were colored by whether they occurred in the expected cell-type group for each locus, with TCR loci expected in T cells or lymphocyte-enriched samples and immunoglobulin loci expected in B cells or lymphocyte-enriched samples; bulk PBMC samples were plotted separately because they contain a mixture of lymphocyte populations. This approach provides both a direct read-level estimate of deletion size and a biological consistency check based on the expected same-chromosome, same-strand, sequential recombination geometry of V(D)J rearrangement.

##### FANS validation cohort brain *FGF14* TR expansion flanking sequence analysis

Cell-type-specific repeat-length distributions at the *FGF14* GAA repeat locus (GRCh38, chr13:102,161,539–102,161,869) were evaluated using TRGT output generated by SniffCell v0.9.4. Analyses were restricted to neuron- and oligodendrocyte-assigned reads from four full-depth brain samples. For each spanning read, the repeat-containing sequence length was calculated after removing the flanking sequence specified by the TRGT FL tag. Per-read repeat length was then expressed in GAA-repeat units by adjusting the motif count of the assigned consensus allele according to the difference between the read and consensus allele lengths. The 5′ sequence context was examined from the conserved upstream sequence through the beginning of the repeat tract. An allele was classified as carrying the common 5′-flanking variant (5′-CFV) when the sequence CAACCAACTTTCTGTTAGTCATAGTACCCCAA was present immediately upstream of the repeat. Alleles lacking the TTAGTCATAGTACCCCA segment and containing the reference flanking sequence CAACCAACTTTCTGT were classified as non-CFV. Classification was performed by exact sequence matching in the TRGT consensus alleles and checked in the corresponding Medaka consensus VCFs and allele-assigned spanning reads. The 5′-CFV was considered a flanking polymorphism rather than an interruption of the GAA tract. In each of the four donors included in the figure, the shorter allele carried the 5′-CFV, whereas the longer allele carried the reference 5′ sequence.

For sequence-interruption analysis, the principal repeat tract was identified from the TRGT MS annotation, and cyclic rotations of the canonical motif (GAA, AAG, and AGA) were treated as equivalent. Consensus sequences from both TRGT and Medaka were screened for the reported AGG and A-rich AAAGAAGAAG interruption patterns. Read-level screening was restricted to the internal repeat-containing sequence to avoid matching the external flanks. The analysis was independently repeated in the matched FANS data using the Medaka consensus sequences and reads aligned to the longer, expanded haplotype-specific consensus. A putative somatic interruption was considered supported only when a noncanonical motif absent from the consensus allele recurred in multiple expanded neuron reads. Isolated sequence differences in individual ONT reads were not interpreted as somatic interruptions because long, low-complexity repeats are susceptible to read-level insertion/deletion and alignment errors.

##### FACS/FANS validation cohort TR and SV calling

SniffCell tandem-repeat (TR) and structural-variant (SV) candidates were summarized for two peripheral blood mononuclear cell (PBMC) samples and four brain samples using the full-depth data and independently processed 60x and 30x downsampled datasets. PBMC TR candidates were required to have at least five reads supporting the change (n_change_support_reads >= 5), at least five total change-group reads (n_change_reads >= 5), and at least ten baseline-group reads (n_baseline_reads >= 10).

The starting full-depth brain callset required n_change_support_reads >= 2, n_change_reads >= 5, n_baseline_reads >= 5, and motif_size >= 3. Each candidate was annotated for change length, recovery in the 60x and 30x datasets, recurrence in other donors, and supporting-read sequence complexity. The sequence-complexity score counted four artifact-associated properties: median GC content <0.25, median sequence entropy <0.86, top 4-mer fraction >0.12, and dinucleotide purity >0.28. SV candidates were required to have site coverage n_overlapped >= 15. Events with hard cell-type assignment conflicts or links spanning all cell types represented in the corresponding tissue were excluded. To remove germline or shared candidates, remaining SVs were compared with the run-level shared-both-groups AD-logic VCF, and overlapping events were removed. This SV filtering procedure was applied independently and consistently to the full-depth, 60x, and 30x datasets. For each sample and depth, retained TR and SV candidates were counted and displayed as stacked bars. The plotted full-depth callset contains 122 candidates: 115 TRs and 7 SVs. Across all displayed depth/sample combinations, the figure contains 187 detections.

#### SMaHT brain cohort analysis

The samples were obtained from the SMaHT consortium. Regional distributions were evaluated for 25 neuron-specific and 43 oligodendrocyte-specific TR loci. A locus was considered observed in a brain region when at least one expansion call for that locus and cell-type was detected in a donor sampled from that region. Regional locus counts represent unique loci observed in each region, irrespective of the number of supporting donors.

To account for unequal regional sampling, prevalence was calculated using donor–locus opportunities. For each cell-type and region, the number of unique donor–locus expansion calls was divided by the product of the number of cell-type-specific loci and the number of donors sampled in that region. The analysis included 12 donors with cerebellum samples, 9 with frontal-lobe samples, 8 with temporal-lobe samples, and 8 with hippocampal samples.

Regional enrichment was assessed using donor-stratified permutation tests. For each donor–locus pair, we preserved the brain regions sampled for that donor, the number of sampled regions containing the expansion call, and the cell-type classification of the locus. The observed regional labels were then randomly reassigned without replacement among the regions available for that donor. This procedure preserves donor-specific sampling, locus recurrence, and the regional breadth of each expansion while testing the null hypothesis that calls have no preferential regional distribution.

Cell-type-associated tandem repeat (TR) expansions were evaluated against matched blood TR genotypes to distinguish brain-enriched alleles from alleles also detectable in blood. We restricted the analysis to Neuron and Oligodendrocyte expansion calls at globally cell-type-exclusive loci. A locus was considered cell-type-exclusive when expansion calls occurred in only one of the two cell types across the individuals. Recurrent events were defined as exclusive events observed in at least two donors. Matched blood genotypes were obtained from TRGT VCFs. Same-donor ONT blood TRGT was preferred; PacBio blood TRGT was used when ONT blood was unavailable or under-covered. For each SniffCell sample-level call, the matched blood record was queried at the same TR coordinates and allele lengths and allele-support read counts were extracted from the TRGT genotype fields. A blood genotype was considered to support a SniffCell expansion if it had adequate coverage and at least one blood allele longer than the SniffCell boundary, with allele support greater than or equal to the call-specific supporting-read threshold. For each plotted event-donor row, Neuron and Oligodendrocyte values were normalized to matched blood as: cell-type gain = max(cell-type median allele length − matched blood maximum allele length, 0). Thus, blood is fixed at zero and zero-valued cell-type points indicate no observed gain above the longest matched-blood allele.

For each event, the repeat motif was reduced to its primitive repeat unit and canonicalized across cyclic rotations and reverse complements. AAG-like repeats were defined as canonical motifs containing only A/G bases and a cyclic AAG, AGA, or GAA trinucleotide.

#### CARD brain cohort analysis

The samples were obtained from the CARD consortium, which includes 155 frontal-cortex donors. GAA-associated repeat loci were defined using the Adotto v2 lite tandem-repeat catalog. Motifs were restricted to ≤10 bp and were classified as GAA-element motifs when they contained at least one cyclic GAA element in either the reported orientation or its reverse complement. This definition therefore included rotationally equivalent and reverse-complement motifs, while excluding motifs such as GAGA that do not contain a cyclic GAA element. GAA-element loci were further divided into pure GAA, pure GAAA, pure GGAA, other 4-bp GAA-rich motifs, and other 5–10-bp GAA-rich motifs. Within CARD, the number of unique loci expanded in Neurons or Oligodendrocytes was calculated at recurrence thresholds of ≥1, ≥2, ≥3, and ≥5 donors. A locus could contribute to both cell types if qualifying expansions were detected in both Neuron and Oligodendrocyte samples.

For cross-cohort replication, calls were restricted to expansions of at least 100 bp. Loci were harmonized by exact GRCh38 chromosome, start, and end coordinates. A locus was considered Neuron-directed when at least one donor in that cohort had a Neuron expansion; the presence of an Oligodendrocyte expansion at the same locus did not exclude it from the primary analysis. A stricter Neuron-only analysis, requiring no Oligodendrocyte expansion in the same cohort, was performed as a sensitivity analysis. Pairwise replication was directional and was calculated as the number of source-cohort Neuron loci detected at the same coordinates in the target cohort divided by the total number of qualifying loci in the source cohort. Three-way replicated loci were required to have a qualifying Neuron expansion at identical coordinates in CARD, SMaHT, and the four-brain benchmark.

Enrichment of GAA-element loci among cross-cohort replicated loci was assessed using two-sided Fisher’s exact tests, comparing the replication frequencies of GAA-element and non-GAA loci within each source cohort. Replication percentages were source-normalized because cohort sizes and the total numbers of detected loci differed substantially. No-call loci were not interpreted as confirmed negatives because locus callability, sequencing inputs, and consensus construction differed among cohorts. All aggregation, statistical testing, and visualization were performed using Python with pandas, SciPy, NumPy, and Matplotlib.

