## Supplementary Information for "Cell-type-resolved somatic variant discovery from bulk long-read sequencing"

### 1. Re-analysis of normal human cell-type DNA methylation atlas and benchmarking with sorting samples

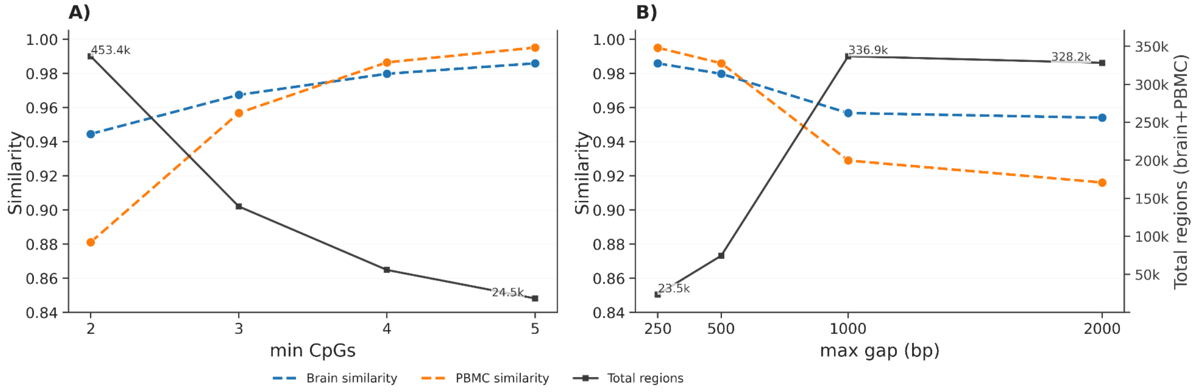


***Supplementary Fig. 1 | Genome-wide evaluation of SniffCell find parameterization for ctDMR detection. a, Influence of the minimum CpG requirement per region on ctDMR similarity and yield.*** *Increasing the minimum CpG threshold progressively improves concordance with reference FACS/FANS-derived profiles (brain and PBMC) but reduces the total number of detected regions. A threshold of ≥3 CpGs achieves high similarity while preserving substantial genome-wide coverage.* ***b, Influence of the maximum inter-CpG distance on ctDMR similarity and yield.*** *Restricting CpGs to ≤2,000 bp maintains high concordance (brain ~0.96; PBMC ~0.92) while retaining >100,000 ctDMRs genome-wide, representing a balanced trade-off between accuracy and region recovery.*

We obtained BEDgraph files from the normal human cell-type DNA methylation atlas (GEO accession: GSE186458), generated by whole-genome bisulfite sequencing (WGBS). These files provide genome-wide CpG methylation levels for each fluorescence-activated nuclei sorting (FACS/FANS)-purified cell-type profiled by Lofyer et al. [14] Given the high intra–cell-type homogeneity of DNA methylation patterns, we used this atlas as a reference framework to define cell-type-specific differentially methylated regions (ctDMRs) (See **methods**). These reference ctDMRs were then compared with ctDMRs derived from our independently generated brain, and blood and brain FACS/FANS datasets. We first performed a parameter sweep on the ctDMR discovery threshold where we tried to keep the mean DNA methylation similarity (1 - DNA methylation delta) between the ctDMR regions from the re-processed Lofyer et al.'s atlas and the FACS/FANS sorted samples greater than 0.9. Across the tested parameterizations, SniffCell exhibited a pronounced yield-concordance trade-off (**Supplementary Fig. 1**). Increasing minimum required CpGs improved agreement with orthogonal reference data, with brain similarity increasing from 0.944 to 0.986 and PBMC similarity from 0.881 to 0.995 as minimum required CpGs was raised from 2 to 5, but at the cost of a marked reduction in callable ctDMRs, from a mean of 453.4k to 24.5k regions (**Supplementary Fig. 1a**). By contrast, increasing maximum allowed gaps among CpGs (max_gap_bp) generally increased yield, expanding the mean number of callable ctDMRs from 23.5k at 250 bp to 336.9k at 1000 bp, with similarly elevated yield at 2000 bp (328.2k), while reducing concordance (brain, 0.986 to 0.954;

PBMC, 0.995 to 0.916; **Supplementary Fig. 1b**). During the parameter sweep we observed that selecting at least 3 CpGs (**Supplementary Fig. 1a**) among 2000 regions (**Supplementary Fig. 1b**) are sufficient enough to maintain a DNA methylation similarity (> 0.9) between the altas and the FACS/FANS sorted samples.

#### 1.1. Benchmarking sniffcell find module with FACS/FANS samples

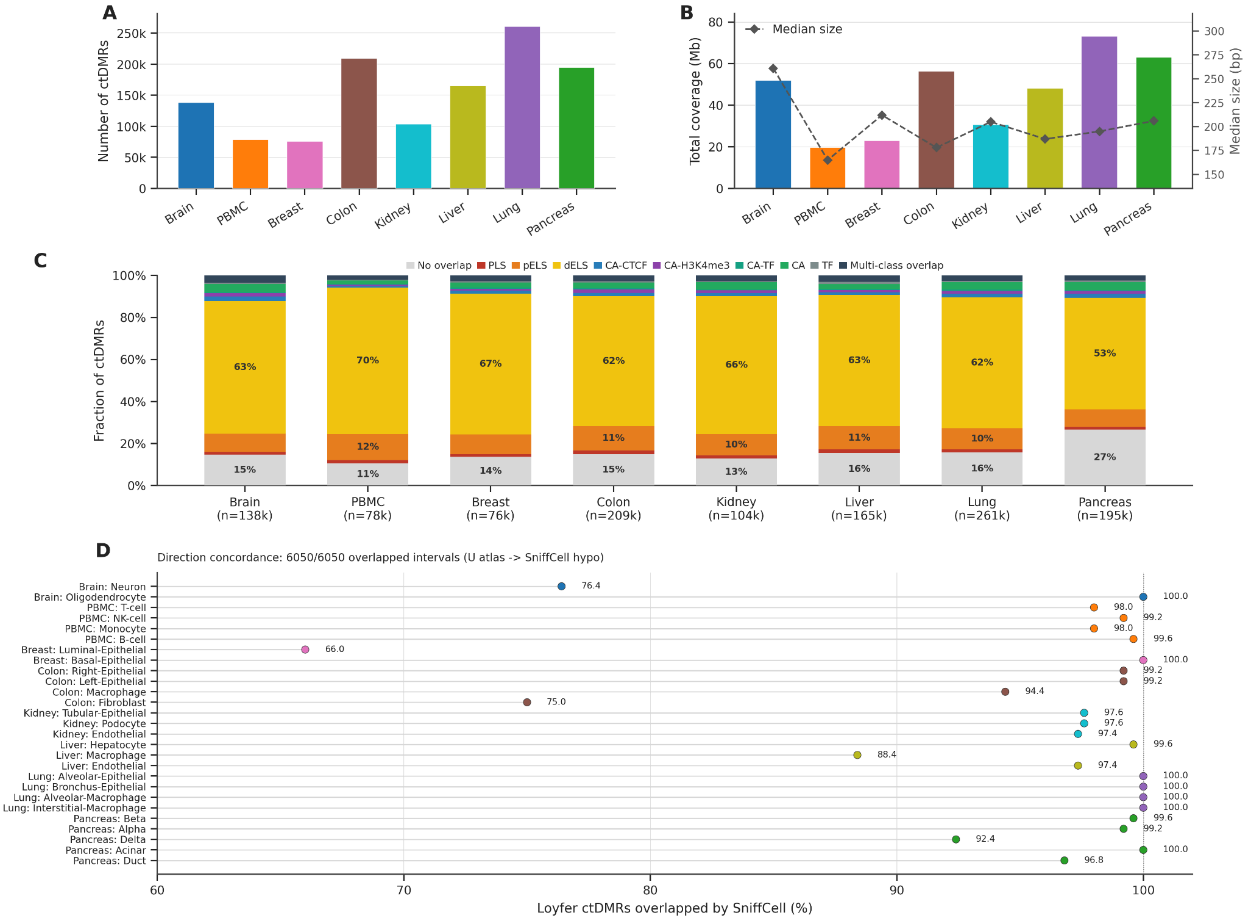


***Supplementary Fig. 2 | Genome-wide distributions of ctDMR in 8 major tissues. a. The number of ctDMRs of 8 major tissues. b. The average genome coverage and size of the ctDMRs of 8 major tissues. c. The cCRE overlap of the ctDMRs of 8 major tissues. d. the overlap with the corresponding cell-types compared to Lofyer et al.,'s cell-type selections.***

To further characterize cell-type-resolvable tissues, we applied SniffCell to eight common human tissues using the same approach. Lung exhibited the largest number of ctDMRs (261k), whereas Breast contained the fewest (76k; **Supplementary Fig. 2A**). We next examined the genomic coverage and median size of ctDMRs across tissues; total ctDMR coverage ranged from 19.6 Mb in PBMC to 73.2 Mb in Lung, with median region sizes ranging from 165 bp to 261 bp (**Supplementary Fig. 2B**). Consistent with preferential localization to regulatory sequence, 85.2% of brain ctDMRs and 89.4% of PBMC ctDMRs overlapped cCRE4 annotations, with the region-based breakdown dominated by enhancer-like classes, particularly distal enhancer-like signatures (dELS; 63.2% and 69.6%, respectively) and proximal enhancer-like signatures (pELS; 8.7% and 12.4%; **Supplementary Fig. 2C**). Across tissues, ctDMRs showed similar enrichment in enhancer-associated elements, with dELS comprising 53.1%-69.6% of regions and pELS comprising 8.4%-12.4% (**Supplementary Fig. 2C)**. These results suggest that tissue-specific methylation differences are largely associated with enhancer regulatory elements. From the DNA methylation atlas of normal human cell types, we directly obtained their 9,520 ctDMR regions stratified to specific cell-types. We further assessed how well the SniffCell reference atlas covered the corresponding the previous cell-type ctDMRs and observed coverage from 66.0% to 100.0% across matched cell-types, with all overlapped intervals showing 100% directional concordance (**6,050/6,050; Supplementary Fig. 2D**).

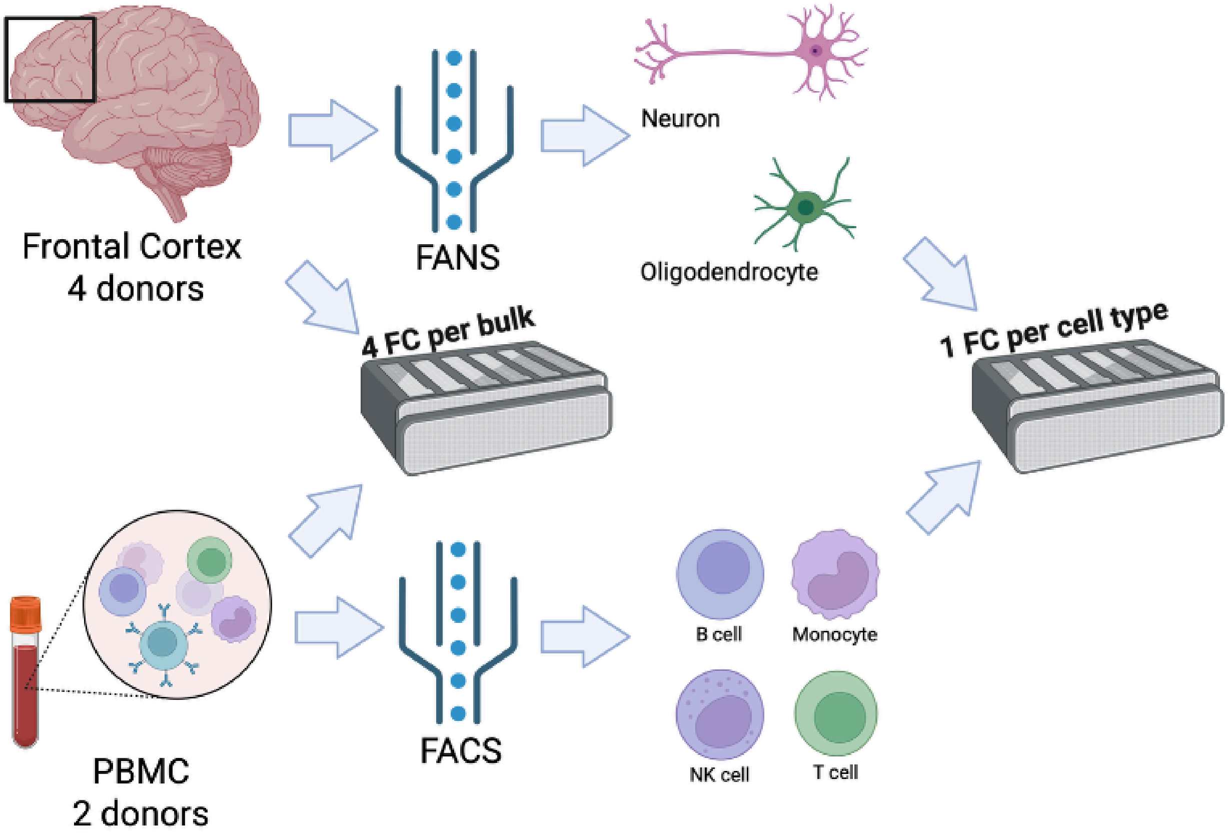


***Supplementary Fig. 3 | Schematic illustration of FACS/FANS experiments.*** *The sequencing of the bulk tissue is four flowcell ONT, while the sequencing for each FACS/FANS cell type is sequenced by one flowcell ONT. Made with [www.biorender.com](www.biorender.com)*

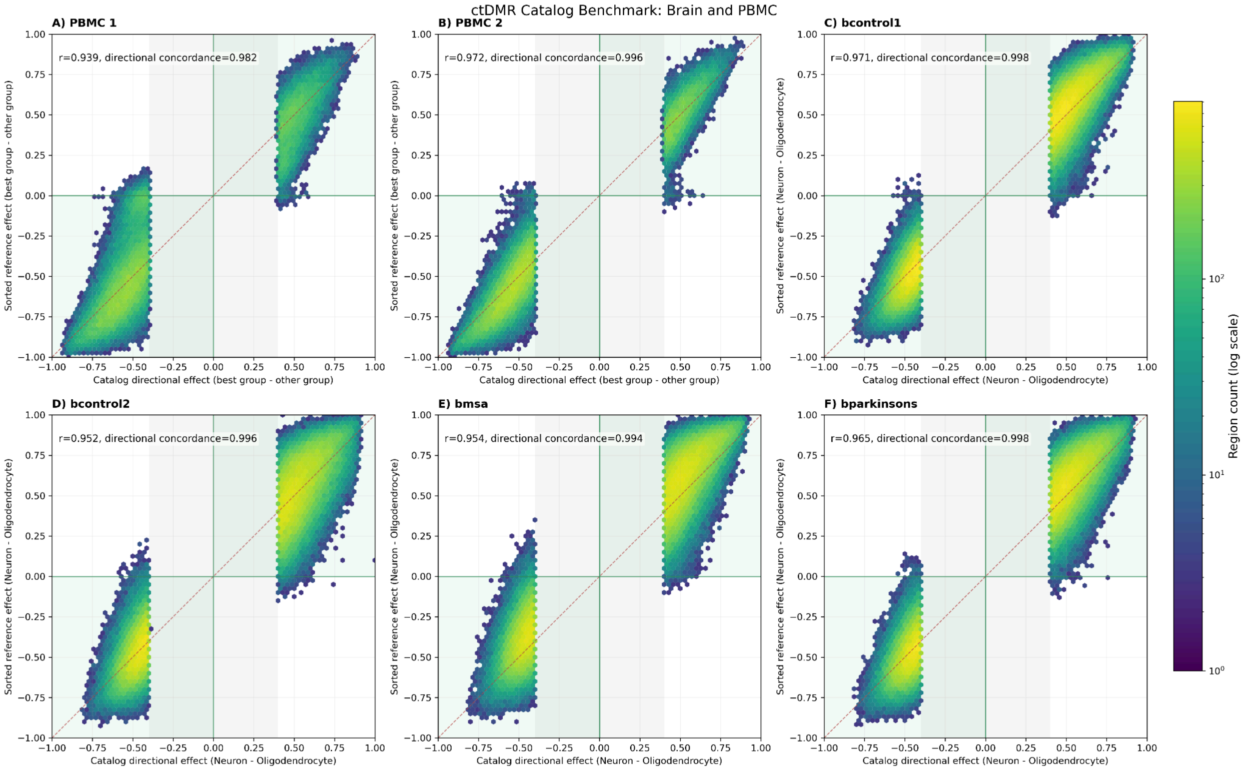


***Supplementary Fig. 4 | Genome-wide per-site benchmarking of the ctDMR discovery from SniffCell find module. a-d. Hexbin plots comparing SniffCell directional effects with FACS/FANS-sorted reference methylomes across four datasets: (a) PBMC1, (b) PBMC2, (c) bcontrol1, (d) bcontrl2 (e) bmsa, and (f) bparkinsons**. Each hexagon represents genomic loci aggregated within ctDMRs, plotted as the directional methylation difference (best group vs other group in **a,b;** neuron vs oligodendrocyte in **c-f**).*

We benchmarked SniffCell find against FACS/FANS-sorted methylation truth sets in brain and PBMC using a unified genome-wide workflow. Modkit pileups were generated once per sample with strand-combined, reference-guided processing, and SniffCell ctDMRs were compared to FACS/FANS truth by direct interval mapping. Region-level benchmarking quantified effect-size concordance, directional concordance, callable ctDMR yield, and per-region DMR overlap, while site-level benchmarking used all significant loci at diff_threshold >= 0.4 without additional segmentation filters (**Supplementary Fig. 4**).

Across all six ctDMR comparisons, SniffCell directional effects showed strong agreement with sorting-derived reference methylation differences (Pearson r = 0.932-0.976; directional concordance = 0.983-0.999), supporting robust recovery of both magnitude and sign of cell-type-specific methylation differences. Directional concordance denotes the proportion of tested ctDMRs for which the sign of the SniffCell directional effect matched the sign of the sorting truth methylation difference, corresponding to regions falling in the concordant quadrants of the hexbin plots (**Supplementary Fig. 4a-f**). In the brain, concordance was high across both controls and disease samples: brain control samples are showing the directional concordance larger than 0.997 (**Supplementary Fig. 4a,b**), while the diseased samples are showing the directional concordance larger than 0.995 (**Supplementary Fig. 4c,d**), which means that SniffCell has the capability to classify the reads in disease settings. Concordance was similarly preserved in PBMC, with PBMC1 showing directional concordance = 0.983, and PBMC2 as 0.999 (**Supplementary Fig. 4e,f**).

Collectively, these results position SniffCell find as the first scalable framework for systematic genome-wide discovery of cell-type–specific methylation signatures, enabling high-resolution characterization of tissue epigenomic landscapes.

### 2. SniffCell deconv and discover for read classifications and variant discovery

#### 2.1. Benchmarking DNA Methylation on SniffCell deconvoluted reads

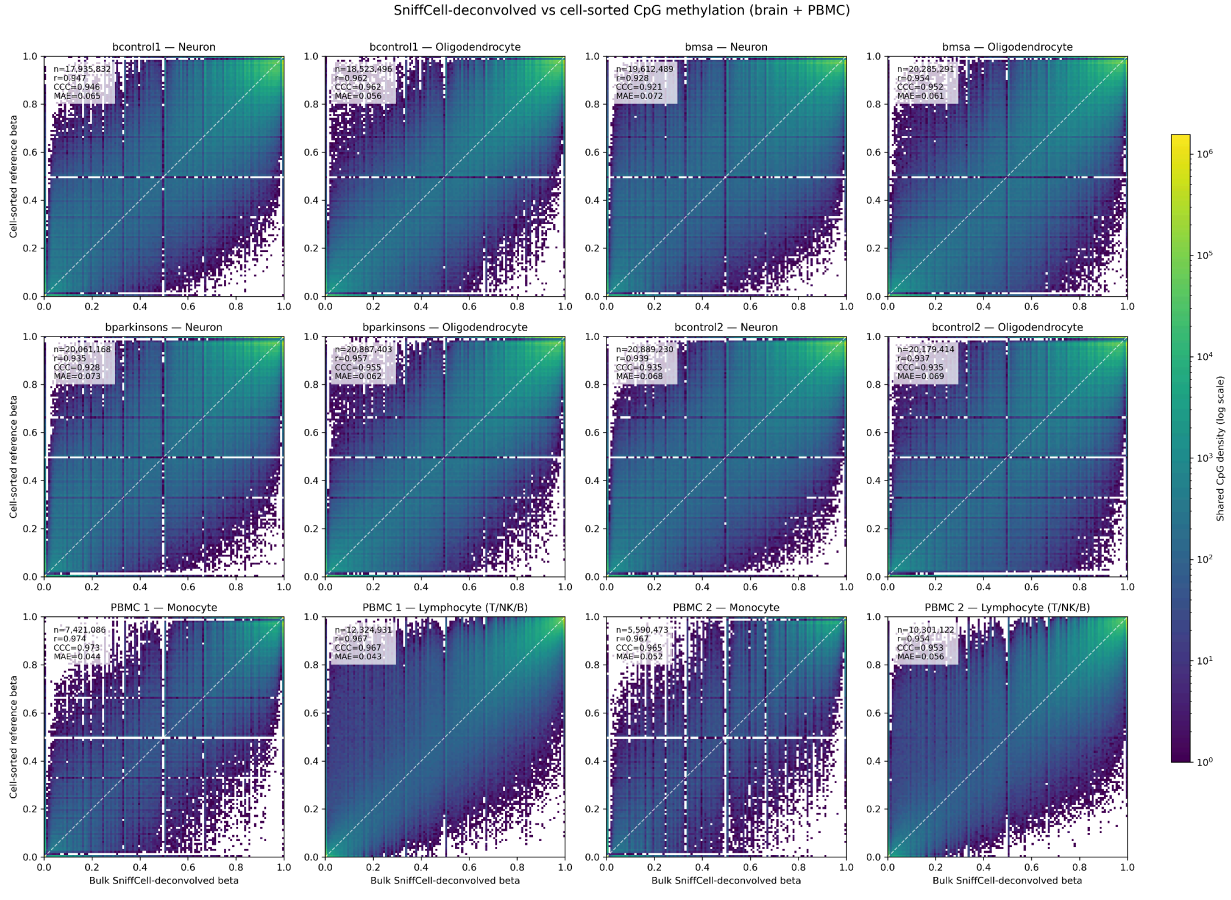


***Supplementary Fig. 5 | Per-CpG concordance of DNA methylation between SniffCell deconvolved split-read BAMs and matched cell-sorted reference samples (log scale). First 10 panels: brain samples, Last 4 panels: PBMC samples***

We first evaluated whether DNA methylation recovered from SniffCell deconvolved split-read BAMs recapitulated methylation profiles from matched cell-sorted reference samples. CpG methylation was extracted from MM/ML modified-base tags using modkit pileup with CpG-aware, strand-combined summaries, and methylation beta values were calculated as modified reads divided by total coverage at each CpG. Brain deconvolution groups were matched to sorted neuron and oligodendrocye references, whereas PBMC monocyte was matched to sorted monocytes and T cells/NK cells/B cells was matched to the combined methylation signal from sorted T cells, NK cells, and B cells. Concordance was assessed at shared CpG sites with coverage greater than 5 reads in both the deconvolved and sorted datasets.

Overall, methylation concordance was high across all benchmarked groups (**Supplementary Fig. 5**). In pooled comparisons, the neuron panel included 27,955,105 shared CpGs and showed Pearson r = 0.933 with MAE = 0.072; oligodendrocytes included 29,493,392 shared CpGs with r = 0.957 and MAE = 0.060; monocytes included 14,766,470 shared CpGs with r = 0.965 and MAE = 0.051; and the combined T cells/NK cells/B cells group included 16,830,776 shared CpGs with r = 0.978 and MAE = 0.041.

Consistent results were observed across the eight individual matched comparisons. Pearson correlation ranged from 0.925 to 0.981 and MAE ranged from 0.036 to 0.075 across 6.5 to 15.6 million shared CpGs. In brain, Pearson r values were 0.945 and 0.961 for b1 neuron and oligodendrocyte, and 0.925 and 0.953 for b2 neuron and oligodendrocyte, with corresponding MAE values of 0.067, 0.058, 0.075, and 0.063. In PBMC, monocyte comparisons yielded r = 0.968 and 0.962 with MAE = 0.047 and 0.056 for pbmc1 and pbmc2, respectively, while the combined T cells/NK cells/B cells groups yielded r = 0.981 and 0.973 with MAE = 0.036 and 0.047.

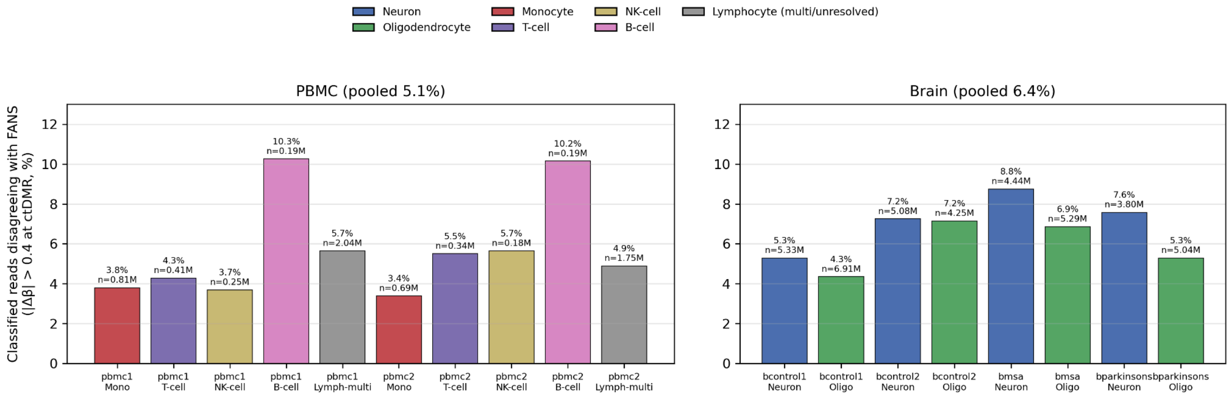


***Supplementary Fig. 6 | Read-level concordance of SniffCell-deconvolved methylation with cell-sorted (*FACS/FANS*) references at ctDMRs.*** *Each bar shows the percentage of classified reads whose own DNA-methylation level at a ctDMR differed from the matched cell-sorted (*FACS/FANS*) reference by more than the 0.4 ctDMR-assignment threshold (|Δβ| > 0.4), for PBMC (left) and brain (right) samples.*

In **Supplementary Fig. 6** Per read methylation was averaged over the CpGs within each ctDMR and compared with the coverage-weighted FACS/FANS reference β of that read's deconv-assigned cell-type at the same ctDMR; the value plotted is the fraction of read x ctDMR observations exceeding the threshold. Bars are colored by cell-type (legend) and annotated with the percentage and the number of read×ctDMR observations. For the reads that could not be resolved to a single subtype in PBMC samples, a combined "Lymphocyte (multi/unresolved)" category compared against the mean of the relevant leaf references; brain samples are labeled bcontrol1, bcontrol2, bmsa, and bparkinsons. Pooled, observation-weighted disagreement was 4.9% for PBMC and 6.3% for the brain (5.9% overall). Disagreement was uniformly low across cell-types (≈3–8%), with the exception of B cells (10.2–10.6%), which were assessed on the fewest reads.

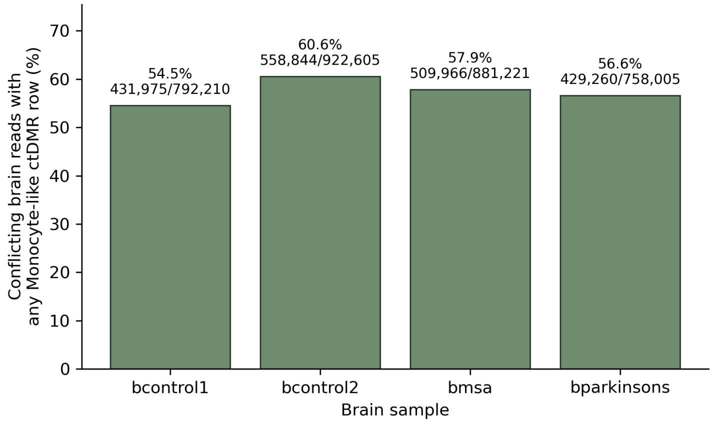


***Supplementary Fig. 7 | monocyte-like ctDMR signal from the reads carrying the conflicting DNA methylation information.***

**Supplementary Fig. 7** shows the percentage of the ctDMR conflicting reads that have a monocyte signal from the ctDMR atlas in 4 brain samples.

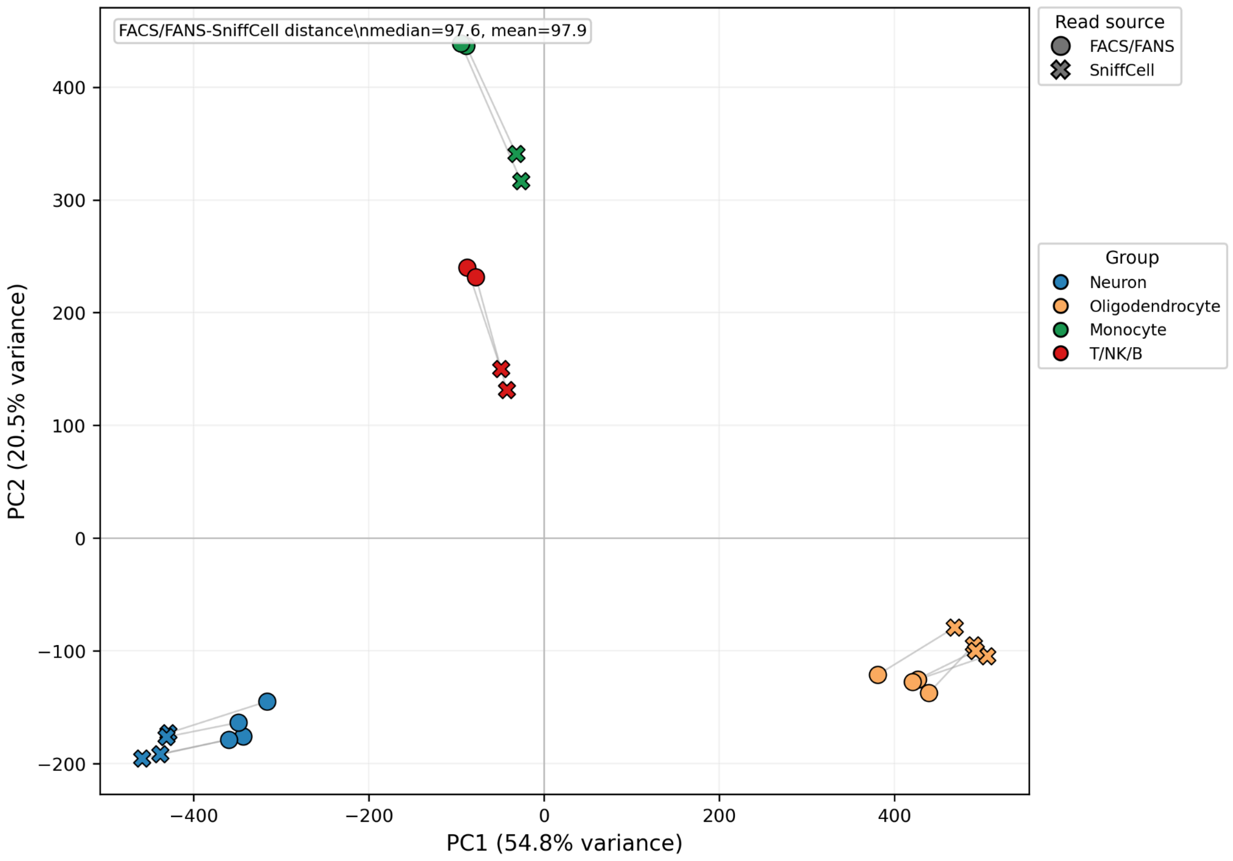


***Supplementary Fig. 8 | PCA plot on both SniffCell deconvoluted reads and FACS/FANS sorted reads. PCA was run on the ctDMR DNA methylation value on the SniffCell deconvoluted reads and FACS/FANS samples' reads.***

**Supplementary Fig. 8** shows the SniffCell deconvoluted reads and sorted reads are showing a high agreement on the PCA plot. We observed a small distance shift between the sorted reads and SniffCell deconvoluted reads.

#### 2.2. Genome-wide coverage of SniffCell with various coverage settings

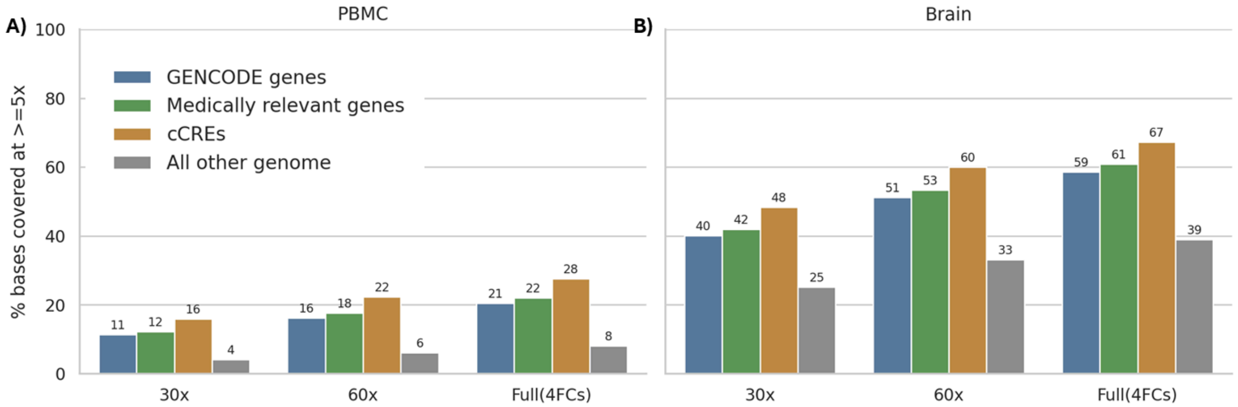


***Supplementary Fig. 9 | A. The percent of bases covered by more than 5X reads on all the GENCODE genes, medically relevant genes and cCRE functional regions in PBMC samples. B. The percent of bases covered by more than 5X reads on all the GENCODE genes, medically relevant genes and cCRE functional regions in Brain samples.***

A gene was considered accessible if at least 80% of its bases were covered by SniffCell-deconvoluted reads at ≥5x. In brain samples, SniffCell-deconvoluted reads covered 67% of genes from GENCODE v49 in the full coverage setting, compared with 39% of matched non-gene intervals. In PBMCs, where we have less ctDMR (**Figure 1H**), 28% of GENCODE genes met this threshold, whereas matched non-gene intervals remained at only 8%. These results show that SniffCell preserves substantial gene-level ascertainment after cell-type deconvolution, enabling cell-type-aware analysis of somatic variants across thousands of genes rather than only isolated loci. **Supplementary Fig. 9** shows how the genomic regions are covered with ctDMRs in various coverage settings.

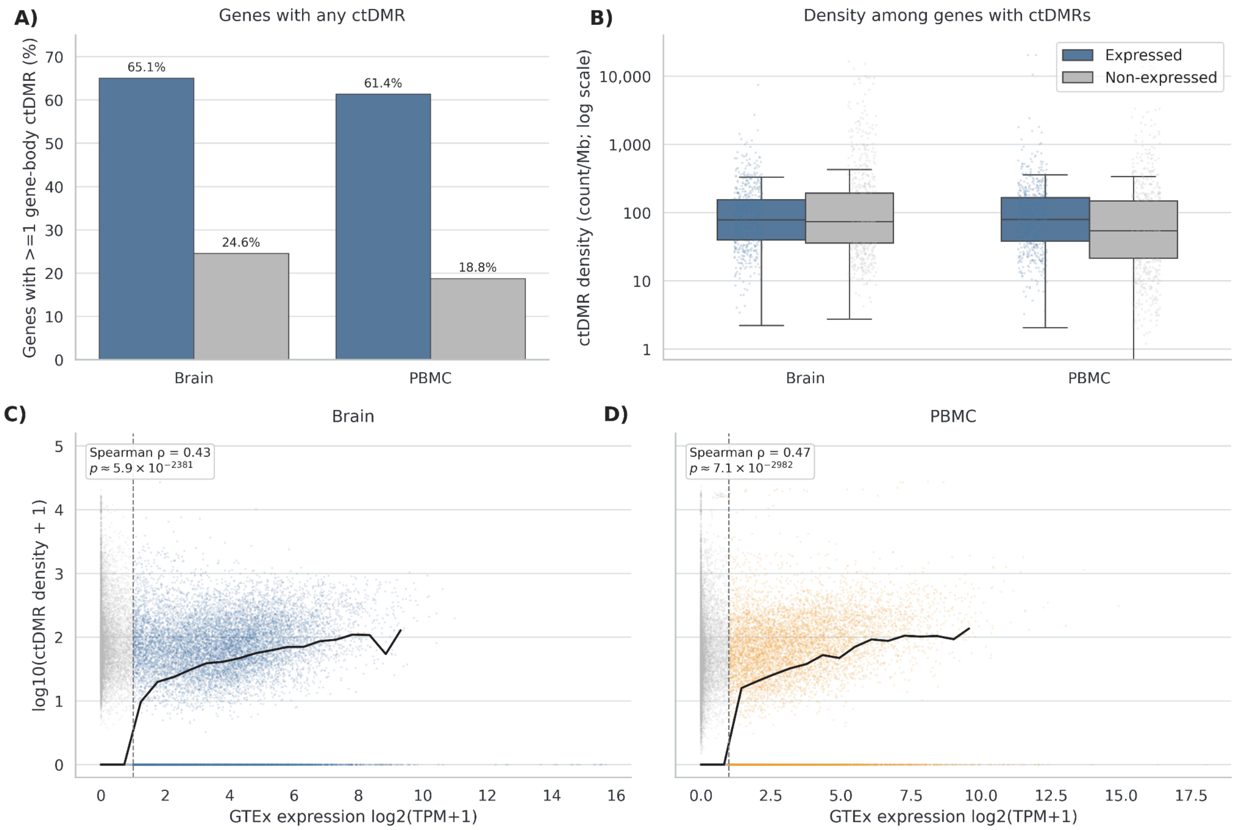


***Supplementary Fig. 10 | ctDMRs exist more frequently in Brain and PBMC expressed genes. A. Over 60% of the brain and PBMC expressed genes in GTEx contain at least one ctDMR, while only 18-24% of the unexpressed genes contain at least one ctDMR. B. For all the genes that have ctDMRs, the distributions of ctDMR density are similar. C. ctDMR density of the brain slightly goes up with the expression from GTEx. D. ctDMR density of PBMC slightly goes up with the expression from blood.***

**Supplementary Fig.** ***10*** shows the expressed genes were much more likely than non-expressed genes to contain at least one gene-body ctDMR in both brain and PBMC. In the brain, 65.1% of expressed genes had a ctDMR compared with 24.6% of non-expressed genes, while in PBMC the corresponding values were 61.4% and 18.8%. Among genes that already contained ctDMRs, ctDMR density showed broad distributions and smaller differences between expressed and non-expressed genes. Across all genes, GTEx expression level was positively associated with ctDMR density in both brain and PBMC, with Spearman correlations of 0.43 and 0.47, respectively.

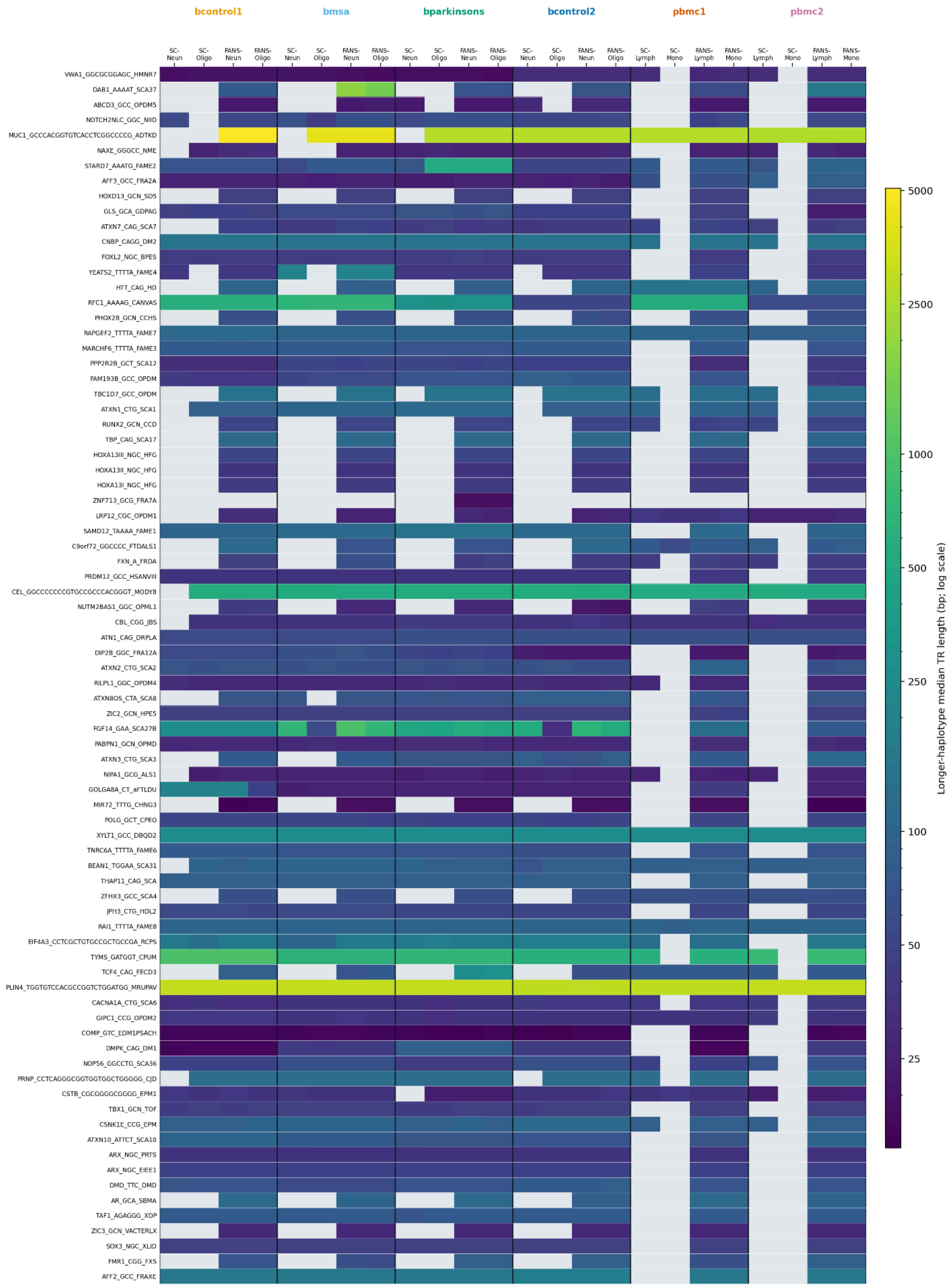

bcontrol1
bmsa
bparkinsons
bcontrol2
pbmc1
pbmc2
SC-Neun
SC-Oligo
FANS-Neun
FANS-Oligo
SC-Lymph
SC-Mono
FANS-Lymph
FANS-Mono
VWA1_GGCGCGGAGC_HMNR7
DAB1_AAAAT_SCA37
ABCD3_GCC_OPDM5
NOTCH2NLC_GGC_NIID
MUC1_GCCCACGGTGTCACCTCGGCCCCG_ADTKD
NAXE_GGGCC_NME
STARD7_AAATG_FAME2
AFF3_GCC_FRA2A
HOXD13_GCN_SD5
GLS_GCA_GDPAG
ATXN7_CAG_SCA7
CNBP_CAGG_DM2
FOXL2_NGC_BPES
YEATS2_TTTTA_FAME4
HTT_CAG_HD
RFC1_AAAAG_CANVAS
PHOX2B_GCN_CCHS
RAPGEF2_TTTTA_FAME7
MARCHF6_TTTTA_FAME3
PPP2R2B_GCT_SCA12
FAM193B_GCC_OPDM
TBC1D7_GCC_OPDM
ATXN1_CTG_SCA1
RUNX2_GCN_CCD
TBP_CAG_SCA17
HOXA13III_NGC_HFG
HOXA13II_NGC_HFG
HOXA13I_NGC_HFG
ZNF713_GCG_FRA7A
LRP12_CGC_OPDM1
SAMD12_TAAAA_FAME1
C9orf72_GGCCCC_FTDALS1
FXN_A_FRDA
PRDM12_GCC_HSANVIII
CEL_GGCCCCCCCGTGCCGCCCACGGGT_MODY8
NUTM2BAS1_GGC_OPML1
CBL_CGG_JBS
ATN1_CAG_DRPLA
DIP2B_GGC_FRA12A
ATXN2_CTG_SCA2
RILPL1_GGC_OPDM4
ATXN8OS_CTA_SCA8
ZIC2_GCN_HPE5
FGF14_GAA_SCA27B
PABPN1_GCN_OPMD
ATXN3_CTG_SCA3
NIPA1_GCG_ALS1
GOLGA8A_CT_aFTLDU
MIR72_TTTG_CHNG3
POLG_GCT_CPEO
XYLT1_GCC_DBQD2
TNRC6A_TTTTA_FAME6
BEAN1_TGGAA_SCA31
THAP11_CAG_SCA
ZFHX3_GCC_SCA4
JPH3_CTG_HDL2
RAI1_TTTTA_FAME8
EIF4A3_CCTCGCTGTGCCGCTGCCGA_RCPS
TYMS_GATGGT_CPUM
TCF4_CAG_FECD3
PLIN4_TGGTGTCCACGCCGGTCTGGATGG_MRUPAV
CACNA1A_CTG_SCA6
GIPC1_CCG_OPDM2
COMP_GTC_EDM1PSACH
DMPK_CAG_DM1
NOP56_GGCCTG_SCA36
PRNP_CCTCAGGGCGGTGGTGGCTGGGGG_CJD
CSTB_CGCGGGGCGGGG_EPM1
TBX1_GCN_TOF
CSNK1E_CCG_EPM
ATXN10_ATTCT_SCA10
ARX_NGC_PRTS
ARX_NGC_EIEE1
DMD_TTC_DMD
AR_GCA_SBMA
TAF1_AGAGGG_XDP
ZIC3_GCN_VACTERLX
SOX3_NGC_XLID
FMR1_CGG_FXS
AFF2_GCC_FRAXE
5000
2500
1000
500
250
100
50
25
Longer-haplotype median TR length (bp; log scale)

***Supplementary Fig. 11 | SniffCell's coverage and calling result on STRchive pathogenic TR variants. SniffCell reaches 35 of 80 loci coverage and 67 of 80 loci coverage in PBMC and brain respectively. The FACS/FANS validation shows good concordance between SniffCell and FACS/FANS sorted reads.***

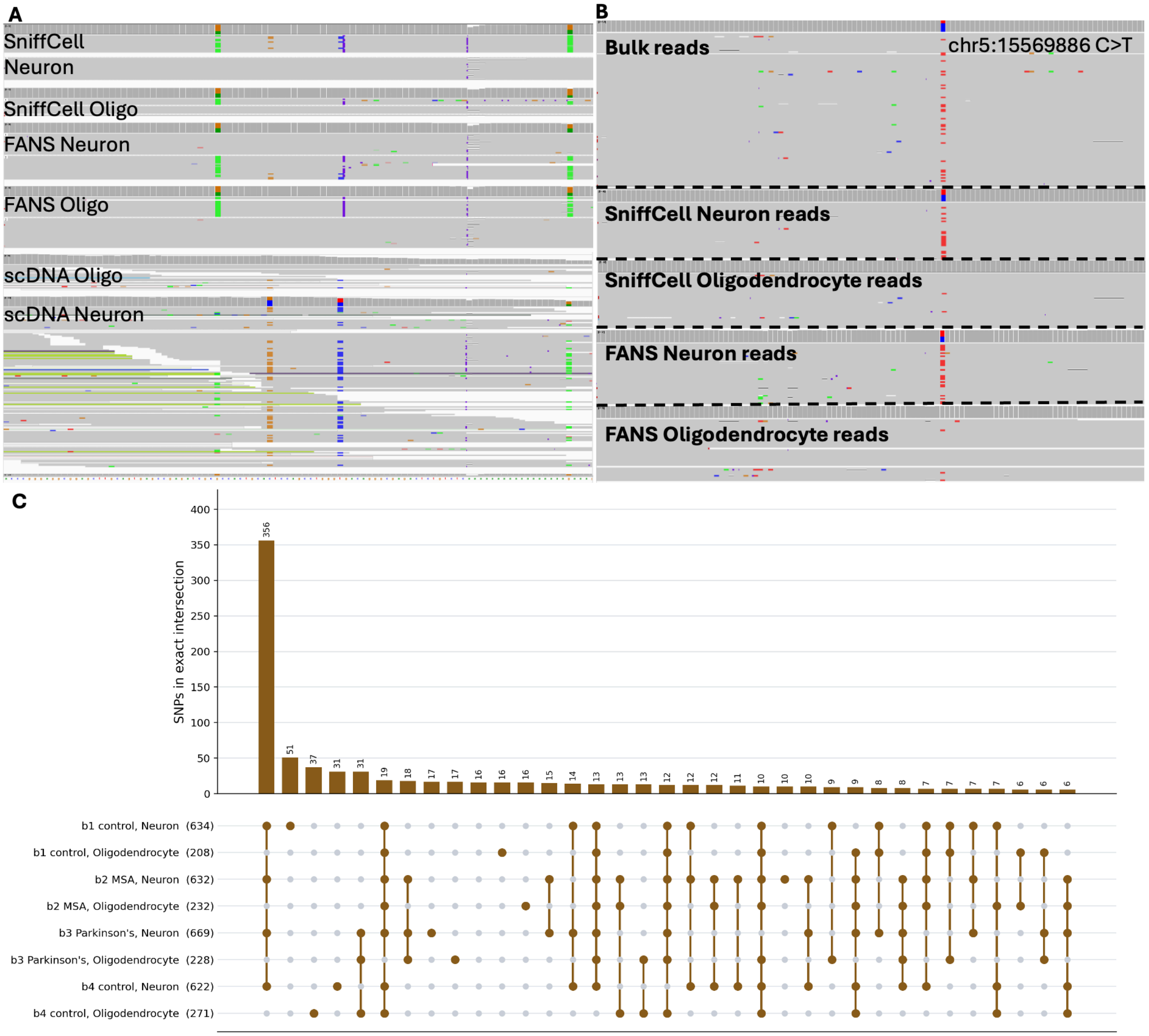


***Supplementary Fig. 12 | A) Potential validated SNV detected by SniffCell B) ONT sequencing artifact at chr5:15569886 C>T from bcontrol1 C) The sharing of "germline-like" SNV detected in neuron SniffCell and FANS brain samples***

**Supplementary Fig. 12** shows a persistent SNV-like signal among the read alignments. However the base quality in that base is in uniform low quality, which indicates the untrustworthiness of the somatic cell-type-specific SNV. However, this SNV was still called a germline SNP in the bulk sequencing data which could lead to false positive downstream analysis. Supplementary Fig. 12B shows we also identified a potential neuron-enriched SNP candidate in *FBXL7* (chr5:15569886 C>T), present in the majority of neuron reads but at very low allele frequency in oligodendrocyte reads. The SNP candidate is also enriched in the FANS sorted neuron sample and not shown in the oligodendrocyte. However, the base quality at this site was low, suggesting that the signal may reflect an Oxford Nanopore basecalling artifact, potentially related to active non-CpG methylation in brain neurons. Interestingly, the same variant was called as germline in the bulk sample, highlighting the possibility that some variants classified as germline in bulk sequencing may instead reflect cell-type-enriched signals.

#### 2.3. PBMC variant discovery from SniffCell deconvoluted reads

SniffCell identified cell-type-specific structural variants (SVs) and tandem repeats (TRs) from deconvoluted reads. Variant discovery was performed within each inferred cell-type group, followed by post hoc comparative analysis across cell-types. SV detection and filtering followed a Sniffles-, Truvari-, and Kanpig-based pipeline, whereas TR analysis used intermediate TRGT outputs to recover read-level repeat lengths. We benchmarked the resulting variant calls against FACS-sorted samples using CIGAR-string matching and manual IGV inspection. Our evaluation focused on the flip rate, defined as cases where a read carrying a cell-type-specific SV or TR expansion is assigned by SniffCell to an opposing cell-type. For example, an SV or TR expansion originating from monocytes in a PBMC sample would be considered a flip if SniffCell assigned the read to T cells, B cells, or NK cells.

We first analyzed the PBMC samples. The PBMC samples contain 2 control panels which serves perfectly as our benchmarking dataset. We first investigated the somatic recombination (V(D)J recombination) sites in the immune system. The V(D)J recombination has an expected length range which can be derived from the genes' locations on the reference genome. **Supplementary Table 3** shows the expected length range of the deletions which are reflected as the ranges in **Figure 3A**.

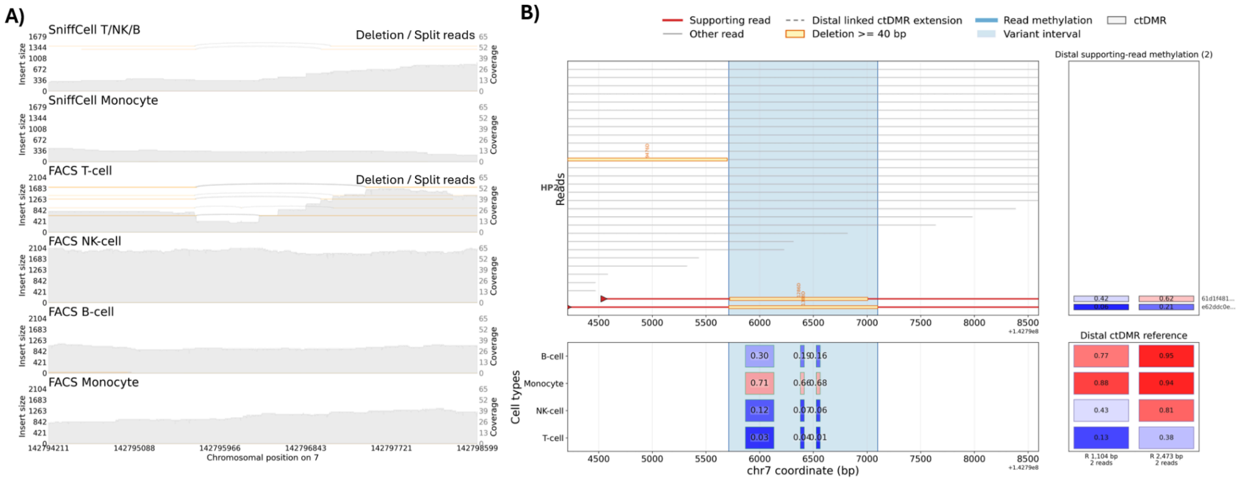


***Supplementary Fig. 13 | Samplot of SV001. A)*** *Somatic deletions are classified as Lymphocytes. FACS T-cell shows the deletion is only restricted to T cells at this TRB region.* ***B)*** *SniffCell anno and SniffCell viz annotates the variant as T cell due to the matched DNA methylation in two downstream ctDMRs.*

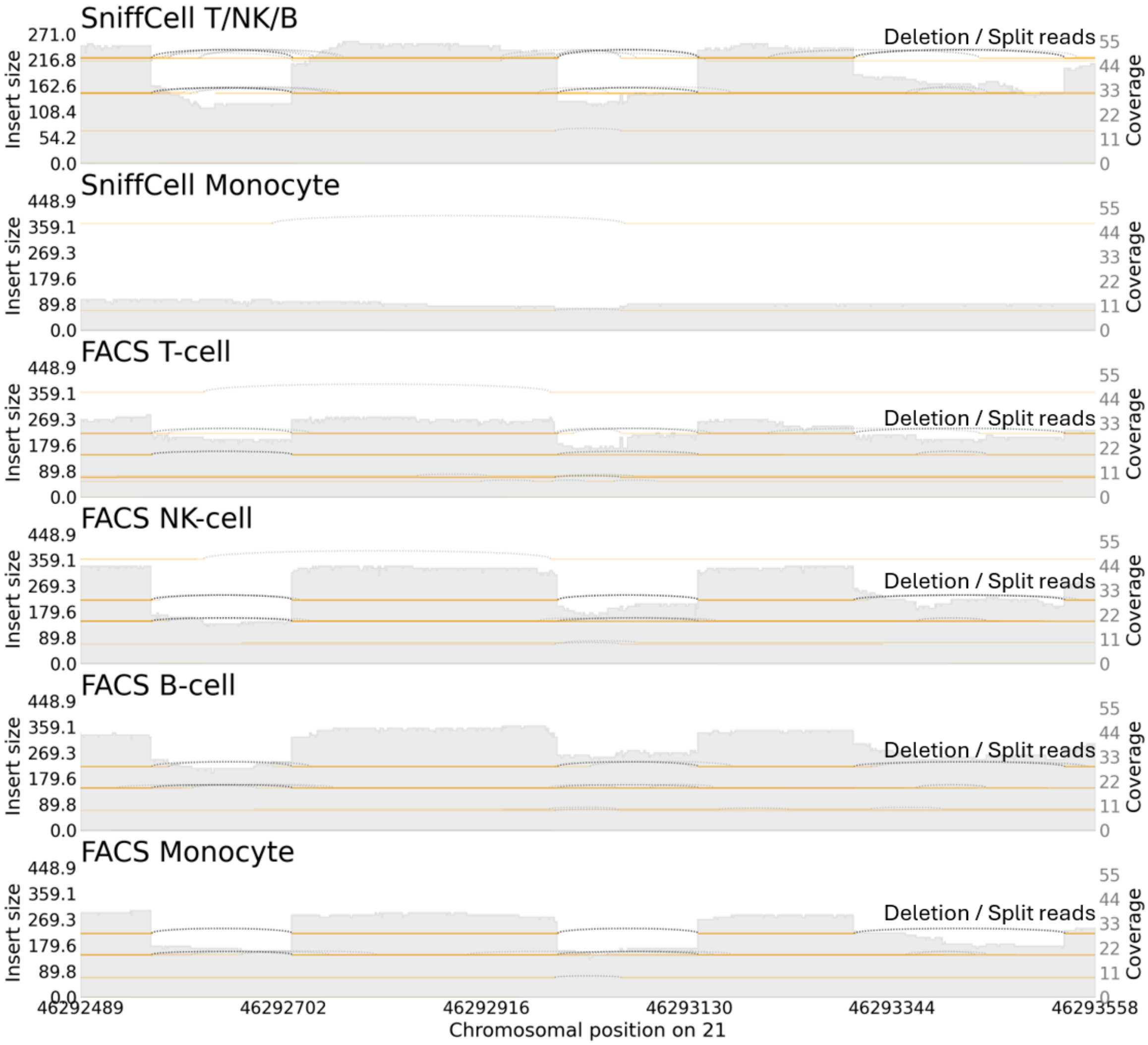


**Supplementary Fig. 14 | Samplot of SV002 at pbmc2.** Somatic deletions are classified as Lymphocytes. FACS validation shows the somatic mutation also shows up at monocyte reads.

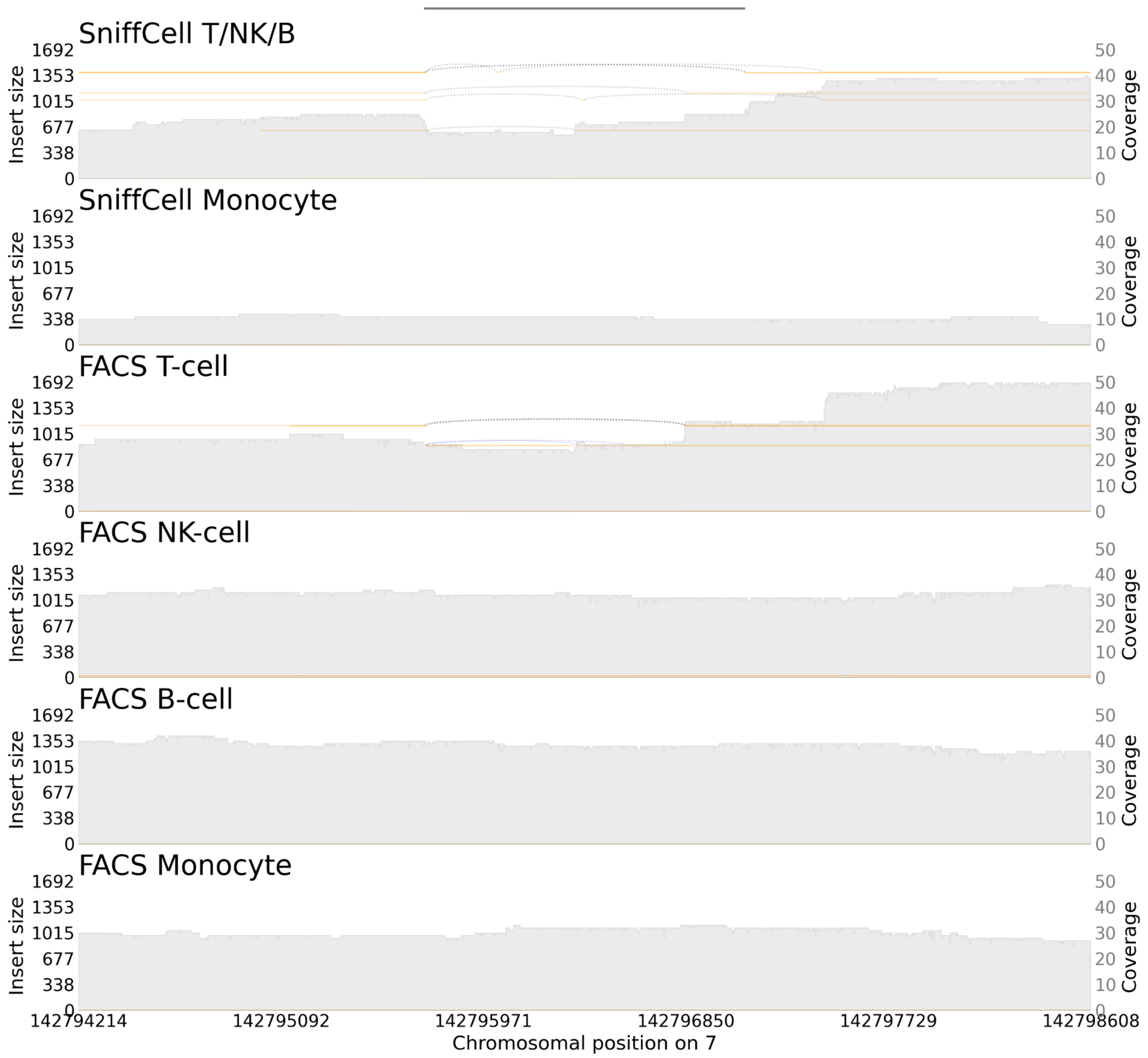


**Supplementary Fig. 15 | Samplot of SV003 at pbmc2.** Somatic deletions are classified as Lymphocytes. SniffCell assigns reads to T cell based on further DNA methylation classification.

#### 2.4. Brain variant discovery from SniffCell deconvoluted reads

##### 2.4.1. Brain SVs

Brain SVs are shown as IGV screenshots due to lack of support for displaying insertion in samplot.

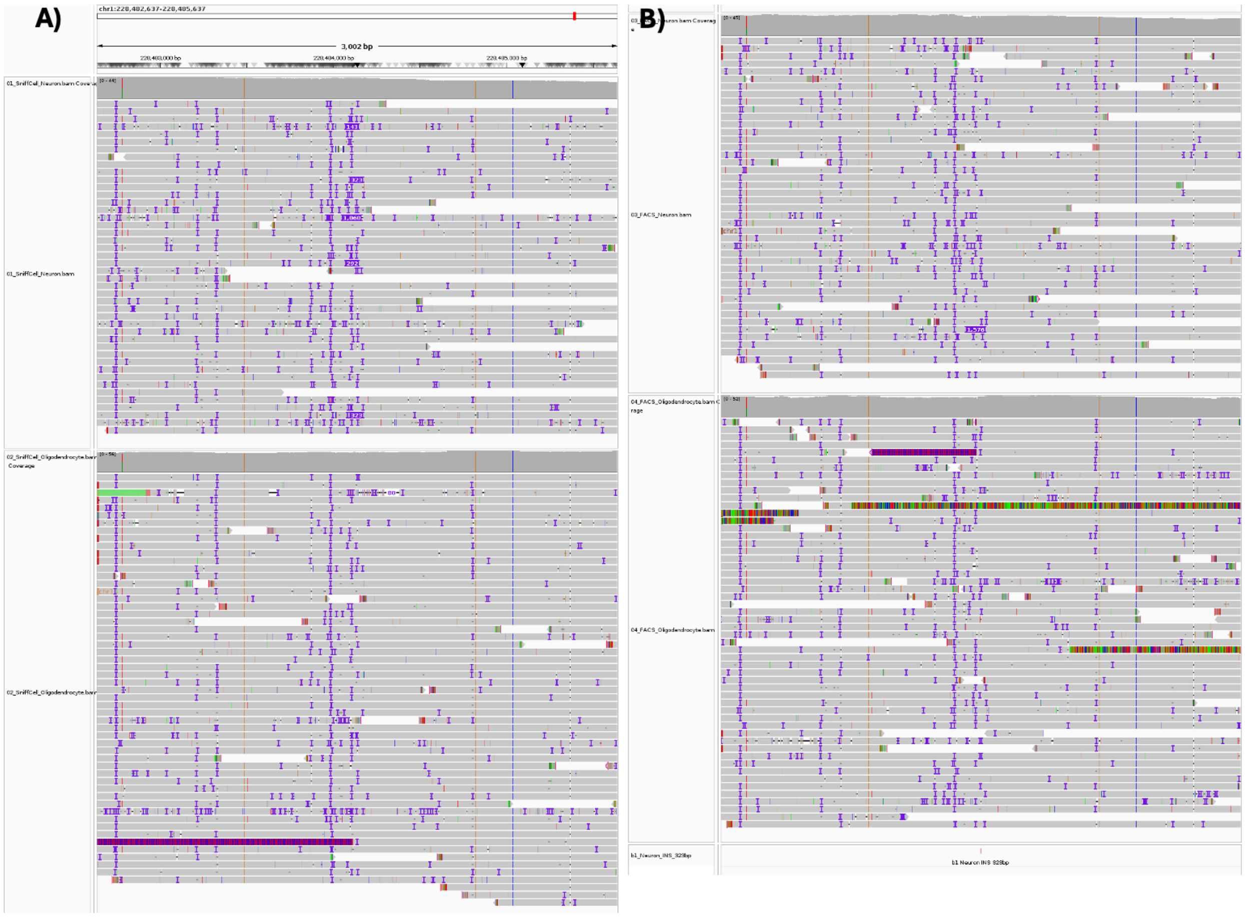


**Supplementary Fig. 16 | IGV screenshot of SV004. A)** The insertion is called by SniffCell as a somatic INS at neuron. **B)** The insertion is also observed as an insertion by one read in FANS neuron cells.

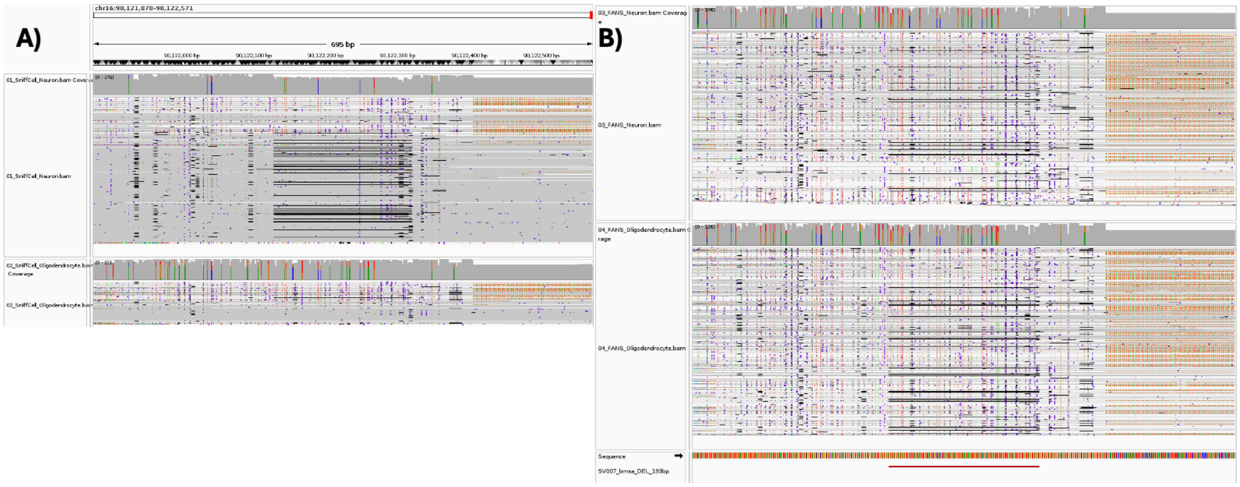


**Supplementary Fig. 17 | IGV screenshot of SV005. A)** The somatic deletion is present and restricted in SniffCell-classified neuron BAMs with only low quality aligned reads at

Oligodendrocyte reads as false positive assignment but does not affect the somatic SV calling.
**B)** The deletion is present in FANS sorted BAM for both cell types.

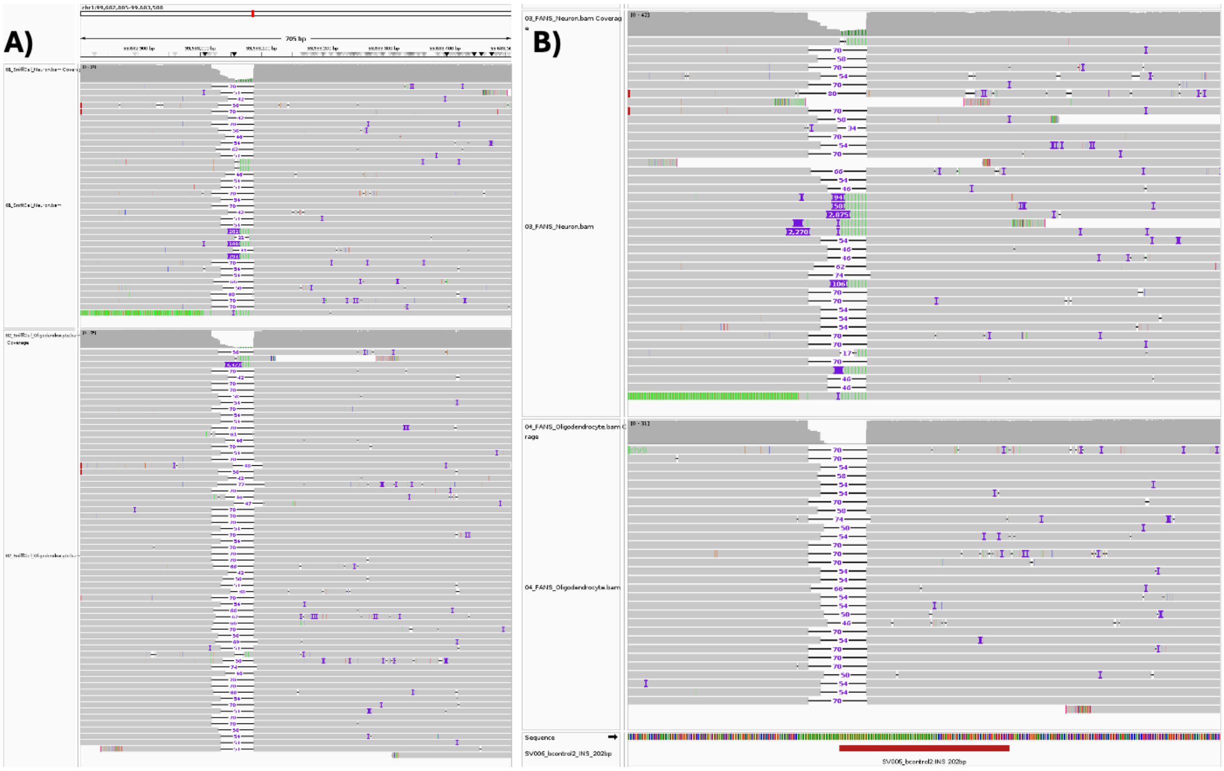


**Supplementary Fig. 18 | IGV screenshot of SV006. A)** The somatic insertion is present and restricted in SniffCell-classified neuron BAMs with one read at Oligodendrocyte reads as false positive assignment but does not affect the somatic SV calling. **B)** The somatic insertion is present and restricted in FANS neuron BAMs.

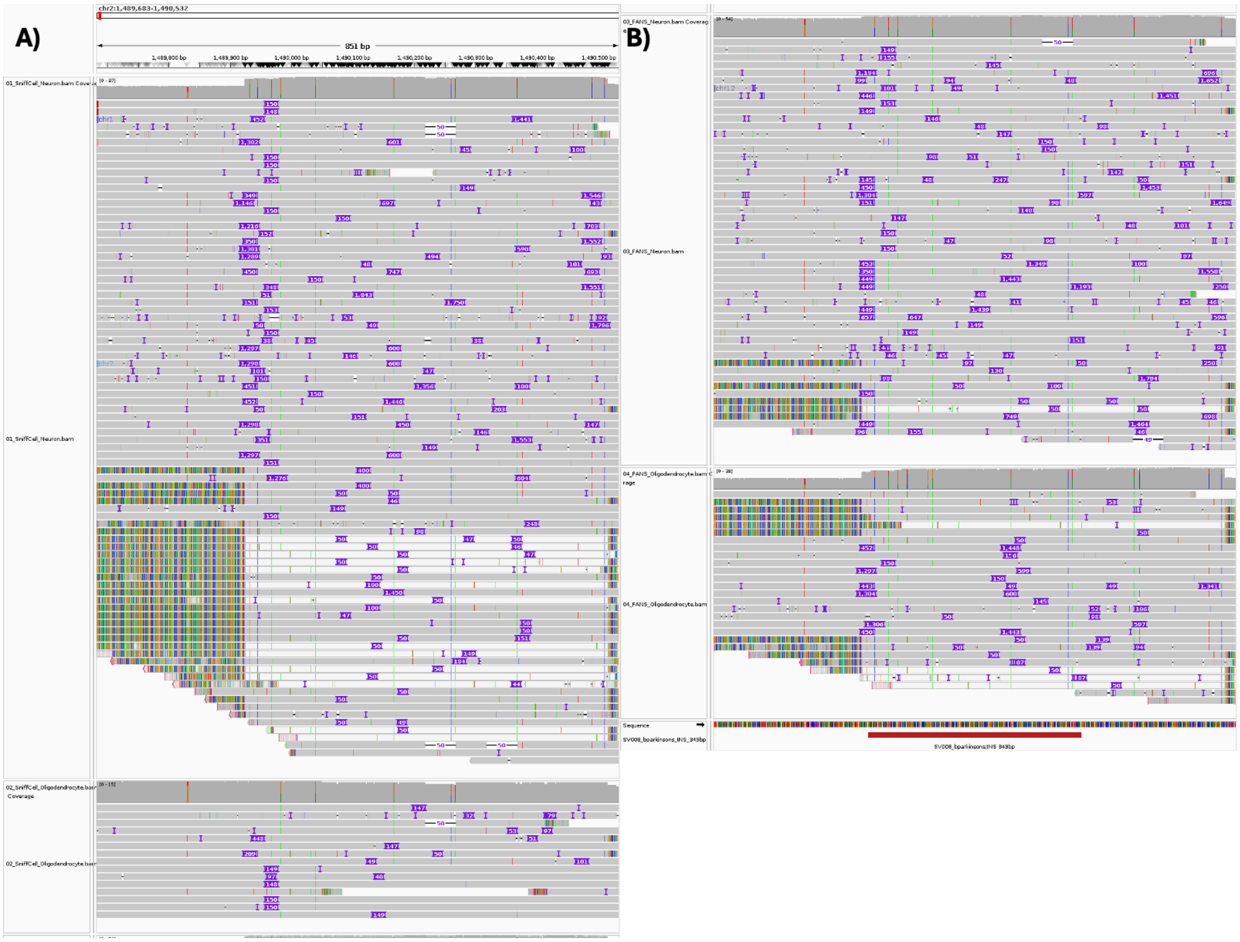


**Supplementary Fig. 19 | IGV screenshot of SV007. A)** The somatic insertion of 349 bp is present and restricted in SniffCell-classified neuron BAMs.**B)** The somatic insertion is present and restricted in FANS neuron BAMs.

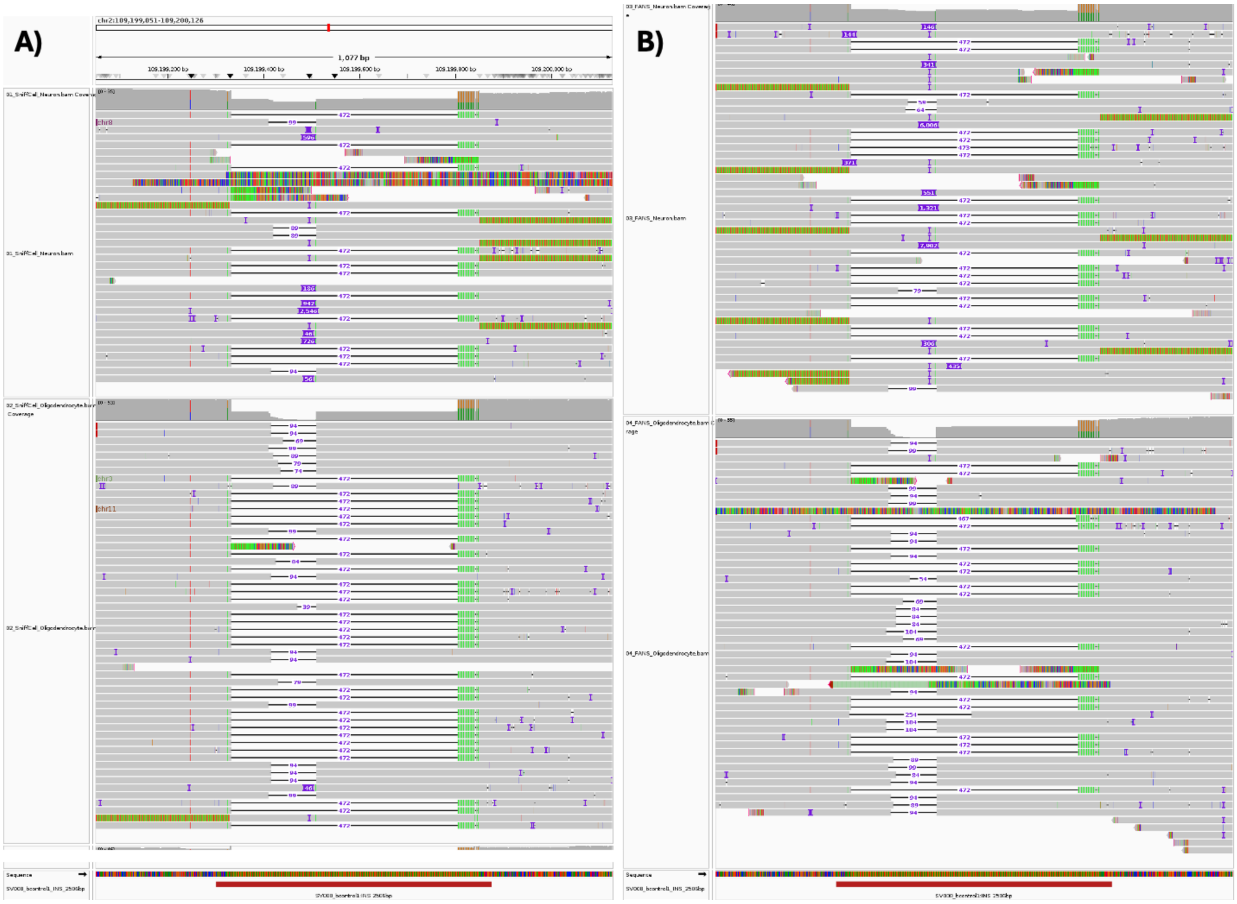

**Supplementary Fig. 20 | IGV screenshot of SV008. A)** The somatic insertion is present and restricted in SniffCell-classified neuron BAMs with one read at Oligodendrocyte reads as false positive assignment but does not affect the somatic SV calling. **B)** The somatic insertion is present and restricted in FANS neuron BAMs.

##### 2.4.2. SGIP1 neuron expansion

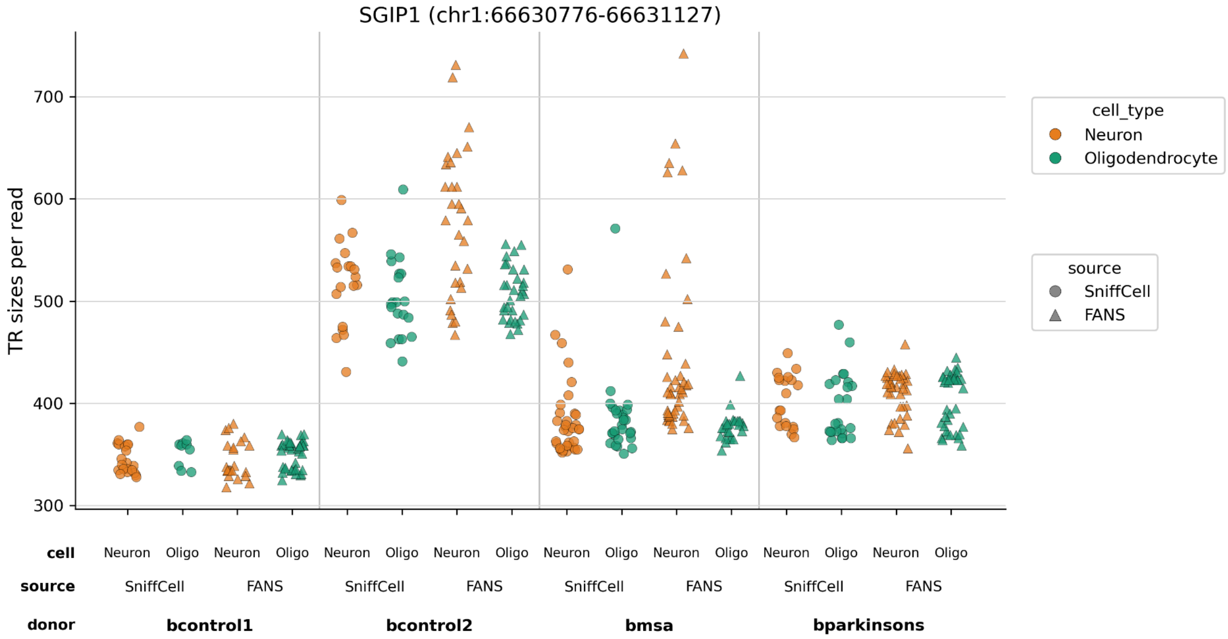


**Supplementary Fig. 21 | SGIP1 repeat expansion in 4 brain benchmarking samples.**

##### 2.4.3. The lack of FGF14 flanking variant in somatically expanded alleles

To investigate the relationship between the 5' common flanking variant (5'-CFV, chr13:102,161,544-102,161,756, TAGTCATAGTACCCCAA) described by Pellerin et al.[33] and cell-type-specific somatic instability at the *FGF14* GAA repeat locus, we examined phased allele sequences from SniffCell-deconvolved and FANS-sorted neuronal and oligodendrocyte fractions across all samples in our cohort. Consistent with the reported population allele frequency of ~70%, the 5'-CFV was detected on at least one allele in every sample examined. In all cases, the 5'-CFV was carried exclusively on the shorter, non-expanded allele (~58–70 estimated GAA repeats), and was entirely absent from the expanded allele, which replicates at the single-cell-type level the mutually exclusive relationship between the 5'-CFV and pathogenic expansions reported in germline studies[33]. The 5'-CFV allele showed uniformly stable read-length distributions across neuronal and oligodendrocyte fractions (range 694–738 bp), with no evidence of cell-type-specific somatic length change. In contrast, the non-FV expanded allele exhibited pronounced neuronal somatic instability in affected individuals, with read lengths spanning 1,020–2,114 bp in neurons compared to 1,385–1,428 bp in oligodendrocytes from the same donors. These findings demonstrate that the stabilizing effect of the 5'-CFV is cell-type-autonomous: the variant confers repeat stability on its own allele even within neurons that simultaneously harbor a somatically-expanding allele on the homologous chromosome.

#### 3.1 SMaHT brain cohort analysis

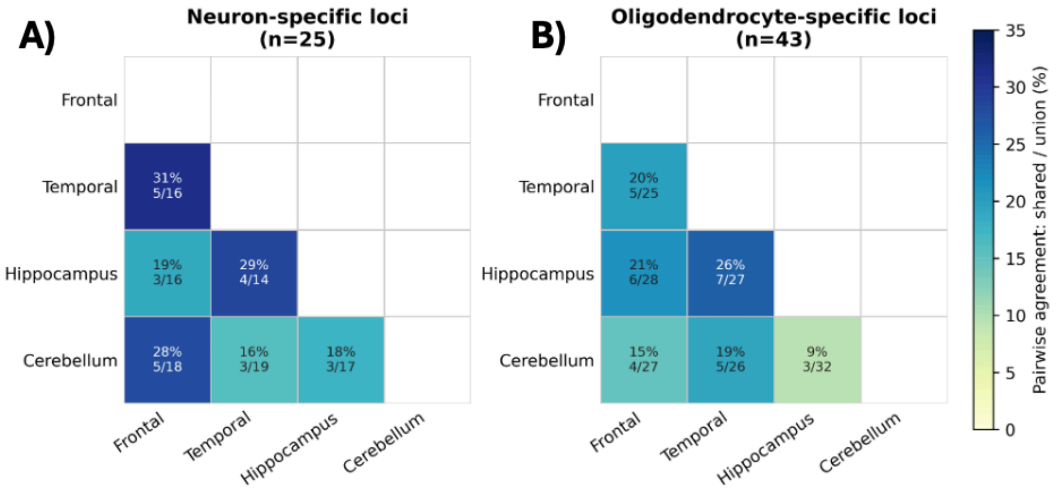


**Supplementary Fig. 22 | Pairwise overlap of all cell-type-specific TR expansion loci across brain regions stratified by cell-type. A) Pairwise overlap of all cell-type-specific TR expansion loci across brain regions of neuron-specific expansion loci. B) Pairwise overlap of all cell-type-specific TR expansion loci across brain regions of oligodendrocyte-specific expansion loci.**

#### 3.2 CARD brain cohort analysis

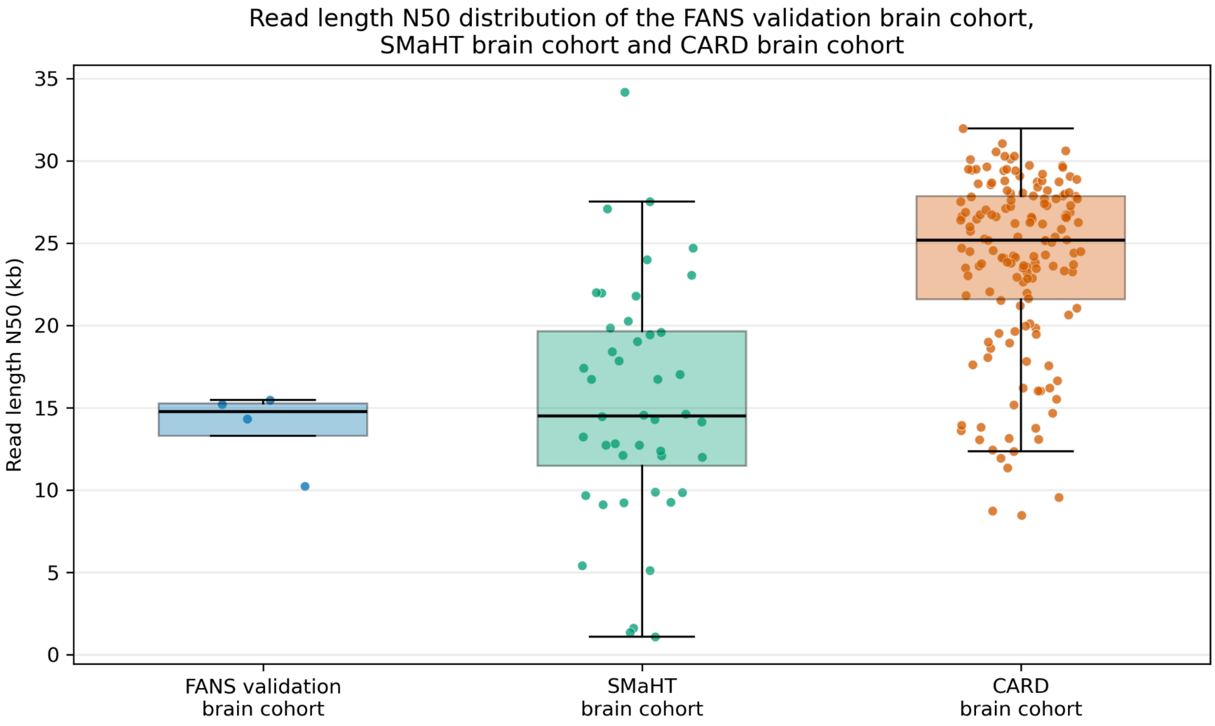


**Supplementary Fig. 23 | Read length N50 distribution of the FANS validation brain cohort, SMaHT brain cohort and CARD brain cohort.**

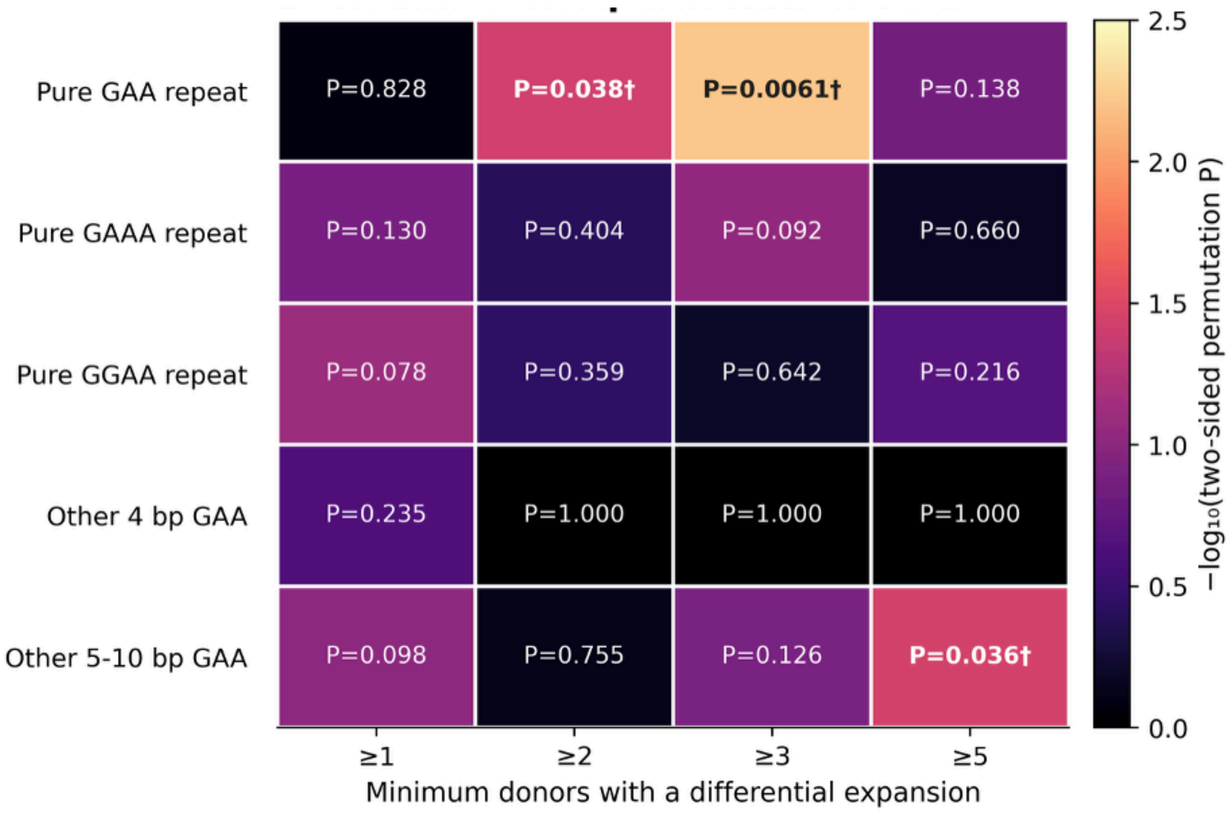


**Supplementary Fig. 24 | A) permutation test of CARD brains cohort GAA-rich neuron enrichment.**

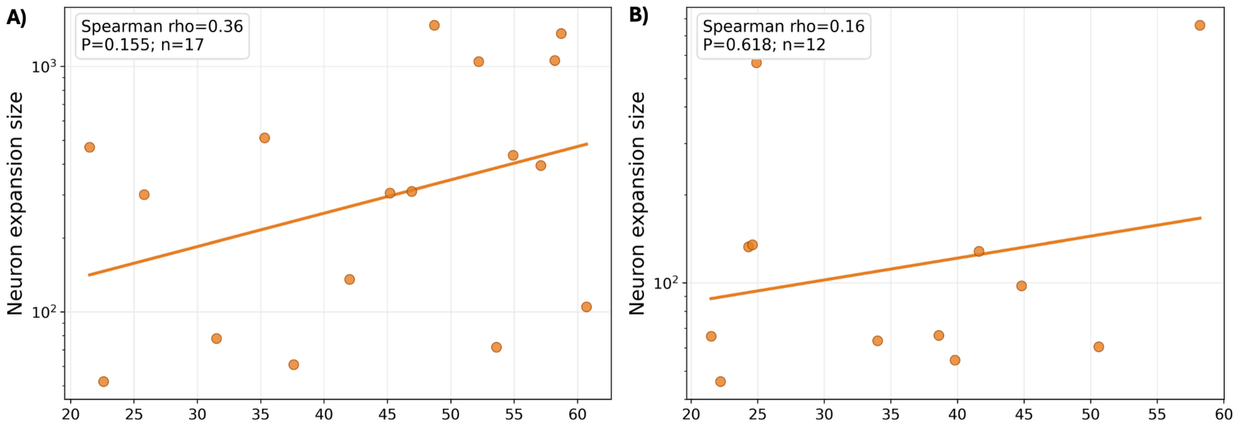


**Supplementary Fig. 25 | Age associations of neuron-specific differential expansions at SH3RF3 and LINGO2. A) Association between age at death and neuron-specific differential expansion at the SH3RF3 AATGG repeat (chr2:109,199,301–109,199,876).** Each point represents one of 17 donors with a Neuron-specific call. Expansion magnitude is expressed in AATGG motif units as the maximum Neuron–Oligodendrocyte read-length difference divided by the 5-bp motif length (Spearman ρ = 0.36, P = 0.155). **B) Association between age at death and neuron-specific differential expansion at the LINGO2 AGGA repeat (chr9:28,189,170–28,189,834).** Each point represents one of 12 donors with a Neuron-specific call. Expansion magnitude is expressed in AGGA motif units as the maximum Neuron–Oligodendrocyte read-length difference divided by the 4-bp motif length (Spearman ρ = 0.16, P = 0.618). Orange lines indicate log-linear fits.

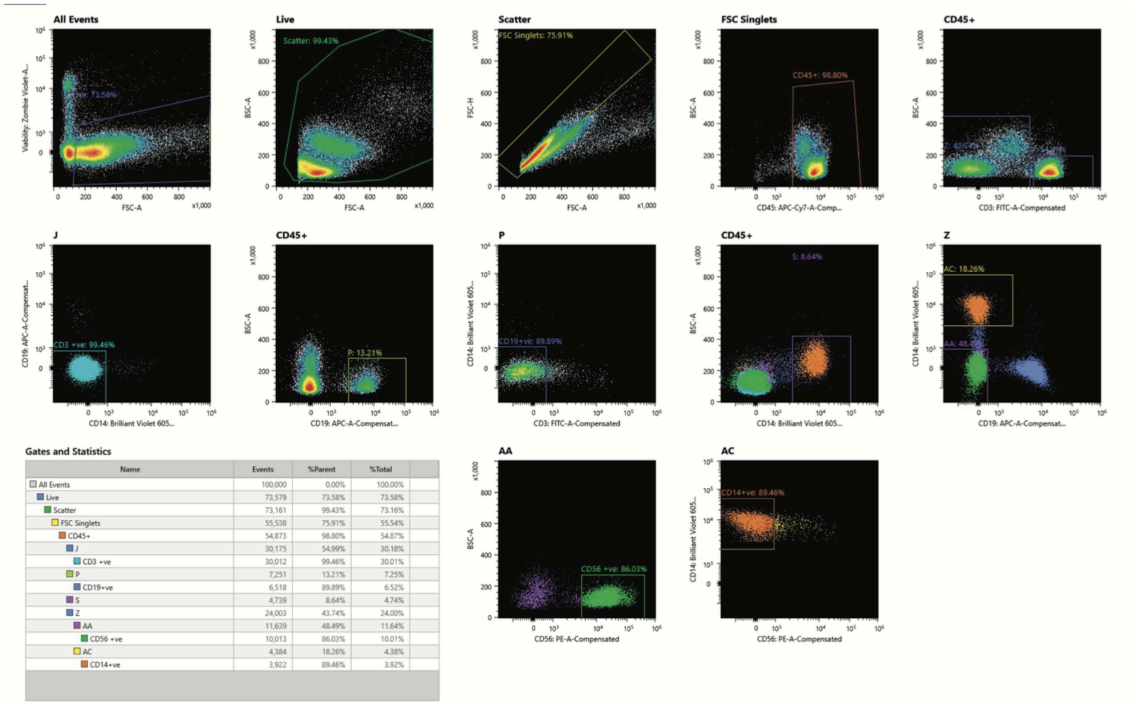


| Name | Events | %Parent | %Total |
|---|---|---|---|
| All Events | 100,000 | 0.00% | 100.00% |
| Live | 73,579 | 73.58% | 73.58% |
| Scatter | 73,161 | 99.43% | 73.16% |
| FSC Singlets | 55,538 | 75.91% | 55.54% |
| CD45+ | 54,873 | 98.80% | 54.87% |
| J | 30,175 | 54.99% | 30.18% |
| CD3 +ve | 30,012 | 99.46% | 30.01% |
| P | 7,251 | 13.21% | 7.25% |
| CD19+ve | 6,518 | 89.89% | 6.52% |
| S | 4,739 | 8.64% | 4.74% |
| Z | 24,003 | 43.74% | 24.00% |
| AA | 11,639 | 48.49% | 11.64% |
| CD56 +ve | 10,013 | 86.03% | 10.01% |
| AC | 4,384 | 18.26% | 4.38% |
| CD14+ve | 3,922 | 89.46% | 3.92% |

**Supplementary Fig. 26 | Fluorescence-activated cell sorting (FACS) gating strategy for isolation of human PBMCs.** *Hierarchical gating strategy used to identify target populations. Cells were sequentially gated on live cells, scatter profile (FSC-A vs BSC-A), and singlets (FSC-H vs FSC-A), then on CD45$^{+}$ leukocytes. Four lineages were resolved and collected: T cells ("CD3 +ve"), B cells ("CD19 +ve"), NK cells ("CD56 +ve"), and monocytes ("CD14 +ve"). Numbers on each plot indicate the frequency of the gated population as a percentage of its parent gate. The accompanying table reports event count, % of parent, and % of total for each gate (out of 100,000 recorded events). Gating and sorting was performed on Sony M900.*
